# Functional genomics in primary T cells and monocytes identifies mechanisms by which genetic susceptibility loci influence systemic sclerosis risk

**DOI:** 10.1101/2022.05.08.22274711

**Authors:** David González-Serna, Chenfu Shi, Martin Kerick, Jenny Hankinson, James Ding, Amanda McGovern, Mauro Tutino, Gonzalo Villanueva Martin, Norberto Ortego-Centeno, José Luis Callejas, Javier Martin, Gisela Orozco

## Abstract

**Objectives:** Systemic sclerosis (SSc) is a complex autoimmune disease with a strong genetic component. However, most of the genes associated to the disease are still unknown because associated variants affect mostly non-coding intergenic elements of the genome. The challenge now is to use functional genomics to translate the genetic findings into a better understanding of the disease.

**Methods:** Promoter capture Hi-C and RNA sequencing experiments were performed in CD4^+^ T cells and CD14^+^ monocytes samples from 10 SSc patients and 5 healthy controls to link SSc-associated variants with their target genes, followed by differential expression and differential interaction analyses between cell types.

**Results:** We linked SSc-associated loci to 39 new potential target genes and confirm 7 previously known genes. We highlight novel causal genes, such as *CXCR5* as the most probable candidate gene for the *DDX6* locus. Some previously known SSc associated genes such as *IRF8, STAT4*, or *CD247* interestingly showed cell type specific interactions. We also identified 15 potential drug targets already in use in other similar immune-mediated diseases that could be repurposed for SSc treatment. Furthermore, we observed that interactions are directly correlated with the expression of important genes implicated in cell type specific pathways and find evidence that chromatin conformation is associated with genotype.

**Conclusions:** Our study reveals potential causal genes for SSc-associated loci, some of them acting in a cell type specific manner, suggesting novel biological mechanisms that might mediate SSc pathogenesis.

## INTRODUCTION

Systemic sclerosis (SSc) is a complex chronic immune-mediated disease that affects the connective tissue, characterized by an immune imbalance, vascular alterations, and an excessive collagen deposition leading to fibrosis (1). Several lines of evidence implicate T cells and monocytes/macrophages as important cell types in SSc pathogenesis (2,3). In this regard, modifications in the proportions of CD4^+^ T cell subpopulations, and their functional alterations may contribute to the vascular dysregulation and fibrosis observed in the disease (4,5). On the other hand, circulating monocytes/macrophages with a profibrotic phenotype are increased in blood from SSc patients (6,7), and changes in human monocyte-derived macrophages transcriptome are related to SSc genetic variants (8).

SSc presents a complex genetic component, and its etiology is poorly understood. Large-scale genetic studies have so far identified 27 independent signals associated with susceptibility to SSc (9,10). Interestingly, many of the different genes assigned to SSc GWAS loci are related to T cell activation and macrophages regulation pathways (11,12), and are shared among immune-mediated diseases, which may be of interest in the drug repositioning of rare diseases like SSc, where there are no specific available treatments (13). However, the majority of single nucleotide polymorphisms (SNPs) associated with SSc map to non-coding regions of the genome that are enriched in enhancer elements, which are often cell-type specific (14,15). These regulatory elements can interact with genes often located hundreds of kilobases away, bypassing in many cases nearby genes (16).

Thus, the current challenge remains in linking disease-associated regions with the genes that they affect, in the specific cell types involved, in order to pinpoint to the mechanisms and the biological pathways implicated in genetically susceptible patients (17). In this regard, many techniques to analyze the three-dimensional genome architecture have emerged, such as chromosome conformation capture (3C) (18,19), that can help us annotate these genetic variants. The most powerful technique developed to date, Hi-C allows the identification of chromosomal interactions genome-wide (20). A more recent technique, capture Hi-C (CHi-C) allows to specifically enrich chromosomal regions of interest, such as disease risk loci (region CHi-C) or promoters (promoter CHi-C, pCHi-C) from Hi-C libraries in a cost-effective way (21). This technique has been successfully applied in different cell types to link enhancers and non-coding disease variants to potential target genes (22). Previously, we successfully applied CHi-C to identify disease causal genes and potential drugs for repositioning in autoimmune diseases using cell lines (23,24). Since the regulation of gene expression and chromatin conformation is highly context specific, it is essential to apply these technologies to primary cells isolated from patients. Moreover, recent evidence points to alterations in chromatin conformation linked to genotype, but these studies have so far only been carried out in a limited way and in cell lines (25,26).

In this study, we used pCHi-C in two of the most relevant cell types in SSc pathogenesis: CD4^+^ T cells and CD14^+^ monocytes from SSc patients and healthy controls in an attempt to annotate gene targets within all known SSc GWAS loci. We also integrate this data with RNA-seq, eQTLs and genotype information to create a multi-omic approach in order to identify interactomic and transcriptomic differences between cell types and disease state that could be of interest in the pathogenesis of SSc.

## MATERIAL AND METHODS

Please see **Supplementary Material and Methods** for more details.

### Isolation of CD4^+^ T cells and CD14^+^ monocytes

Primary CD4^+^ T cells and CD14^+^ monocytes were collected from 10 systemic sclerosis patients and 5 healthy individuals. All SSc patients were diagnosed according to the American College of Rheumatology (ACR)/European Alliance of Associations for Rheumatology (EULAR) 2013 criteria (cohorts characteristics described in **Supplementary Table 1**). All patients and controls gave written informed consent, which was approved by the local ethics committees. Peripheral blood mononuclear cells (PBMCs) were isolated from 70 ml blood samples using Ficoll density gradient centrifugation. The EasySep Human CD14^+^ Positive selection kit (StemCell Technologies, ref:17858) was used to isolate CD14^+^ cells from PBMCs and, subsequently, the Easysep CD4^+^ T cell isolation kit (StemCell Technologies, ref:17952) was used to isolate CD4^+^ T cells from the remaining PBMCs, according to the manufacturer’s instructions.

### Capture Hi-C library generation and processing

5-10 million isolated CD4^+^ T cells and CD14^+^ monocytes were crosslinked in 1% formaldehyde; the reaction was then quenched with 0.125 M glycine. Each Hi-C library was prepared from fixed cells following the Arima HiC kit (Arima Genomics) and the KAPA HyperPrep kit (KAPA Biosystems) following the manufacturer’s protocol. Hi-C samples were then hybridized with the SureSelect custom capture library by following Agilent SureSelectXT HS reagents (ref: G9702A, G9496A) and protocols (ref: G9702-9000).

Reads were mapped on the GRCh38 genome with HiCUP v0.7.4 (27) and bowtie2 v2.3.2 (Statistics in **Supplementary Table 2**). Significant chromatin interactions were identified using CHiCAGO v1.13.1 (28) using a threshold of CHiCAGO score > 5 in different conditions; cell type: CD4^+^ T cells (n=15) and CD14^+^ monocytes (n=15); cell type and disease state: CD4^+^ T cells from SSc patients (n=10) and healthy controls (n=5), and CD14^+^ monocytes from SSc patients (n=10) and healthy controls (n=5). Principal component analysis (PCA) was performed in each cell type in order to detect potential biases (**Supplementary Figure 1**).

Chicdiff v0.6 (29) was used to detect differential interactions between different conditions: CD4^+^ T cells vs CD14^+^ monocytes; SSc patients vs healthy controls CD4^+^ T cells; and SSc patients vs healthy controls CD14^+^ monocytes. For each comparison, only those interactions with CHiCAGO score > 5 in at least one condition were included in differential analysis. Differential interactions with a weighted adjusted p-value < 0.05 were identified as significant. Spearman’s rank-order correlation was performed to test the correlation of log_2_ fold change values in differential interactions between patients and controls in CD4^+^ T cells and CD14^+^ monocytes.

### Genotype calling and allele specific analysis

Genotypes were called using GLIMPSE v1.1.1 (30) from Capture Hi-C reads aligned using HICUP v0.7.4 (27) to a masked GRCh38 genome. Genotype phasing was then carried out using the integrated phasing pipeline (31) which integrates population phasing with reads phasing using the Capture Hi-C reads. Reads were then split using SNPsplit v0.5.0 (32) generating allele specific alignments. Counts were for each allele for each CHiCAGO significant loop were then calculated using bedtools v2.30 (33) and data was integrated in Python 3.9. Allelic imbalance of the reads was tested using a binomial test for each sample which has heterozygous for that SNP and satisfied some requirements (see supplementary methods). All the resulting p-values were then checked for directionality and merged using the Firsher’s method for meta-analysis. Resulting p-values were then corrected using Benjamini-Hochberg method.

### RNA-seq library generation and processing

RNA was isolated from 0.5 million purified cells using the RNeasy microkit (QIAGEN, ref: 74004). Libraries were generated using Illumina Truseq Stranded Total RNA reagents and protocol, except for Control 1 CD4^+^ and CD14^+^ samples, for which library preparation failed. Reads were mapped using STAR v2.7.3a on the GRCh38 genome with GENCODE annotation v32. Reads were de-duplicated and then counted (**Supplementary Table 3**). Final count matrices were analyzed using edgeR v3.28.1 to perform normalization and differential expression analysis. Differentially expressed genes were called with an adjusted p-value of 0.1 (FDR 10%). Functional enrichment analyses were performed with g:Profiler (34) with default settings.

### Linking differential expression and differential interactions in CD4^+^ T cells vs CD14^+^ monocytes

Genes corresponding with the promoter end of significant differential interactions observed between CD4^+^ and CD14^+^ cells were overlapped with those differentially expressed. One-sided Fisher’s exact test was performed in order to calculate the enrichment of genes with differential interactions in those differentially expressed. In this set of overlapping genes, Spearman’s rank-order correlation was performed to test the correlation of log_2_ fold change values in differential interactions and differential expression. Finally, in order to test the distribution of log_2_ fold change values, a binomial exact test was performed on a subset of overlapping genes obtained adding a more stringent cutoff (absolute value of median log_2_FC > 2 for each gene). Functional enrichment analyses were performed with g:Profiler (34) with default settings.

### Defining SSc GWAS loci

All independent non-MHC disease-associated signals for SSc were selected from the largest meta-GWAS performed to date (9). We defined 23 regions based on linkage disequilibrium data and SNP proximity from the total of 27 independent signals described in GWAS. The window ranges and total number of SNPs in each of the 23 final loci are specified in **Supplementary Table S4**.

### Identifying eQTL genes for SSc GWAS loci

Publicly available eQTL databases were downloaded from the DICE eQTL database (35) of isolated immune cells and the eQTLgen (second release) database (36) of blood eQTLs. Genes were linked to each GWAS loci if the lead SNP was linked to a gene in the respective databases.

### Defining enhancers and TADs in CD4^+^ T cells and CD14^+^ monocytes

In order to define enhancer regions, chromHMM v1.22 annotations from 9 CD4^+^ T cells and 4 CD14^+^ monocytes were downloaded from the epiMAP project (37). For each cell type, enhancer regions were defined as those with state number from chromHMM corresponding to enhancer activity present in at least one sample. TADs definitions for CD4^+^ T cells and CD14^+^ monocytes were obtained from Javierre et al. (22).

### Overlap between pCHi-C, SSc GWAS loci, and enhancer regions

In order to prioritize certain interactions observed in pCHi-C data of particular interest in SSc GWAS loci, the SNP set previously defined in “Defining SSc GWAS loci” was overlapped with enhancer regions of each cell type (**Supplementary Table 4**). This new SNP set was then overlapped with the promoter interacting regions (PIRs) of significant pCHi-C interactions, defining candidate interacting genes as those in which their PIR overlaps with our significant SSc SNP set and enhancer regions. Garfield v2 (38) was used to estimate the enrichment of the GWAS SNPs in CD4^+^ T cells and CD14^+^ monocytes enhancer regions using a p-value threshold of 1×10^−8^. Functional enrichment analyses were performed for the sets of interacting genes observed in CD4^+^ T cells and CD14^+^ monocytes with g:Profiler (34).

### Drug target analysis

In order to assess if genes interacting with SSc GWAS loci in CD4^+^ T cells and CD14^+^ monocytes presented potential drug targets that could be repurposed for its use in SSc, those genes interacting with PIR overlapping significant SSc GWAS SNPs and enhancer regions, were used to model a protein-protein interaction (PPI) network using STRING v11 (39) (**Supplementary Table 5**). Protein products from these genes and those in direct PPI with them were used to query the OpenTargets Platform for drug targets. Additionally, the same platform and the Drugbank database were searched for information on clinical studies of drug targets of interest in SSc.

## RESULTS

In this study we generated pCHi-C data for CD4^+^ T cells and CD14^+^ monocytes from 10 SSc patients and 5 healthy controls. CHiCAGO was used to identify significant interactions (CHiCAGO score>5) for each cell type and disease condition (**Supplementary Table 6**) and Chicdiff was used to identify differential interactions between cell types, and between disease conditions for each cell type. A total of 81,624 and 74,853 significant interactions originating from 8,193 and 7,024 captured promoters were identified in CD4^+^ T cells and CD14^+^ monocytes, respectively. Through integration with published ChIP-seq data, we found that PIRs were enriched in H3K27ac and H3K4me3 histone marks from primary CD4^+^ naive T cells and CD14^+^ monocytes (**Supplementary Figure 2**), suggesting that promoters are more likely to interact with active regulatory regions such as enhancers.

### Differential interactions and expression between SSc patients and healthy controls

We first attempted to identify specific interactions that could be present in SSc patients but not in healthy controls, or vice versa, and thus, identify specific genes interacting with enhancer regions and SSc GWAS loci that could be of interest in SSc pathology. We identified a total of 4,858 significant differential interactions (weighted adjusted p-value<0.05) between SSc patients and healthy controls in CD4^+^ T cells, originating from 1,526 captured promoters, although the significance was modest (median weighted adjusted p-value=2.2×10^−2^) as compared with differential interactions between cell types (median weighted adjusted p-value=2.16×10^−10^). Moreover, we could not detect any significant differential interactions in CD14^+^ monocytes, indicating weak differences in cells isolated from blood between patients and controls. None of the 23 SSc GWAS associated regions showed significant differences at the interaction level between patients and controls. Regarding the transcriptomic differences, we identified a total of 62 and 63 differentially expressed genes (FDR < 0.10) between patients and controls in CD4^+^ T cells and CD14^+^ monocytes, respectively (**Supplementary Tables 7** and **8**). In the case of CD4^+^ T cells we observed significant enrichment in pathways related with immune response such as “positive regulation of immune system process” or “leukocyte activation” (**Supplementary Table 9**). However, we could not identify any functional enrichment regarding the 63 genes differentially expressed in CD14^+^ monocytes. Taken together, these results indicate that only modest differences are present in cells isolated from peripheral blood from patients.

### Identification of allele associated chromatin interactions

Recently there have been reports of allele associated chromatin interactions, these studies however were limited in size or had low resolution and based in cell lines (25,26). We wanted to test if allele associated interactions were present in our dataset of primary cells. To do this, for each individual, we assigned reads based on the haplotype of origin and quantified allelic imbalance in the read counts for all the CHiCAGO significant loops. We then aggregate the results for all the samples which were heterozygous for that SNP. After stringent quality control we identify 577 and 541 SNP-loop pairs in CD4^+^ T cells and CD14^+^ monocytes respectively (FDR<0.05) (**supplementary file 1 and 2**), representing 171 and 139 allele associated loops in CD4^+^ T cells and CD14^+^ monocytes respectively. None of the SSc GWAS SNPs displayed allelic imbalance interactions at this statistical power, however we still identify allele associated interactions with important genes related to immunity. For example, in CD4^+^ T cells we find allele associated interactions connecting the SNP rs661849 (located downstream of *IRF6*) and *TRAF3IP3* and *IRF6*. Interestingly this SNP is also an eQTL for these two genes, further validating our approach. In CD14^+^ monocytes we identify allele associated interactions linking a group of SNPs located around the promoter of *GPX3* and the promoters of *GPX3* and *TNIP1*. Again, these SNPs were also identified as eQTLs for both genes, although these specific SNPs were not found to be associated with any disease in the GWAS catalog. Overall, we identify allele associated interactions between SNPs and 62 and 57 genes in CD4^+^ T cells and CD14^+^ monocytes respectively. Of these 38 and 31 respectively were also eQTL for those genes in the eQTLgen database (4 and 3 respectively in the DICE eQTL database).

### Linking differential expression and differential interactions in CD4^+^ T cells vs CD14^+^ monocytes

Next, we decided to look at differences at the interaction and expression level between cell types and how these correlate with each other, without taking disease state into account. We identify 2,257 strongly differentially expressed genes (FDR<0.05, |log_2_FC|>2) between CD4^+^ T cells and CD14^+^ monocytes, of which 919 and 1,338 genes were overexpressed in CD4^+^ T cells and CD14^+^ monocytes, respectively. Overrepresentation analyses showed that each group of genes is, as expected, significantly enriched in terms related with T cells and monocytes specific pathways, including gene ontology terms such as “T cell activation” and “T cell differentiation” in CD4^+^ T cells, and “leukocyte activation” in CD14^+^ monocytes (**Supplementary Tables 10** and **11**). Regarding the interactome, we identify 71,213 significant differential interactions (weighted adjusted p-value<0.05) originating from 8,223 captured promoters. We observed that differentially expressed genes are in fact significantly enriched in differentially interacting genes (fisher exact test p-value=3.54×10^−37^, OR=1.77). Furthermore, from the total of 1,209 differentially expressed genes overlapping differentially interacting genes, we observed that genes overexpressed in a specific cell type significantly correlated with increased number of chromatin interactions in that cell type, and vice versa (Spearman’s rank correlation p-value=1.04×10^−197^, rho=0.73). Finally, we applied a more stringent cutoff in differentially interacting genes (|log_2_FC|>2), leading to a total of 97 differentially expressed genes overlapping differential interactions. In this subset, only 2 of the 97 genes did not behave as expected; 23 and 72 genes were overexpressed and presented an increased number of interactions in CD4^+^ T cells and CD14^+^ monocytes, respectively (exact binomial test p-value=6.01×10^−26^, probability of success=98%) (**Supplementary Figure 3**). Thus, our results show that chromatin conformation is highly cell type specific and linked to gene expression and demonstrate the importance of using the correct cell types to define promoter interactions and linking genes to GWAS loci.

### SSc GWAS loci and CD4^+^ / CD14^+^ promoter interactions

Finally, we identify new potential target genes associated to SSc GWAS loci, as well as the potential implication of different cell types in those associations. We performed a multi-omic approach overlapping 23 regions defined based on the most powerful SSc meta-GWAS performed to date (9) with enhancer regions and our pCHi-C data (see online **Supplementary Material and Methods** for more detail). In addition we have also integrated our findings with two large eQTL databases, eQTLgen, which is the largest blood eQTL meta-analysis (36), and DICE eQTL, which is the largest study that makes use of purified immune cell populations (35) (**supplementary file 3**). From the total of 1,505 SNPs genome-wide significant (p-value<5×10^−8^) SNPs associated with SSc and those in high linkage disequilibrium (r^2^>0.8) with them, 445 (29.6%) and 284 (18.9%) overlapped with enhancer regions from CD4^+^ T cells and CD14^+^ monocytes, respectively. As expected, the GWAS SNPs significantly enriched enhancer regions in both CD4^+^ and CD14^+^ (Garfield enrichment test OR=3.40, p-value 6.7×10^−4^ in CD4^+^ T cells; OR=3.05, p-value 1.7×10^−3^ in CD14^+^ monocytes). In addition, the differences in the number of SNPs overlapping CD4^+^ and CD14^+^ enhancer regions was significant (two proportion z-test p-value=0.001), showing a stronger enrichment with CD4^+^ T cell enhancer regions as compared with that observed in CD14^+^ monocytes. These GWAS SNPs within enhancer regions were overlapped with PIRs from pCHi-C, resulting in a total of 398 and 109 significant interactions in CD4^+^ T cells and CD14^+^ monocytes, respectively (**Supplementary Table 4**). The promoter ends of those interactions correspond to 46 genes, with a total of 40 and 27 interacting genes in CD4^+^ T cells and CD14^+^ monocytes, respectively (**Table 1**).

**Table 1.** Promoter capture Hi-C target genes for the 23 systemic sclerosis associated regions in CD4^+^ T cells and CD14^+^ monocytes.

| Chr | Bp (start - end)<sup>1</sup> | GWAS Locus<sup>2</sup> | pChI-C target genes<sup>3</sup> |  | CD4<sup>+</sup> vs CD14<sup>+</sup> |  |
| --- | --- | --- | --- | --- | --- | --- |
|  |  |  | CD4<sup>+</sup> T cells | CD14<sup>+</sup> monocytes | Differential interactions | Differential expression |
| 1 | 67326053 - 67448804 | rs3790566 (*IL12RB2*) |  |  |  |  |
| 1 | 167445635 - 167465040 | rs2056626 (*CD247*) | **CD247, CREG1** |  | **CD247, CREG1** | **CD247, CREG1** |
| 1 | 173337507 - 173391947 | rs1857066 (*TNFSF4-LOC100506023 - PRDX6*) |  |  |  |  |
| 2 | 190642047 - 190698201 | rs16832798 (*NAB1*) | *MFSD6, NEMP2* | *MFSD6, NEMP2, HIBCH;INPP1* | *MFSD6, NEMP2, HIBCH;INPP1* | *MFSD6, NEMP2, HIBCH, INPP1* |
| 2 | 191035723 - 191108308 | rs3821236 (*STAT4*) | **STAT4, NABP1** |  | **STAT4, NABP1** | **STAT4, NABP1** |
| 3 | 58084620 - 58482701 | rs4076852 (*FLNB -DNASE1L3-PXK*) | *RPP14, KCTD6* | *RPP14, KCTD6* | *RPP14, KCTD6* | *KCTD6* |
| 3 | 119384733 - 119546340 | rs9884090 (*POGLUT1-TIMMDC1-CD80- ARHGAP31*) | **TMEM39A;POGLUT1** |  | **TMEM39A;POGLUT1** | **TMEM39A, POGLUT1** |
| 3 | 160002484 - 160030580 | rs589446 (*IL12A*) |  | *SMC4;IFT80* | *SMC4;IFT80* | *SMC4, IFT80* |
| 4 | 960523 - 990021 | rs11724804 (*DGKQ*) | *GAK;TMEM175, FGFR1* | *GAK;TMEM175, FGFR1* | *FGFR1* | *GAK, TMEM175 FGFR1* |
| 4 | 102477892 - 102615256 | rs230534 (*NFKB1*) | *SLC39A8, NFKB1, UBE2D3;CISD2, SLC9B1, BDH2* | *SLC39A8, UBE2D3;CISD2, BDH2* | *SLC39A8, NFKB1, UBE2D3;CISD2, SLC9B1, BDH2* | *SLC39A8, UBE2D3, CISD2, BDH2* |
| 5 | 151064651 - 151080486 | rs3792783 (*TNIP1*) |  |  |  |  |
| 6 | 106181815 - 106339294 | rs633724 (*ATG5*) |  |  |  |  |
| 7 | 128933913 - 129095960 | rs36073657 (*IRF5-TNPO3*) |  |  |  |  |
| 8 | 11474517 - 11544554 | rs2736340 (*FAM167A-BLK*) |  |  |  |  |
| 8 | 60638547 - 60664239 | rs685985 (*RAB2A-CHD7*) | **ASPH, SDCBP, CHD7** |  | **ASPH, SDCBP** | **ASPH, SDCBP, CHD7** |
| 11 | 554659 - 619789 | rs6598008 (*CDHR5 -IRF7*) |  |  |  |  |
| 11 | 2311894 - 2363262 | rs2651804 (*TSPAN32, CD81-AS1*) | **TSSC4** |  | **TSSC4** | **TSSC4** |
| 11 | 118704617 - 118875175 | rs11217020 (*DDX6*) | *CXCR5, UPK2, DDX6, IFT46;ARCN1* | *CXCR5* | *CXCR5, UPK2, DDX6* | *CXCR5, DDX6, ARCN1* |
| 15 | 74739180 - 75148328 | rs1378942 (*CSK*) | *CSK, CLK3, ULK3, SCAMP2, MPI, FAM219B, COX5A* | *CSK, CLK3, ULK3, SCAMP2, MPI, FAM219B, COX5A, C15orf39* | *CSK, ULK3, SCAMP2, MPI, FAM219B, COX5A, C15orf39* | *CSK, CLK3, ULK3, SCAMP2, MPI, FAM219B, COX5A, C15orf39* |
| 16 | 85932852 - 85979945 | rs11117420 (*IRF8*) |  | **IRF8** | **IRF8** | **IRF8** |
| 17 | 39747478 - 39933464 | rs883770 (*IKZF3-GSDMB*) | **IKZF3, ERBB2, PSMD3** | *ERBB2* | **IKZF3, ERBB2, PSMD3** | **IKZF3, ERBB2, PSMD3** |
| 17 | 75193533 - 75279345 | rs1005714 (*NUP85-GRB2*) |  |  |  |  |
| 19 | 18068862 - 18093031 | rs2305743 (*IL12RB1*) | *PIK3R2, RAB3A* | *RAB3A* | *PIK3R2, RAB3A* |  |

Genes classically associated with systemic sclerosis through proximity to GWAS loci are highlighted in bold.
<sup>1</sup>Bp in GRCh38 (hg 38) assembly
<sup>2</sup>Locus as defined by López-Isac et al.(9)
<sup>3</sup>Genes corresponding with promoter interacting regions overlapping enhancer regions and systemic sclerosis GWAS SNPs
*Bp* base pair, *Chr* chromosome, *GWAS* Genome-wide association studies, *N* number, *pChI-C* promoter capture Hi-C, *SNP* single-nucleotide polymorphism.

The physical interaction maps presented here identify 39 new potential candidate genes and confirm 7 genes which have been previously associated to SSc using genomic proximity. Differential expression and differential interaction data for each of the 46 genes and baited promoters are available in **Supplementary Tables 12** and **13**, respectively. Interestingly, some SSc confirmed genes such as *IRF8, STAT4*, or *CD247*, showed cell type specific interactions (**Figures 1-3**). The rs11117420 (*IRF8*) locus (**Figure 1**) provides a good example in which interactions between SNPs overlapping enhancer regions (represented by H3K27ac mark peaks) and the *IRF8* promoter are found exclusively in one cell type and is associated to differential gene expression between cells, in which CD14^+^ monocytes showed a much higher expression of *IRF8* (log_2_FC=-4.47, FDR=3.11×10^−72^). In the case of the rs3821236 (*STAT4*) locus (**Figure 2**), significant interactions with the *STAT4* promoter were identified exclusively in CD4^+^ T cells, corresponding with a TAD specific for CD4^+^ T cells that is not found in monocytes. In addition, *STAT4* showed a significantly higher expression in CD4^+^ T cells as compared with CD14^+^ monocytes (log_2_FC=7.05, FDR=1×10^−304^). Interestingly, neither of these two loci showed eQTL signals to these genes in either database. rs11117420 was eQTL for a lncRNA gene (*RP11-542M13*.*3*) whilst rs3821236 only showed weak eQTL signals for *GLS* and *MFSD6*. Cell type specific interactions are also observed in the rs2056626 (*CD247*) GWAS locus (**Figure 3**), in which significant interactions between SNPs and the *CD247* promoter are identified only in CD4^+^ T cells, with an increased expression of this gene in CD4^+^ T cells as compared with CD14^+^ monocytes (log_2_FC=7.49, FDR=3.99×10^−210^). In this case we found that rs2056626 is also a strong eQTL for *CD247*.

**Figure 1.**
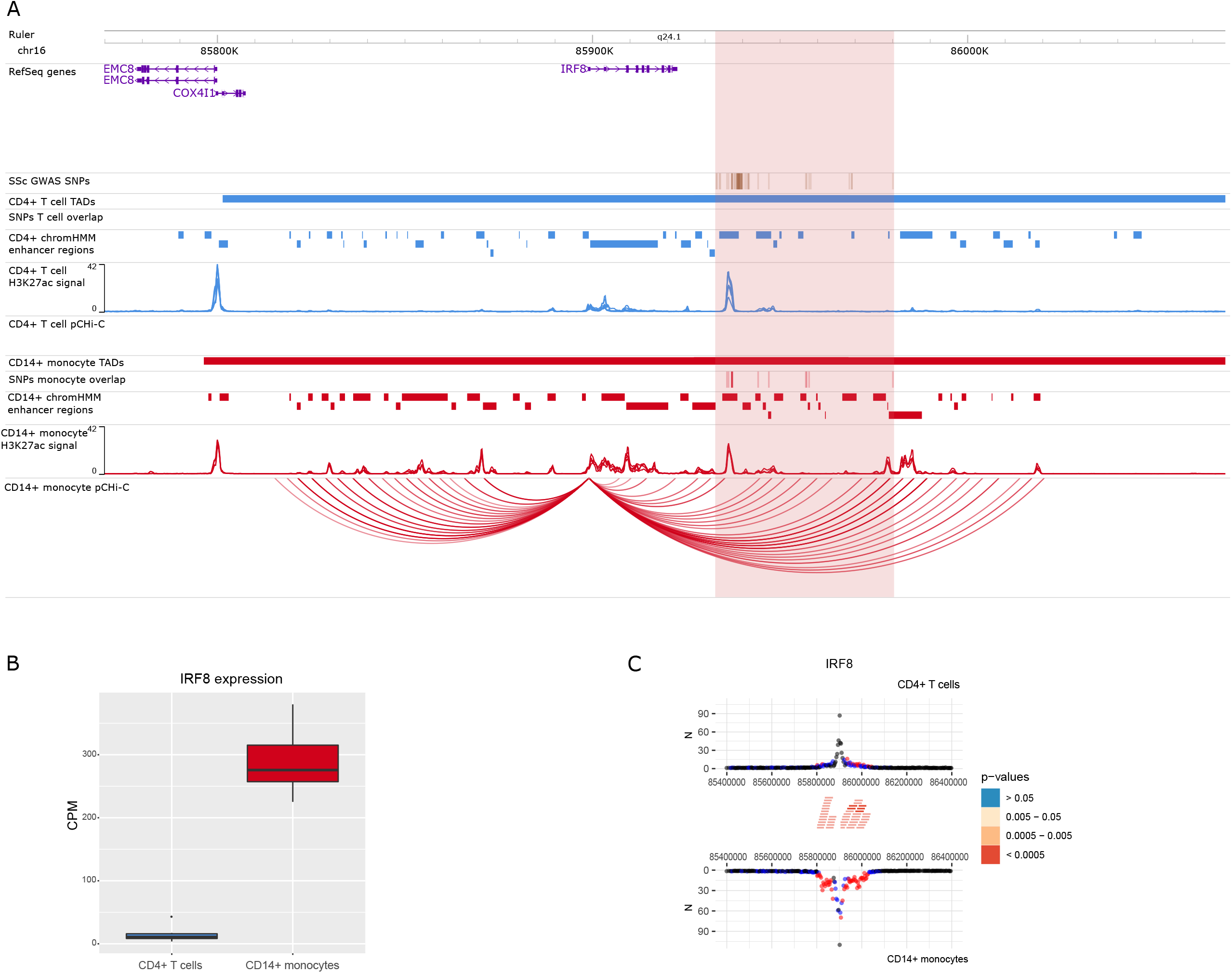
Promoter capture Hi-C (pCHi-C) interactions and gene expression in the rs11117420 (*IRF8*) GWAS locus. (**A**) Genomic coordinates (GRCh38) are shown at the top of the panel. The tracks include RefSeq genes (NCBI), systemic sclerosis (SSc) GWAS SNPs from López-Isac et al.(9) and those in high linkage disequilibrium (LD) (r^2^>0.8), TADs (shown as bars), SNPs overlapping promoter interacting regions (PIRs) and enhancer regions, enhancer regions as defined by chromHMM, H3K27ac signal, and pCHi-C significant interactions (CHiCAGO score > 5) (shown as arcs) in CD4^+^ T cells (blue) and CD14^+^ monocytes (red). The highlighted region in red includes all the systemic sclerosis (SSc) SNPs LD block. (**B**) Gene expression level of *IRF8* from CD4^+^ T cells and CD14^+^ monocytes in counts per million (CPM). (**C**) Chicdiff bait profiles were generated for *IRF8* gene. The plot shows the raw read counts versus linear distance from the bait fragment as mirror images for CD4^+^ T cells and CD14^+^ monocytes. Other-end interacting fragments are pooled and color-coded by their adjusted weighted p-value. Significant differentially interacting regions detected by Chicdiff overlapping SSc GWAS SNPs and enhancer regions are depicted as red blocks.

**Figure 2.**
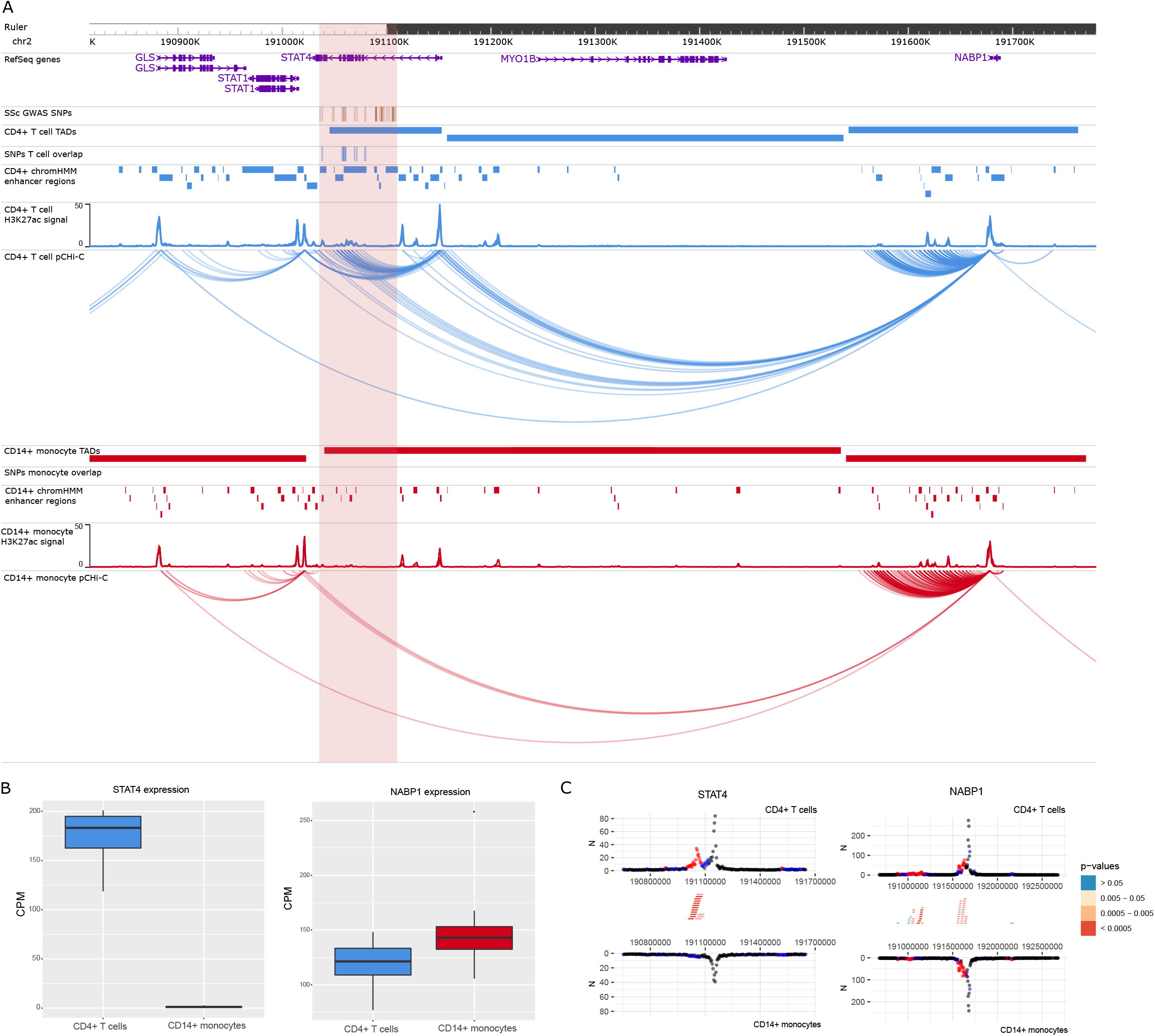
Promoter capture Hi-C (pCHi-C) interactions and gene expression in the rs11117420 (*STAT4*) GWAS locus. (**A**) Genomic coordinates (GRCh38) are shown at the top of the panel. The tracks include RefSeq genes (NCBI), systemic sclerosis (SSc) GWAS SNPs from López-Isac et al.(9) and those in high linkage disequilibrium (LD) (r^2^>0.8), TADs (shown as bars), SNPs overlapping promoter interacting regions (PIRs) and enhancer regions, enhancer regions as defined by chromHMM, H3K27ac signal, and pCHi-C significant interactions (CHiCAGO score > 5) (shown as arcs) in CD4^+^ T cells (blue) and CD14^+^ monocytes (red). The highlighted region in red includes all the systemic sclerosis (SSc) SNPs LD block. (**B**) Gene expression level of *STAT4* and *NABP1* from CD4^+^ T cells and CD14^+^ monocytes in counts per million (CPM). (**C**) Chicdiff bait profiles were generated for *STAT4* and *NABP1* genes. Plots show the raw read counts versus linear distance from the bait fragment as mirror images for CD4^+^ T cells and CD14^+^ monocytes. Other-end interacting fragments are pooled and color-coded by their adjusted weighted p-value. Significant differentially interacting regions detected by Chicdiff overlapping SSc GWAS SNPs and enhancer regions are depicted as red blocks.

**Figure 3.**
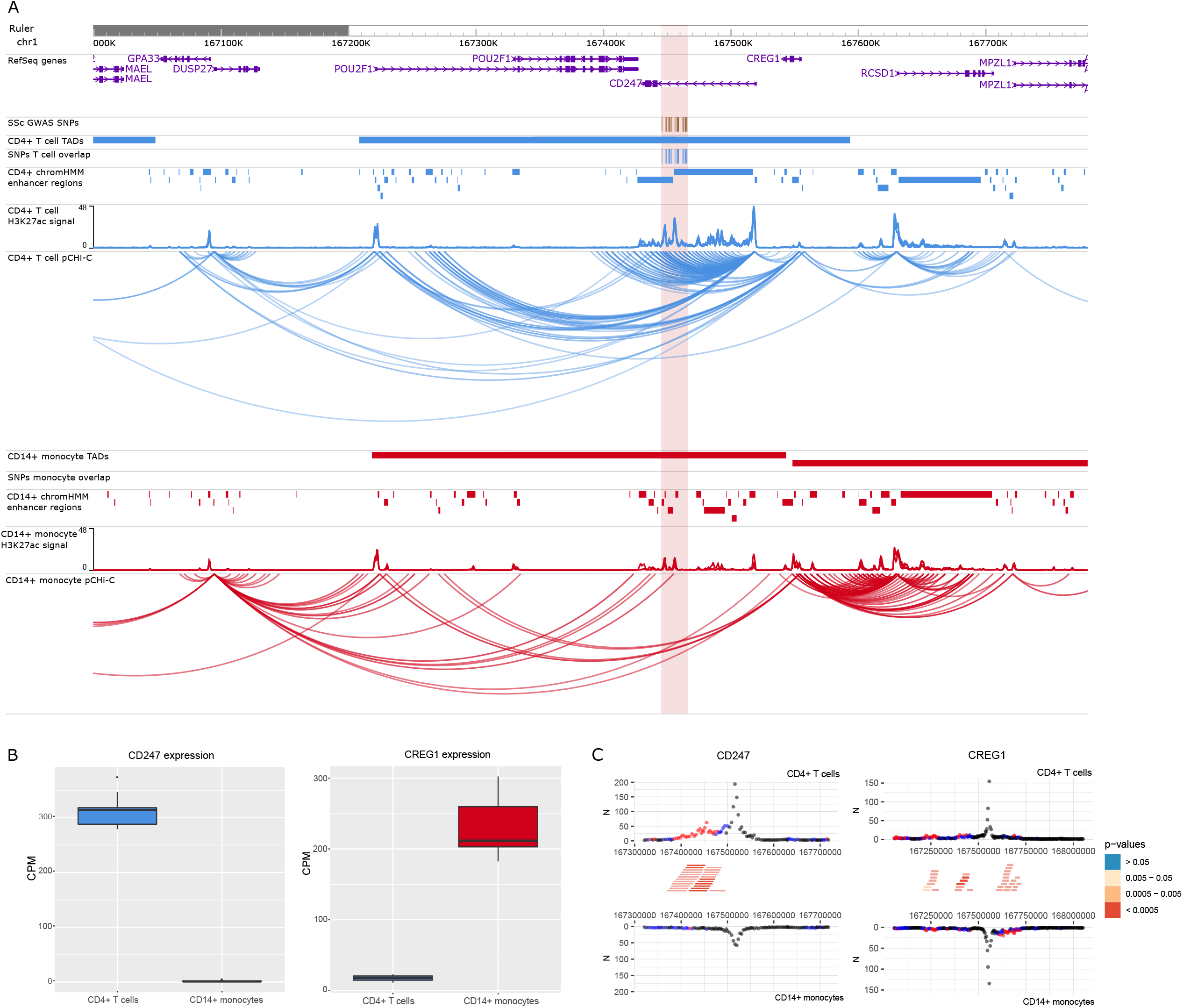
Promoter capture Hi-C (pCHi-C) interactions and gene expression in the rs2056626 (*CD247*) GWAS locus. (**A**) Genomic coordinates (GRCh38) are shown at the top of the panel. The tracks include RefSeq genes (NCBI), systemic sclerosis (SSc) GWAS SNPs from López-Isac et al.(9) and those in high linkage disequilibrium (LD) (r^2^>0.8), TADs (shown as bars), SNPs overlapping promoter interacting regions (PIRs) and enhancer regions, H3K27ac signal, enhancer regions as defined by chromHMM, and pCHi-C significant interactions (CHiCAGO score > 5) (shown as arcs) in CD4^+^ T cells (blue) and CD14^+^ monocytes (red). The highlighted region in red includes all the systemic sclerosis (SSc) SNPs LD block. (**B**) Gene expression level of *CD247* and *CREG1* from CD4^+^ T cells and CD14^+^ monocytes in counts per million (CPM). (**C**) Chicdiff bait profiles were generated for *CD247* and *CREG1* genes. Plots show the raw read counts versus linear distance from the bait fragment as mirror images for CD4^+^ T cells and CD14^+^ monocytes. Other-end interacting fragments are pooled and color-coded by their adjusted weighted p-value. Significant differentially interacting regions detected by Chicdiff overlapping SSc GWAS SNPs and enhancer regions are depicted as red blocks.

On the other hand, we identified new potential candidate genes interacting with SSc GWAS associated SNPs, for example in the rs11217020 (*DDX6*) locus (**Figure 4**) we found significant interactions between SNPs overlapping enhancer regions and not only *DDX6*, but also other potential candidate genes including *CXCR5, UPK2*, and *IFT46/ARCN1* promoters in CD4^+^ T cells. In CD14^+^ monocytes, only a significant interaction with *CXCR5* promoter was found. All of these interactions are intra-TAD, except for the one including *IFT46/ARCN1* promoters, and we observed a significantly higher gene expression of *CXCR5* (log_2_FC=3.21, FDR=1.05×10^−09^) and *DDX6* (log_2_FC=1.14, FDR=2.38×10^−83^) in CD4^+^ T cells, while *ARCN1* showed a slight overexpression in CD14^+^ monocytes (log_2_FC=-0.21, FDR=3.69×10^−03^). *ARCN1* and *CXCR5* are also eQTL hits for this SNP in blood (Supplementary Table), further supporting their role in SSc pathogenesis.

**Figure 4.**
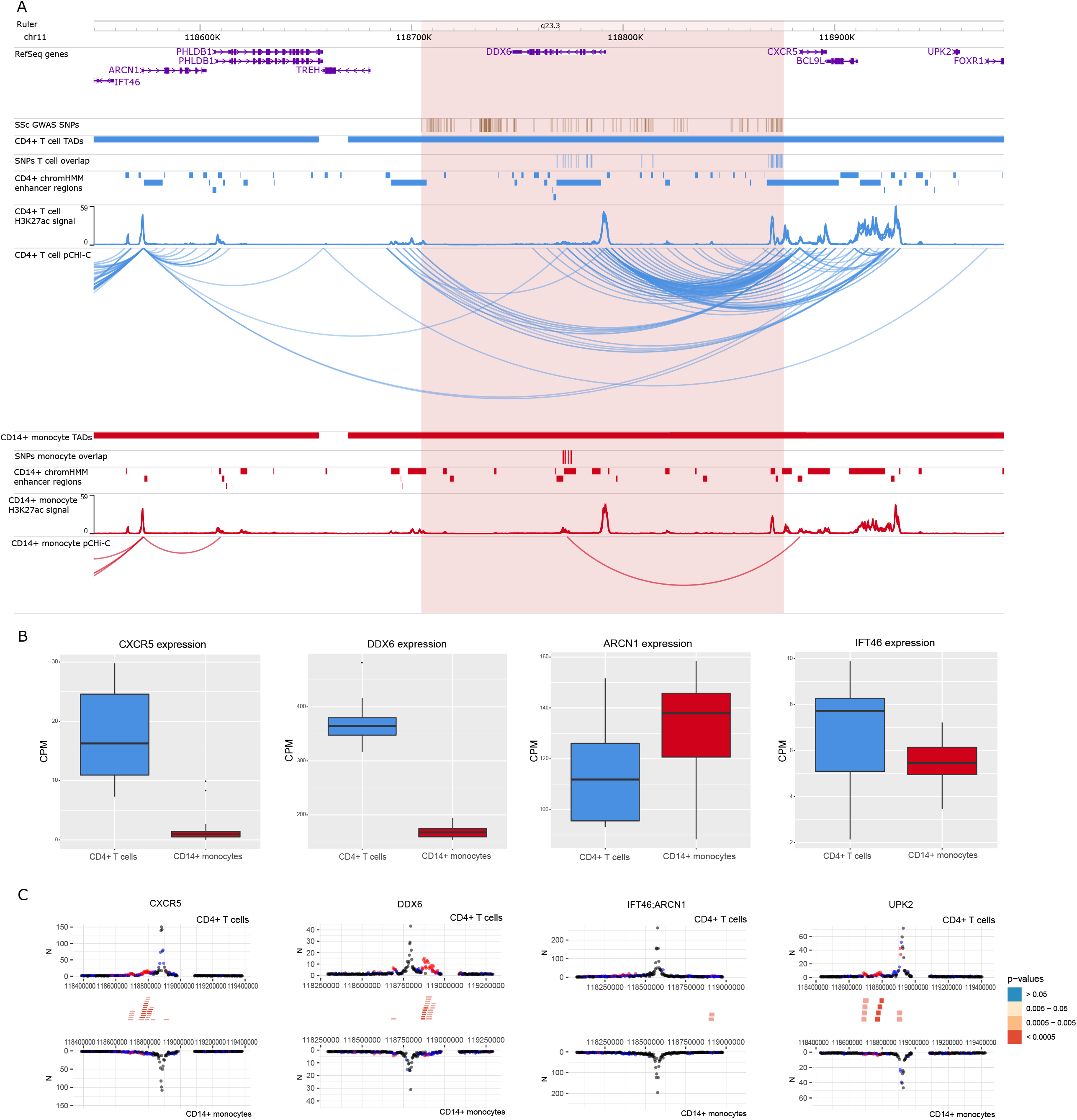
Promoter capture Hi-C (pCHi-C) interactions and gene expression in the rs11217020 (*DDX6*) GWAS locus. (**A**) Genomic coordinates (GRCh38) are shown at the top of the panel. The tracks include RefSeq genes (NCBI), systemic sclerosis (SSc) GWAS SNPs from López-Isac et al.(9) and those in high linkage disequilibrium (LD) (r^2^>0.8), TADs (shown as bars), SNPs overlapping promoter interacting regions (PIRs) and enhancer regions, enhancer regions as defined by chromHMM, H3K27ac signal, and pCHi-C significant interactions (CHiCAGO score > 5) (shown as arcs) in CD4^+^ T cells (blue) and CD14^+^ monocytes (red). The highlighted region in red includes all the systemic sclerosis (SSc) SNPs LD block. (**B**) Gene expression level of *CXCR5, DDX6, ARCN1*, and *IFT46* from CD4^+^ T cells and CD14^+^ monocytes in counts per million (CPM). (**C**) Chicdiff bait profiles were generated for *CXCR5, DDX6, IFT46/ARCN1* (shared capture bait), and *UPK2* genes. Plots show the raw read counts versus linear distance from the bait fragment as mirror images for CD4^+^ T cells and CD14^+^ monocytes. Other-end interacting fragments are pooled and color-coded by their adjusted weighted p-value. Significant differentially interacting regions detected by Chicdiff overlapping SSc GWAS SNPs and enhancer regions are depicted as red blocks.

In order to identify what pathways could be driving disease in the two different cell types, we performed a functional enrichment analysis including the genes interacting with SSc GWAS loci for each cell type. In CD4^+^ T cells, the set of 40 interacting genes observed showed enrichment in virus response and pancreatic carcinoma (**Supplementary Table 14**). In accordance with this, a higher incidence of cancer in SSc patients compared with the general population has been suggested in several studies (40). On the other hand, the set of 27 interacting genes observed in CD14^+^ monocytes showed enrichment in tyrosine kinase activity (**Supplementary Table 15**), which plays an important role in fibrosis, and has been related with SSc pathogenesis, being tyrosine kinase inhibitors one of the most promising antifibrotic therapies for SSc and other fibrotic diseases (41).

Plots of the interactions for the rest of the SSc GWAS loci in CD4^+^ T cells and CD14^+^ monocytes can be found in **Supplementary Figures 4-22**. All eQTL hits are available in **Supplementary File 3**.

## DISCUSSION

Our investigation integrates four dimensions in the study of SSc genetics; GWAS, chromosome conformation, gene expression, and cell-specificity. In this regard, our findings stress the importance of using the correct cell type in the functional interpretation of GWAS associations. We identified new target genes, and confirmed others, in SSc GWAS loci in two of the main cell types associated with the disease, CD4^+^ T cells and CD14^+^ monocytes. We also further validate the presence of interactions that are altered by common genomic variations and show how these are correlated with eQTLs.

One of the new candidate genes observed in pCHi-C data corresponds to CXC chemokine receptor type 5 (*CXCR5*) in the *DDX6* GWAS locus (**Figure 4**). *CXCR5* plays an important role in the differentiation of follicular helper T (Tfh) cells, and is highly expressed in CD4^+^ and CD8^+^ T cells (42). In addition, a recently published study observed that Tfh cells (CD4^+^CXCR5^+^PD-1^+^) are increased in systemic sclerosis, and correlate with SSc severity (43). In line with the above, interactions with the promoter of this gene were identified specifically in CD4^+^ T cells in our study, and the gene’s expression was specific to this cell type. Furthermore, *CXCR5* has been associated through GWAS studies in other similar immune-mediated diseases, such as rheumatoid arthritis or inflammatory bowel disease (44,45). Thus, *CXCR5* represents a good candidate gene contributing to SSc pathology, with a particular interest in CD4^+^ T cells.

Another interesting example is found in the rs685985 (*RAB2A-CHD7*) locus, a recently discovered locus associated with SSc (9) (**Supplementary Figure 16**). Within this region, we observed significant interactions between SSc GWAS SNPs and the closest gene, *CHD7*, in CD4^+^ T cells. *CHD7* is a chromatin remodeler that has been associated with lymphocyte (and other immune-related cells) counts in blood through GWAS (46). Regarding the rs589446 (*IL12A*) locus (**Supplementary Figure 9**), we identified long-range interactions between SSc GWAS SNPs and the promoter of *SMC4* in CD14^+^ monocytes. *SMC* family genes play a central role in organizing and compacting chromosomes. In this line, a recent study showed *SMC4* promotes an inflammatory innate immune response, which is directly associated with monocyte activity, through enhancing NEMO transcription, an essential modulator of NF-Κb (47). Although *IL12A* has been traditionally set as the most probable candidate gene for this association, we did not observe any interactions between SSc GWAS SNPs and the promoter of this gene. Here, it is important to note the increased difficulty to identify significant short-range interactions (<50 kb) as background read count levels are dependent on the distance between fragments (28). This phenomenon represents a limitation in this kind of studies, as most of the GWAS SNPs are classically related with the closest gene, being these SNPs located within the gene itself in some cases. In this regard, newer high resolution Hi-C methods should help overcome the limitation of detecting very short range interactions (48).

Regarding previously confirmed genes associated with SSc, we described interactions between *IRF8* promoter and SSc GWAS SNPs that were only present in CD14^+^ monocytes (**Figure 1**), corresponding with an upregulated expression of this gene in CD14^+^ monocytes as compared with CD4^+^ T cells. This transcription factor plays an important role in differentiation and regulation of monocytes and macrophages (49). Furthermore, variants in *IRF8* have been associated with monocyte counts across different populations (50), and a downregulation of *IRF8* in monocytes and macrophages of SSc patients that may affect the fibrotic phenotype of the disease was reported (51). A recent study demonstrated that the deletion of an enhancer region corresponding with our SSc GWAS locus in mice model decreased *Irf8* expression, resulting in an overproduction of inflammatory Ly6c^+^ monocytes (52). Thus, our results confirm the association of *IRF8* with SSc through physical chromatin interactions particularly in CD14^+^ monocytes. Although, given the evidence, this locus is very likely to affect *IRF8*, we did not find evidence of eQTL signals in the two databases we have explored. This is likely due to limitations of eQTL studies which require very large datasets to identify signals, which are very laborious to apply to all possible cell populations (in this case Monocytes). A recent study has also identified limitations in the design of eQTL studies when used to assign genes to GWAS loci (53). In other loci, *CD247* and *STAT4* have been described in many previous GWAS studies as main candidate genes associated with SSc (9,10). In our study interactions were exclusively found in CD4^+^ T cells (**Figures 2-3**). These findings are in line with literature, as both genes play an important role particularly in T cell signaling and differentiation (54,55). Thus, our results highlight the importance of studying GWAS signals with the specific cell types in which interactions are found, acting as a starting point for follow-up functional studies that can relate these signals with the disease.

As a proof of concept, we wanted to determine the potential for these genes to indicate novel treatment options for SSc. From the 46 genes that present PIRs overlapping significant SSc GWAS SNPs and enhancer regions, we identified a total of 21 drugs with interest in SSc targeting protein products in strong protein-protein interaction (PPI) with 13 of those genes (5 of them specific for CD4^+^ T cells interactions) (**Table 2**). Fifteen of these drugs correspond to potential drug targets already in use, or at least in completed clinical phase III, in other similar immune-mediated diseases that could be repurposed for SSc treatment, such as metformin or dimethyl fumarate. Apart from new potential drug targets, tocilizumab and nintedanib were two of the drugs highlighted in our analysis, both of them approved by Food and Drug Administration (FDA) for its use in SSc-associated interstitial lung disease (56,57). We also identified 4 drugs which present advanced clinical trials developed in SSc (tofacitinib, bosentan, methylprednisolone and mycophenolic acid).

**Table 2.**
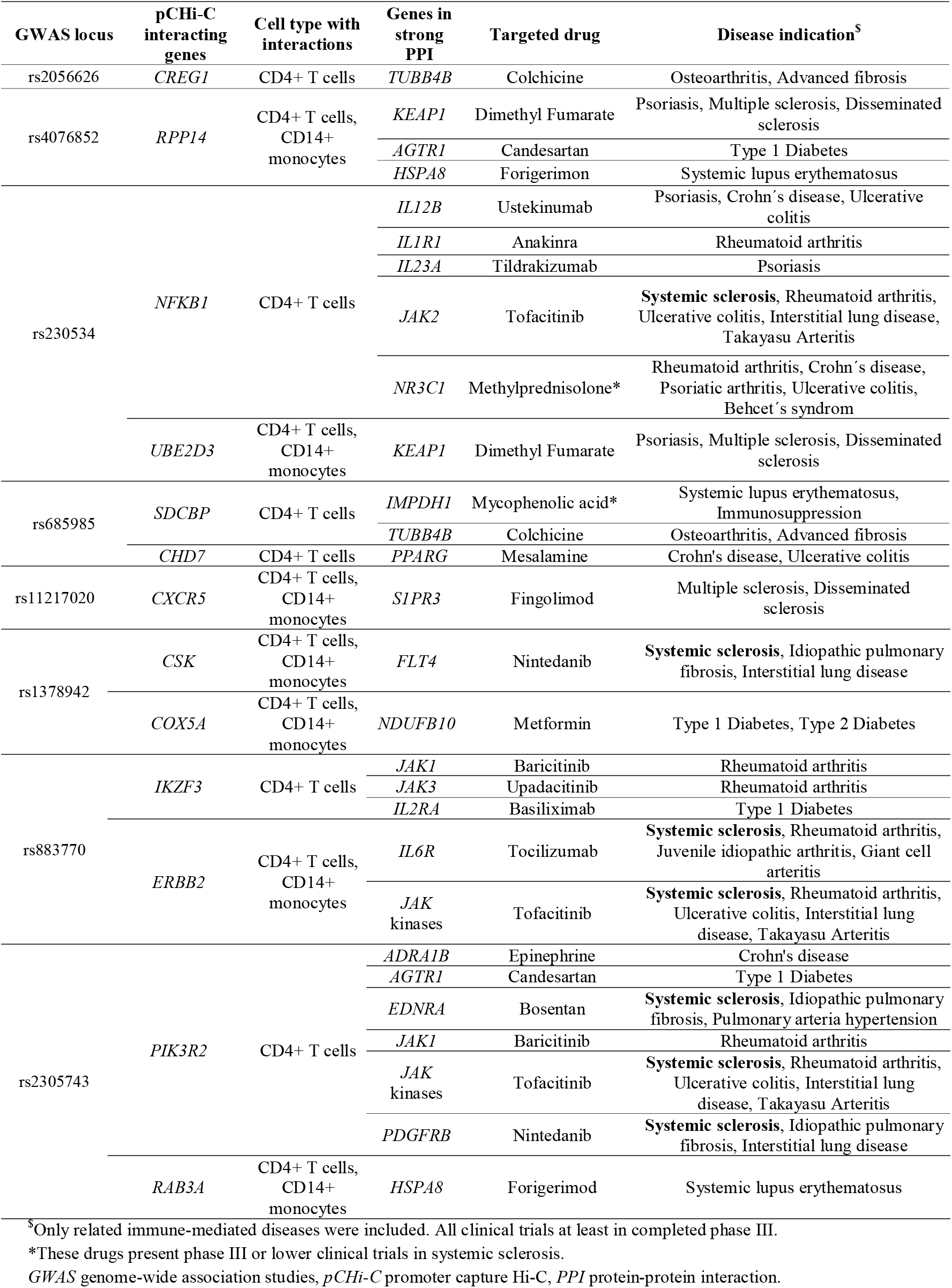
Summary of potential targets for drug repurposing in systemic sclerosis based on pCHi-C data.

Our results reveal that 3D chromatin structure is largely preserved between SSc patients and healthy controls at least in CD4^+^ T cells and CD14^+^ monocytes derived from peripheral blood. So far, there is only one published study in which authors attempt to observe differences at the interaction level between patients and healthy controls in CD4^+^ T cells from juvenile idiopathic arthritis patients (58). However, no differences at the interactome level were observed, which supports our hypothesis and underlines the difficulty to describe these subtle differences with current technology. Interestingly, it has been shown that subtle differences in chromatin interactions may be correlated with large functional effects on gene expression (25). More significant differences could be expected if cells were isolated from the site of active disease, and further studies involving these samples would be of great interest. On the other hand, we identify many significant allele-associated interactions and we are the first to show that this analysis is possible in primary cells isolated from patients. We show that many of the SNPs associated with these loops are also eQTLs for the genes they interact with. The overall number of loops presenting allelic imbalance is still very low compared to the loops tested (0.2%) but in line with previous studies (25,26). We think this is due to difficulties in assigning reads to a specific allele (only reads overlapping phased heterozygous sites can be tested) and the limited number of read pairs in long range interactions, consequently limiting statistical power.

Finally, we wanted to describe general differences between cell types at the interaction and expression level and how these are correlated. We observed that overexpressed genes in a specific cell type correlated with an increased number of interactions, and that those genes were enriched in specific pathways related with T cells and monocytes signaling, activation, and differentiation. These results demonstrate that interactions are directly related with the expression of important genes implicated in cell type specific pathways. In this regard, a recent study observed that disease-associated genes tend to be connected by cell-type-specific interactions (59). Thus, our data presented here will aid future studies to identify cell types enriched with interactions overlapping GWAS loci.

## Data Availability

The processed datasets generated from this study are available in the GEO repository under the accession number GSE212100.
Raw reads and genotypes can be provided upon reasonable request from the authors.

## ACKNOWLEDGEMENTS

We greatly acknowledge the expert technical assistance of Sofía Vargas Roldán at the Institute of Parasitology and Biomedicine López-Neyra [IPBLN]–CSIC, as well as the assistance given by Information Technology Services and the use of the Computational Shared Facility at The University of Manchester, United Kingdom. This work is part of the Doctoral Thesis “Deciphering the genomic architecture of systemic sclerosis”.

## DATA AVAILABILITY STATEMENT

The processed datasets generated from this study are available in the GEO repository under the accession number GSE212100.

Raw reads and genotypes can be provided upon reasonable request from the authors.

