## Supplementary materials, figures and tables for "Functional genomics in primary T cells and monocytes identifies mechanisms by which genetic susceptibility loci influence systemic sclerosis risk"

González-Serna D et al.

Corresponding to: David González-Serna and Gisela Orozco

**Table of contents**

**Material and Methods**	**3**

Supplementary Figures	9

1. Principal components from pCHi-C data for each sample in CD4^+^ T cells and CD14^+^ monocytes	9
2. Enrichment of features within promoter interacting regions of pCHi-C interactions	10
3. Distribution of the 97 differentially expressed genes overlapping differentially interacting genes in CD4^+^ T cells vs CD14^+^ monocytes comparison	11
4. Promoter capture Hi-C interactions in the *IL12RB2* locus.	12
5. Promoter capture Hi-C interactions in the *TNFSF4-LOC100506023-PRDX6* locus.	13
6. Promoter capture Hi-C interactions in the *NAB1* locus.	14
7. Promoter capture Hi-C interactions in the *FLNB-DNASE1L3-PXK* locus.	15
8. Promoter capture Hi-C interactions in the *POGLUT1-TIMMDC1-CD80-ARHGAP31*locus.	16
9. Promoter capture Hi-C interactions in the *IL12A* locus.	17
10. Promoter capture Hi-C interactions in the *DGKQ* locus.	18
11. Promoter capture Hi-C interactions in the *NFKB1* locus.	19
12. Promoter capture Hi-C interactions in the *TNIP1* locus.	20
13. Promoter capture Hi-C interactions in the *ATG5* locus.	21
14. Promoter capture Hi-C interactions in the *IRF5-TNPO3* locus.	22
15. Promoter capture Hi-C interactions in the *FAM167-BLK* locus.	23
16. Promoter capture Hi-C interactions in the *RAB2A-CHD7* locus.	24
17. Promoter capture Hi-C interactions in the *CDHR5 -IRF7* locus.	25
18. Promoter capture Hi-C interactions in the *TSPAN32,CD81-AS1* locus.	26
19. Promoter capture Hi-C interactions in the *CSK* locus.	27
20. Promoter capture Hi-C interactions in the *IKZF3-GSDMB* locus.	28
21. Promoter capture Hi-C interactions in the *NUP85-GRB2* locus.	29
22. Promoter capture Hi-C interactions in the *IL12RB1* locus.	30
23. Flow cytometry CD4+ T cells	31
24. Flow cytometry CD14+ Monocytes	31

Supplementary Tables	32

1. Clinical and demographic data of systemic sclerosis patients and healthy controls	32
2. Number of reads after HiCUP alignment and filtering of pCHi-C data	33
3. Number of reads from RNA-seq data	35

4. Number of SSc GWAS SNPs and significant interactions in each locus	36

5. Protein-Protein interaction network from genes with significant interactions in CD4^+^ T cells and CD14^+^ monocytes	37

6. Number of significant pCHi-C interactions and captured promoters identified by group	44

7. Differentially expressed genes in SSc vs. controls CD4^+^ T cells.	44

8. Differentially expressed genes in SSc vs. controls CD14^+^ T cells.	45

9. Gene set enrichment analysis of differentially expressed genes in SSc vs. controls CD4^+^ T cells.	47

10. Gene set enrichment analysis of overexpressed genes in CD4^+^ T cells.	49

11. Gene set enrichment analysis of overexpressed genes in CD14^+^ monocytes.	52

12. Differential expression corresponding to genes with significant interactions overlapping SSc GWAS loci in CD4^+^ T cells vs CD14^+^ monocytes.	63

13. Differential interaction values corresponding to genes with significant interactions overlapping SSc GWAS loci in CD4^+^ T cells vs CD14^+^ monocytes	64

14. Gene set enrichment analysis of genes interacting with SSc GWAS loci overlapping enhancer regions in CD4^+^ T cells	65

15. Gene set enrichment analysis of genes interacting with SSc GWAS loci overlapping enhancer regions in CD14^+^ monocytes	65

References	66

**Material and methods**

**Isolation of CD4^+^ T cells and CD14^+^ monocytes**

Primary CD4^+^ T cells and CD14^+^ monocytes were collected from 10 systemic sclerosis patients and 5 healthy individuals (cohorts characteristics described in **Supplementary Table 1**) with informed consent and with ethical approval by the “Comité Ético de Investigación Provincial de Granada” (CEI) from Junta de Andalucía. Patients were not involved in the design and conduct of this research. All SSc patients were diagnosed according to the American College of Rheumatology (ACR)/European Alliance of Associations for Rheumatology (EULAR) 2013 criteria (1). Peripheral blood mononuclear cells (PBMCs) were isolated from 70 ml blood samples using Ficoll density gradient centrifugation. EasySep Human CD14^+^ Positive selection kit (StemCell Technologies, ref:17858) was used to isolate CD14^+^ cells from PBMCs and, subsequently, Easysep CD4^+^ T cell isolation kit (StemCell Technologies, ref:17952) was used to isolate CD4^+^ T cells from the remaining PBMCs, according to the manufacturer’s instructions. Performance of the isolation strategy was assessed using Flow Cytometry Assay on a test sample. CD4^+^ T cells had a purity of 99.8% and a viability of 93.9% in the selected cell population which is 95.7% of the total counts (**supplementary figure 23**) whilst CD14^+^ Monocytes had a purity of 100% and a viability of 76.3% in the selected cell population which is 86.5% of the total counts (**supplementary figure 24**).

**Promoter capture Hi-C probe design**

First, gene annotations for 18,755 protein coding genes were extracted from Ensembl’s genebuild database version 97 GRCh38. The transcription start sites (TSS) for each gene was located using the first base of the gene coordinates with respect to gene orientation. Capture regions were identified by mapping the TSS coordinates to the *in silico* digested (Arima Hi-C) genome and extracting the fragments containing the TSS coordinates as well as the fragments directly upstream and downstream. Therefore, each TSS is represented by a total of 3 contiguous restriction fragments. The average length of the 3 consecutive restriction fragments for each TSS is 786bp and the median is 927bp, with a range of 54-4174bp. If the length of the restriction fragment is no more than 700bp, the entire restriction fragment is covered by the probes. If the length is more than 700bp, then the middle part will not be in the probe design. Using this final set of restriction fragments, a BED file was prepared for input into the SureDesign (Agilent) probe design web tool. Probes were designed using a 1X tiling approach, with moderate repeat masking and maximum performance boosting optimized for the SureSelect XT HS and XT Low Input capture workflows.

**Capture Hi-C library generation**

5-10 million isolated CD4^+^ T cells and CD14^+^ monocytes were crosslinked for 10 min in 1% formaldehyde and agitated at room temperature, the reaction was then quenched with ice cold 0.125 M glycine for 5 min. Crosslinked cells were washed in ice-cold PBS, and the supernatant was discarded, the pellets were then stored at -80ºC.

Each Hi-C library was prepared from fixed cells following the Arima HiC kit (Arima Genomics) and the KAPA HyperPrep kit (KAPA Biosystems) following the manufacturer’s protocol. Briefly, crosslinked cells were lysed, and DNA was digested by two different restriction enzymes, and ligated. The ligated DNA was reverse-crosslinked and fragmented by sonication (Covaris S220), followed by DNA size selection, biotin enrichment, adaptor ligation, and final library amplification. A quality control step to determine the number of PCR cycles needed for library amplification was performed for each library using the NEBNext Library Quant kit (NEBNext, ref: E7630). Final quality and quantity were assessed by Bioanalyzer 2100 (Agilent) and Qubit (Thermo Fisher).

Hi-C samples were then hybridized with the SureSelect custom capture library by following Agilent SureSelectXT HS reagents (ref: G9702A, G9496A) and protocols (ref: G9702-9000). Briefly, target regions are hybridized with designed probes, followed by capture enrichment of these regions through streptavidin pulldown using Dynabeads MyOne Streptavidin T1 (ThermoFisher, ref: 65601). Post-capture amplification was carried out using nine PCR cycles. Final quality and quantity were assessed by Bioanalyzer 2100 (Agilent) and Qubit (Thermo Fisher).

**Promoter capture Hi-C sequencing and processing**

Sequencing for 30 prepared pCHi-C libraries was performed using five lanes of Illumina NovaSeq S4 flow cell on Illumina NovaSeq6000, generating 150 bp paired-end reads, and leading to an average of 500 million reads per sample (Novogene Company LTD). Sequencing data was filtered and the adapters were removed using fastp v0.19.4 (2). Subsequent mapping with GRCh38 and filtering was performed with HiCUP v0.7.4 (3) and bowtie2 v2.3.2 (4), taking as maximum and minimum di-tag lengths 700 and 100, respectively. Only intrachromosomal interactions were included in the analysis, and off-target di-tags where neither end mapped to a targeted fragment were removed (Statistics in **Supplementary Table 2**). Significant chromatin interactions were identified using CHiCAGO v1.13.1 (5) using a threshold of CHiCAGO score > 5 in different conditions; cell type: CD4^+^ T cells (n=15 biological replicates) and CD14^+^ monocytes (n=15); cell type and disease state: CD4^+^ T cells from SSc patients (n=10) and healthy controls (n=5), and CD14^+^ monocytes from SSc patients (n=10) and healthy controls (n=5). Principal component analysis (PCA) was performed in each cell type in order to detect potential biases and the two first PCs were plotted using R 3.6.1 (**Supplementary Figure 1**). Libraries generated using the Arima Hi-C protocol have different characteristics from the original CHi-C protocol, for this reason, several changes were made to the analysis. The restriction fragments were binned for the analysis as follows: every 20 consecutive restriction fragments are binned into one bin; if a baited region is encountered, a separate bin is created for the baited region that includes the 3 captured fragments and the fragment before and after for a total of 5 fragments per region captured. Consecutive baits were merged into a single bait. This design leads to a baitmap of 17,313 baited regions, capturing 18,630 promoters. Default settings were used for the CHiCAGO pipeline, except for design file parameters minFragLen and maxFragLen, set to 140 and 20000, respectively. The weights of the model used by CHiCAGO in the defined conditions were re-estimated using the function “fitDistCurve.R” included in CHiCAGO package after running the pipeline on a merged file including the total 30 samples.

In order to detect enrichment of features in the interactions obtained with CHiCAGO for each cell type, narrowPeak bed files of H3K4me3 and H3K27ac were obtained as follows: H3K4me3 (ENCODE; ENCFF828YVL) and H3K27ac (ENCODE; ENCFF790PVU) in primary CD4^+^ naive T cells; H3K4me3 (ENCODE; ENCFF651GXK) and H3K27ac (ENCODE; ENCFF471QGU) in primary CD14^+^ monocytes. The “peakEnrichment4Features” function from the CHiCAGO package was used to detect enrichment of each feature in pCHi-C data. Finally, Chicdiff v0.6 (6) R package was used to detect differential interactions between different conditions: CD4^+^ T cells vs CD14^+^ monocytes; SSc patients vs healthy controls CD4^+^ T cells; and SSc patients vs healthy controls CD14^+^ monocytes. Code was modified to include age, sex, and disease status (only in cell type comparison) as covariates. For each comparison, only those interactions with CHiCAGO score > 5 in at least one condition were included in differential analysis. Differential interactions with a weighted adjusted p-value < 0.05 were identified as significant. Spearman’s rank-order correlation was performed to test the correlation of log_2_ fold change values in differential interactions between patients and controls in CD4^+^ T cells and CD14^+^ monocytes.

**Genotype calling and allele specific analysis**

Genotypes were called using the Capture Hi-C reads for each individual. Reads were aligned using HiCUP v0.7.4 (3) and bowtie2 v2.3.2 (4) to a masked GRCh38 genome generated using the 1000 genomes project phase 3 SNPs. Genotypes were then called using GLIMPSE v1.1.1 (7) and the 1000 genomes phase 3 reference, making sure that all known restriction sites were excluded from the first step of the genotype calling. Two samples were checked against previously generated imputed genotype array data and resulted in 99.27% and 99.32% concordance. Genotype phasing was then carried out using the integrated phasing pipeline (8) which integrates population phasing using SHAPEIT2 (9) with reads phasing using the Capture Hi-C reads using HAPCUT2 (10), resulting in around 2 million heterozygous SNPs phased per sample. Aligned reads (to masked genome) for each library were then split using SNPsplit v0.5.0 (11) generating allele specific alignments. Counts were for each allele for each significant loop were then calculated using bedtools pair_to_pair (12) and data was integrated in Python 3.9. Allelic imbalance of the reads for each loop that overlapped a SNP was tested using a binomial test as implemented in scipy for each sample which was heterozygous for that SNP. In addition, we filtered SNP/loop pairs so that at least 2 samples were heterozygous and phased for the SNP, at least 10 reads were present for each SNP and at least 1 read for each allele had to be present. All the resulting values were then checked for directionality so that all samples had to have the same directionality in the imbalance and the p-values were merged using the Firsher’s method for meta-analysis as implemented in scipy. Resulting p-values were then corrected using Benjamini-Hochberg method as implemented in statsmodels (13).

**RNA-seq library generation**

A total of 0.5 million purified cells was resuspended in 700 uL Qiazol lysis reagent (QIAGEN, ref: 79306) to isolate RNA, then 140 uL of chloroform was added. After centrifugation at 12000 x g for 15 min, approximately 350 uL of the upper layer containing the RNA was transferred and mixed with 525 uL of 100% ethanol. RNA isolation was continued from this point using the RNeasy microkit (QIAGEN, ref: 74004) reagents and protocol. Final quantity was assessed by Qubit (Thermo Fisher). Libraries for RNA-seq were prepared using Illumina Truseq Stranded Total RNA reagents and protocol, except for Control 1 CD4^+^ and CD14^+^ samples, for which library preparation failed. Library quality and quantity was assessed by Bioanalyzer. The 28 libraries generated were sequenced using three lanes on Illumina HiSeq4000, generating 75 bp paired-end reads, and leading to an average of 30 million reads per sample (Genomics Facility, University of Manchester).

**RNA-seq data processing**

RNA-seq reads were quality trimmed and adapters were removed using fastp v0.19.4 (2). Reads were then mapped using STAR v2.7.3a (14) on the GRCh38 genome with GENCODE annotation v32. Reads were de-duplicated with Picard tools v2.22.2 (“Picard Toolkit.” 2019. Broad Institute, GitHub Repository. http://broadinstitute.github.io/picard/; Broad Institute) and then counted using HTSeq v0.12.3 (15) (**Supplementary Table 3**). Final count matrices were analysed in R 3.6.1, using edgeR v3.28.1 (16) to perform normalization and differential expression analysis. Three differential expression comparisons were performed: CD4^+^ T cells vs CD14^+^ monocytes; SSc patients vs healthy controls in CD4^+^ T cells; and SSc patients vs healthy controls in CD14^+^ monocytes. Age, sex, and disease status (only in cell type comparison) were used as covariates. Only those genes with a mean of more than 1 log_2_ transformed counts per million (CPM) across all samples were included in further analysis, for a total of 11,221 coding genes. Differentially expressed genes were called with an adjusted p-value of 0.1 (FDR 10%). Due to the extreme differences in expression regarding the CD4^+^ vs CD14^+^ comparison, only those with an absolute value of log_2_ fold change (|log_2_FC|) > 2 and FDR < 5% were taken in further analysis. Functional enrichment analyses were performed with g:Profiler (17) with default settings, taking gene ontology and biological pathways as data sources.

**Linking differential expression and differential interactions in CD4^+^ T cells vs CD14^+^ monocytes**

Genes corresponding with the promoter end of significant differential interactions observed between CD4^+^ and CD14^+^ cells were overlapped with those differentially expressed. One-sided Fisher’s exact test was performed in order to calculate the enrichment of genes with differential interactions in those differentially expressed. In this set of overlapping genes, Spearman’s rank-order correlation was performed to test the correlation of log_2_ fold change values in differential interactions and differential expression. The log_2_ fold change value for each gene with differential interactions was obtained as the median log_2_ fold change value of all interactions corresponding to a specific promoter. Finally, in order to test the distribution of log_2_ fold change values, a binomial exact test was performed on a subset of overlapping genes obtained adding a more stringent cutoff, including only those differentially interacting genes with an absolute value of median log_2_FC > 2 for each gene. Functional enrichment analyses were performed with g:Profiler (17) with default settings, taking gene ontology and biological pathways as data sources.

**Defining SSc GWAS loci**

All independent non-MHC disease-associated signals for SSc were selected from the largest meta-GWAS performed to date (18). We defined 23 regions based on linkage disequilibrium data and SNP proximity from the total of 27 independent signals described in GWAS. First, we took all SNPs with GWAS level significant association (p-value < 5x10^-8^) (427 SNPs) and calculated SNPs in high linkage disequilibrium with those SNPs (r^2^ > 0.8) from meta-GWAS datasets (18) using PLINK v1.07 (19), resulting in a total of 1,505 SNPs. To facilitate pCHi-C analysis, independent signals corresponding to the same locus were grouped. These grouped independent signals interacted with the same promoters, and did not represent a change if they were taken as separated loci. The window ranges and total number of SNPs in each of the 23 final loci are specified in **Supplementary Table S4**.

**Defining enhancers and TADs in CD4^+^ T cells and CD14^+^ monocytes**

In order to define enhancer regions, chromHMM v1.22 annotations (20) from 9 CD4^+^ T cells (BSS00183, BSS00185, BSS00186, BSS00188, BSS00189, BSS00190, BSS00191, BSS00192 and BSS00274) and 4 CD14^+^ monocytes (BSS00178, BSS00179, BSS00180, and BSS00181) were downloaded from the EpiMap project (21). For each cell type, enhancer regions were defined as those with state number from chromHMM corresponding to enhancer activity (7, 8, 9 ,10, 11, and 15) present in at least one sample. TADs definition for CD4^+^ T cells and CD14^+^ monocytes were obtained from Javierre et al. (22).

**Overlap between pCHi-C, SSc GWAS loci, and enhancer regions**

In order to prioritize certain interactions observed in pCHi-C data of particular interest in SSc GWAS loci, the SNP set previously defined in “Defining SSc GWAS loci'' was overlapped with enhancer regions of each cell type using the GenomicRanges (23) package implemented in R 3.6.1 (**Supplementary Table 4**). This new SNP set was then overlapped with the promoter interacting regions (PIRs) of significant pCHi-C interactions, defining candidate interacting genes as those in which their PIR overlaps with our significant SSc SNP set and enhancer regions. Garfield v2 (24) was used to estimate the enrichment of the GWAS SNPs in CD4^+^ T cells and CD14^+^ monocytes enhancer regions using a p-value threshold of 1x10^-8^. A test for equality of proportions or two proportion z-test (“prop.test” in R 3.6.1) was performed to calculate the enrichment of SNPs overlapping enhancer regions between cell types, correcting by total number of base pairs covered by enhancer regions for each cell type. For each candidate interacting gene, the median of log_2_FC values and weighted adjusted p-values was calculated taking all differential interactions with the PIR overlapping our SSc SNP set and enhancer regions. Functional enrichment analyses were performed for the sets of interacting genes observed in CD4^+^ T cells and CD14^+^ monocytes with g:Profiler (17) using default settings, taking gene ontology and biological pathways as data sources.

**Visualization tools**

The WashU Epigenome browser (25) was used to plot pCHi-C interactions, enhancer regions defined by chromHMM and H3K27ac peaks (from EpiMap (21) samples previously defined), and TADs (from Javierre et al. (22)) in CD4^+^ T cells and CD14^+^ monocytes. Juicebox (26) was used to further investigate and visualize our pCHi-C data.

**Drug target analysis**

In order to assess if genes interacting with SSc GWAS loci in CD4^+^ T cells and CD14^+^ monocytes presented potential drug targets that could be repurposed for its use in SSc, those genes interacting with PIR overlapping significant SSc GWAS SNPs and enhancer regions, were used to model a protein-protein interaction (PPI) network using STRING v11 (27) with the highest interaction confidence score (>0.9), calculated as a combined probability from different evidences of interactions corrected for the probability of observing a random interaction (**Supplementary Table 5**). Protein products from these genes and those in direct PPI with them were used to query the OpenTargets Platform (28) for drug targets. Additionally, the same platform and the Drugbank database (29) were searched for information on clinical studies of drug targets of interest in SSc. Only drug targets with at least completed phase III clinical trials in SSc and/or similar immune-mediated diseases were included in **Table 2**.

**Supplementary Figures**

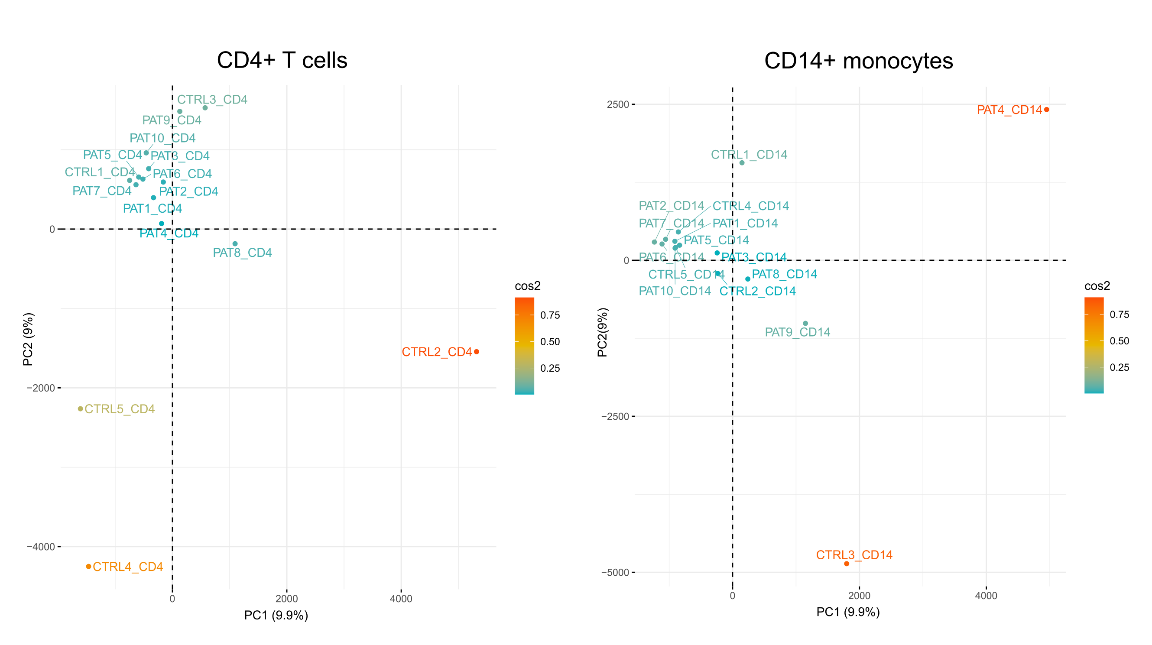
**Supplementary Figure 1.** Representation of the first two principal components (PC) from pCHi-C data for each sample in CD4^+^ T cells and CD14^+^ monocytes. The percentage of explained variance of each PC is written in brackets. The cos2 gradient represents the quality of representation (in percentage) of each sample for these two specific PCs.

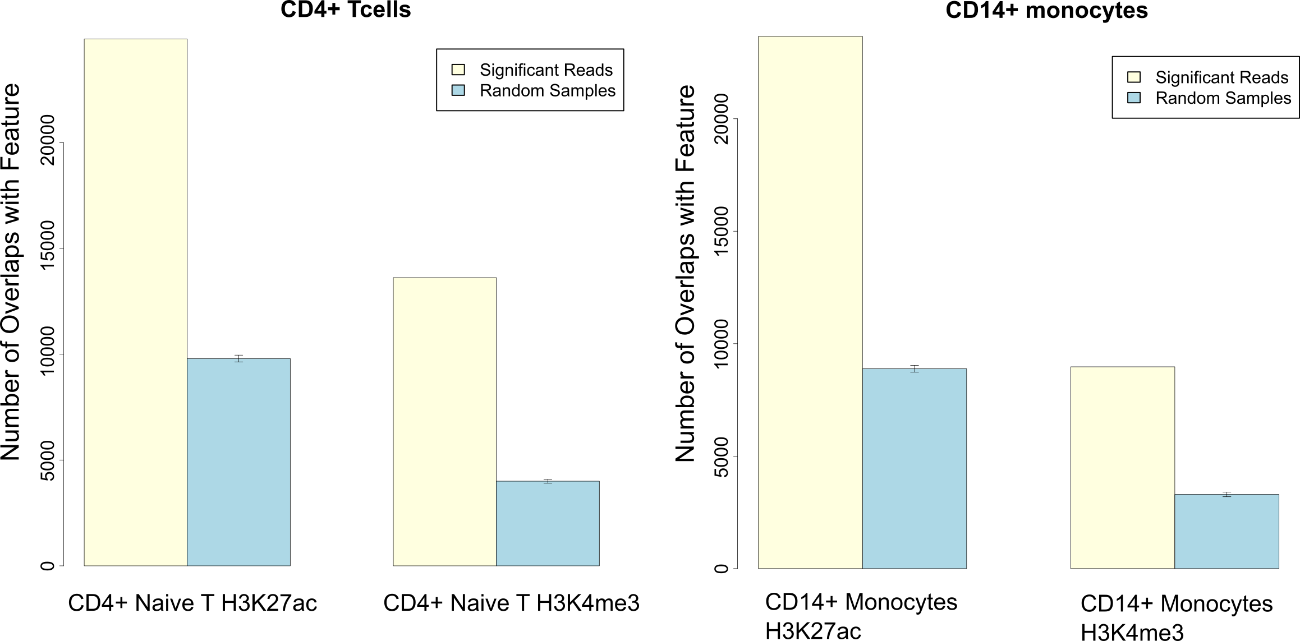

**Supplementary Figure 2.** Enrichment of features within promoter interacting regions (PIRs) of pCHi-C interactions. Peak locations of H3K4me3 and H3K27ac (ENCODE) in primary CD4^+^ naive T cells and CD14^+^ monocytes were tested against PIRs of interactions using the “peakEnrichment4Features” function of CHiCAGO package. The graphs show the number of overlaps with the feature in the interaction data (yellow) vs the mean number of overlaps in 100 sampled interactions from the non-significant pool (blue). Error bars correspond to the 95% confidence interval.

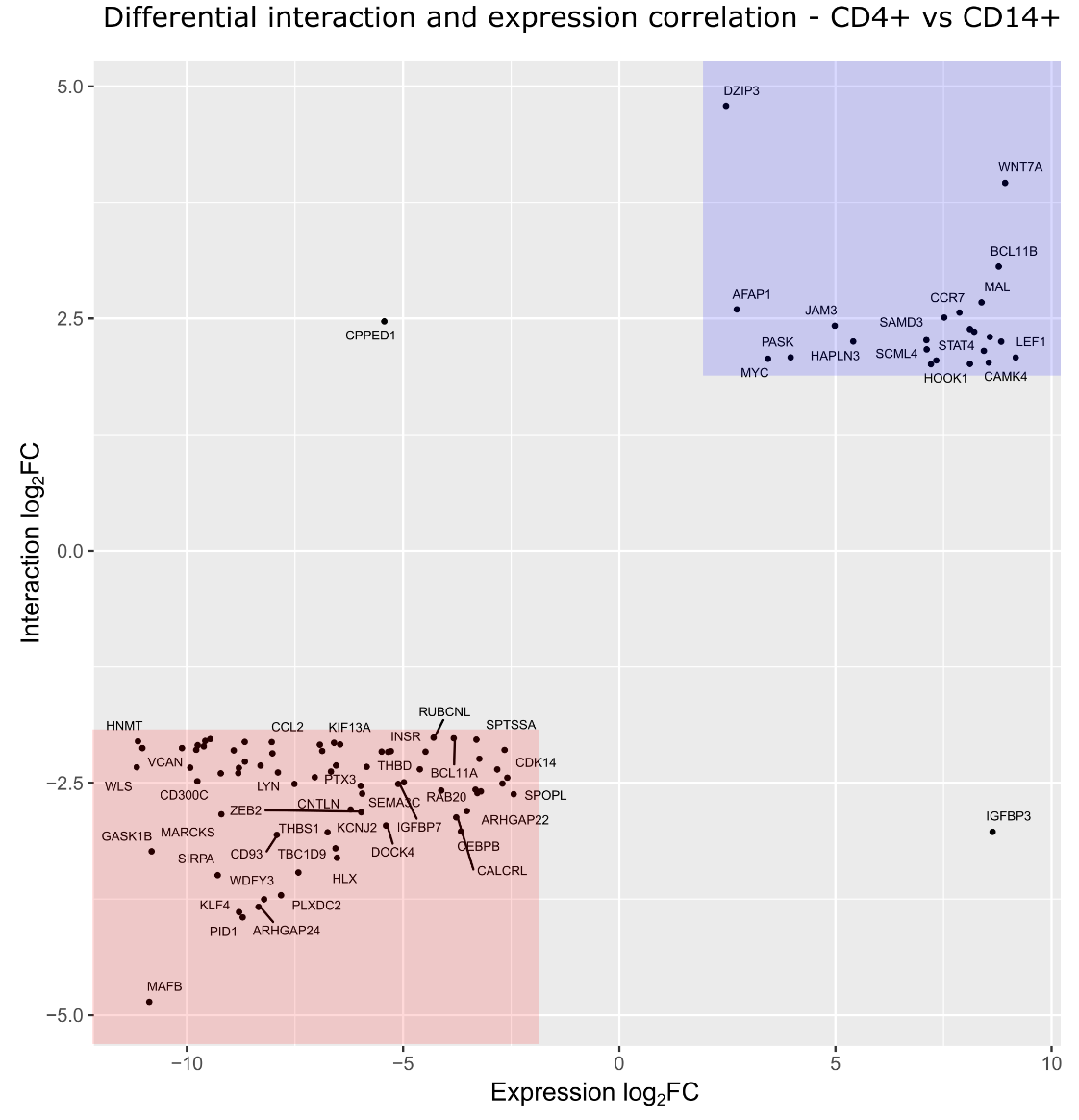

**Supplementary Figure 3.** Distribution of the 97 differentially expressed genes (|log_2_FC| > 2) overlapping differentially interacting genes (|log_2_FC| > 2) in CD4^+^ T cells vs CD14^+^ monocytes comparison. The blue rectangle represents the region including genes which are overexpressed (log_2_FC > 2) and present more interactions (log_2_FC > 2) in CD4^+^ T cells as compared with CD14^+^ monocytes. The red rectangle represents the region including genes which are overexpressed (log_2_FC < -2) and present more interactions (log_2_FC < -2) in CD14^+^ monocytes as compared with CD4^+^ T cells.

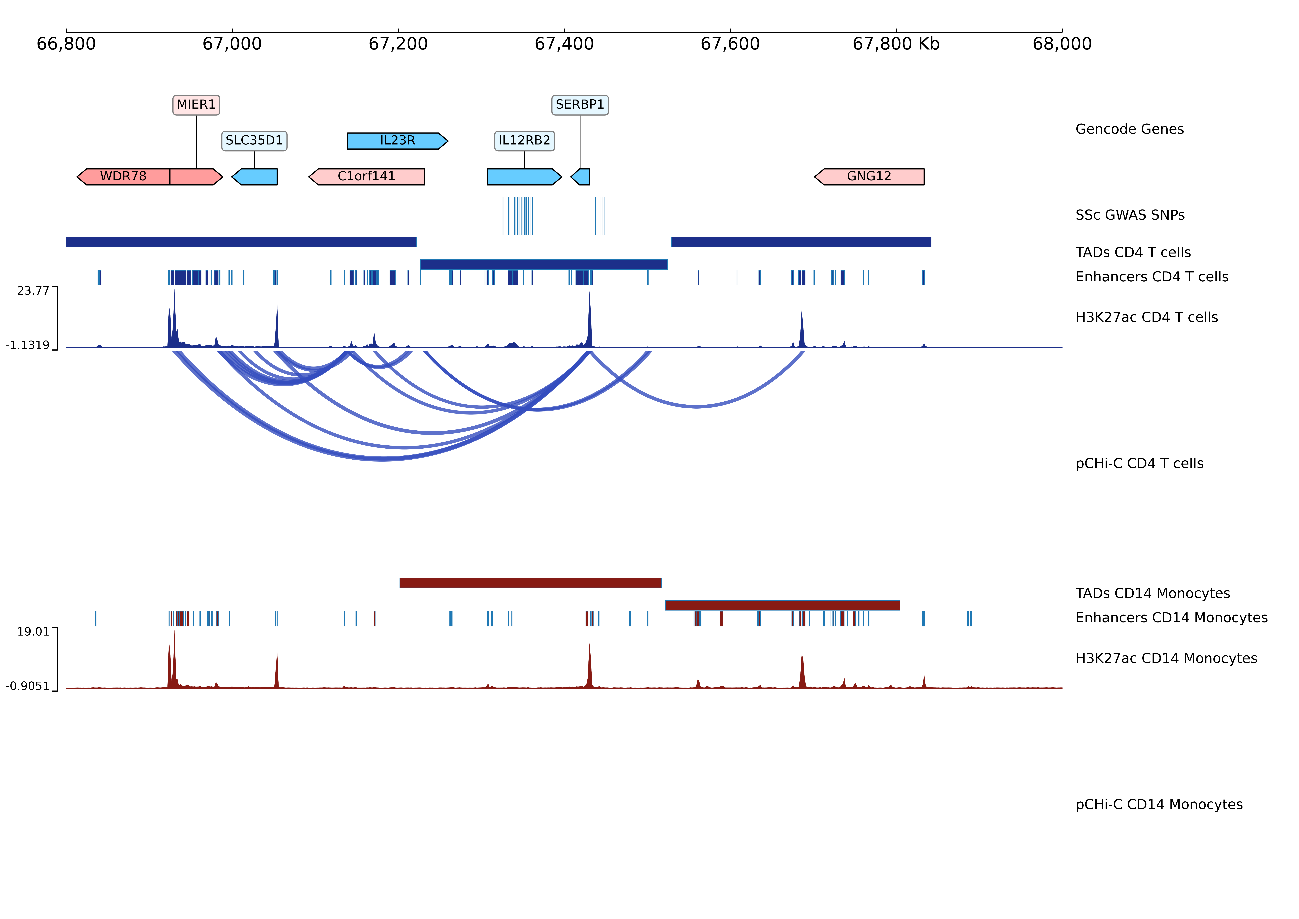
**Supplementary Figure 4.** Promoter capture Hi-C (pCHi-C) interactions in the rs3790566 *(IL12RB2)* locus. Genomic coordinates (GRCh38) are shown at the top of the panel. The tracks include Gencode Genes, systemic sclerosis (SSc)-associated SNPs from López-Isac *et al* (18) and their proxies (r2>0.8), topologically associating domains (TADs) (shown as bars), H3K27ac signal, enhancer regions as defined by chromHMM, and pCHi-C significant interactions (CHiCAGO score > 5) (shown as arcs) in CD4^+^ T cells (blue) and CD14^+^ monocytes (red).

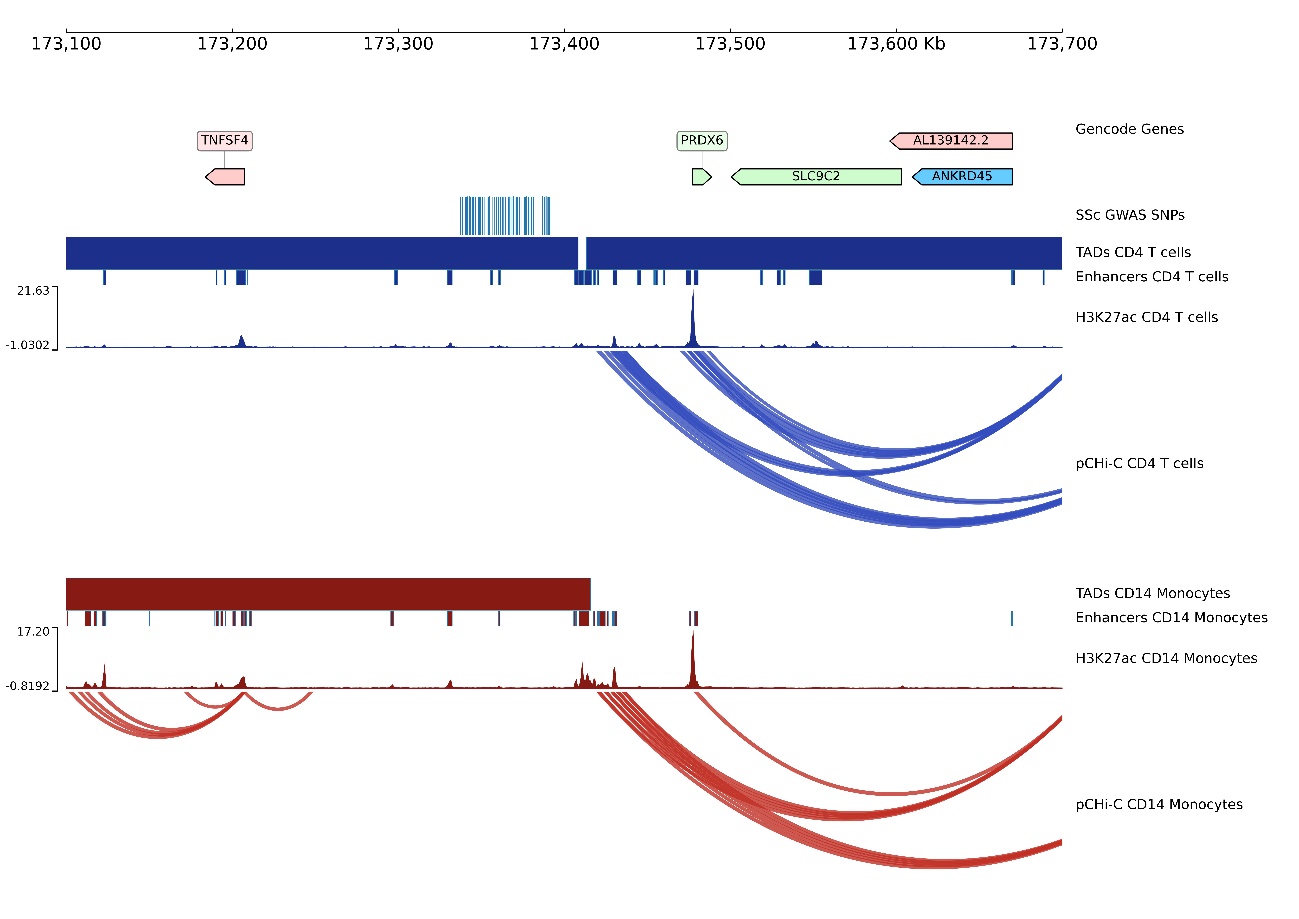

**Supplementary Figure 5.** Promoter capture Hi-C (pCHi-C) interactions in the rs1857066 *(TNFSF4- LOC100506023 - PRDX6)* locus. Genomic coordinates (GRCh38) are shown at the top of the panel. The tracks include Gencode Genes, systemic sclerosis (SSc)-associated SNPs from López-Isac et al (18) and their proxies (r2>0.8), topologically associating domains (TADs) (shown as bars), H3K27ac signal, enhancer regions as defined by chromHMM, and pCHi-C significant interactions (CHiCAGO score > 5) (shown as arcs) in CD4^+^ T cells (blue) and CD14^+^ monocytes (red).

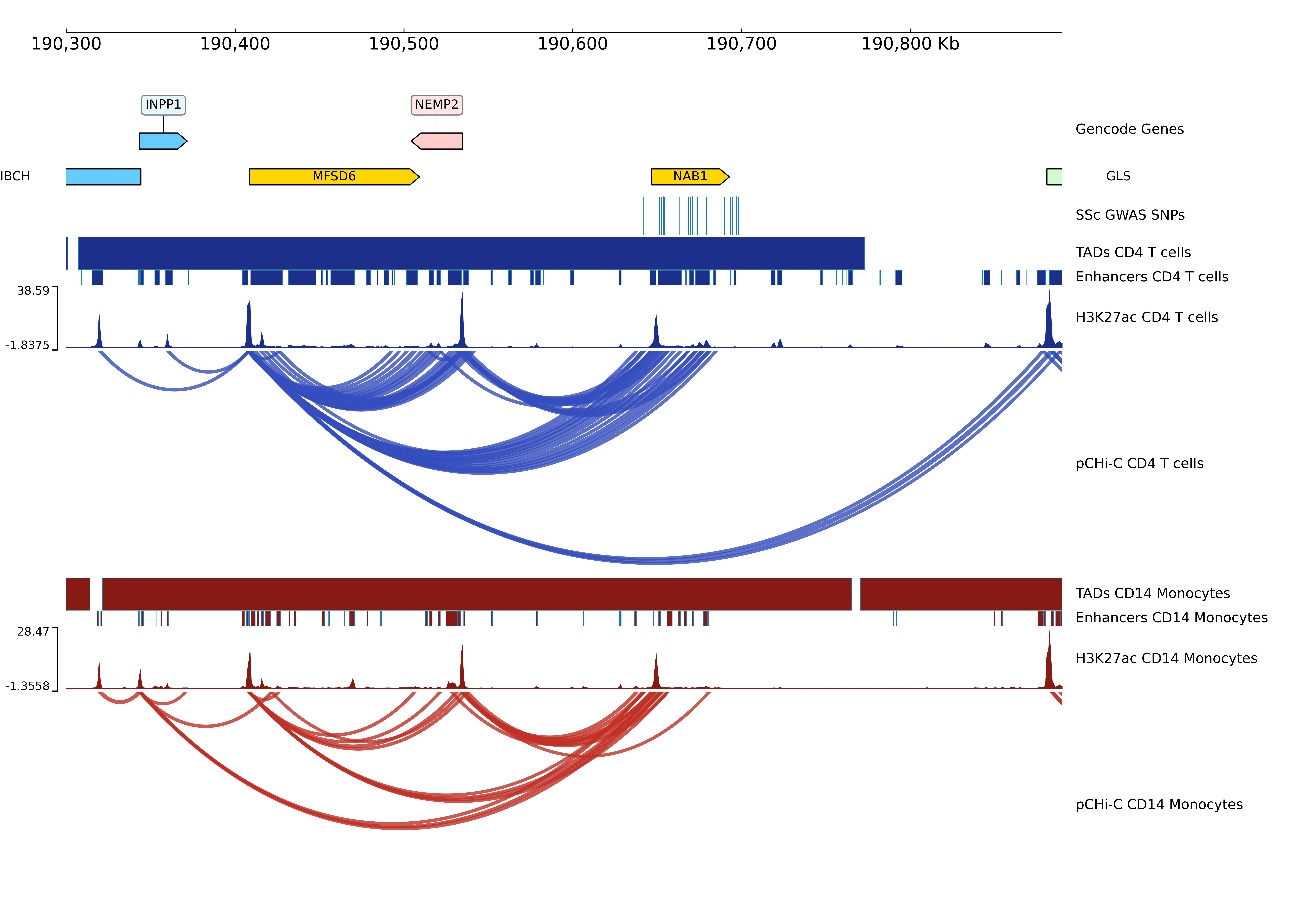

**Supplementary Figure 6.** Promoter capture Hi-C (pCHi-C) interactions in the rs16832798 *(NAB1)* locus. Genomic coordinates (GRCh38) are shown at the top of the panel. The tracks include Gencode Genes, systemic sclerosis (SSc)-associated SNPs from López-Isac et al (18) and their proxies (r2>0.8), topologically associating domains (TADs) (shown as bars), H3K27ac signal, enhancer regions as defined by chromHMM, and pCHi-C significant interactions (CHiCAGO score > 5) (shown as arcs) in CD4^+^ T cells (blue) and CD14^+^ monocytes (red).

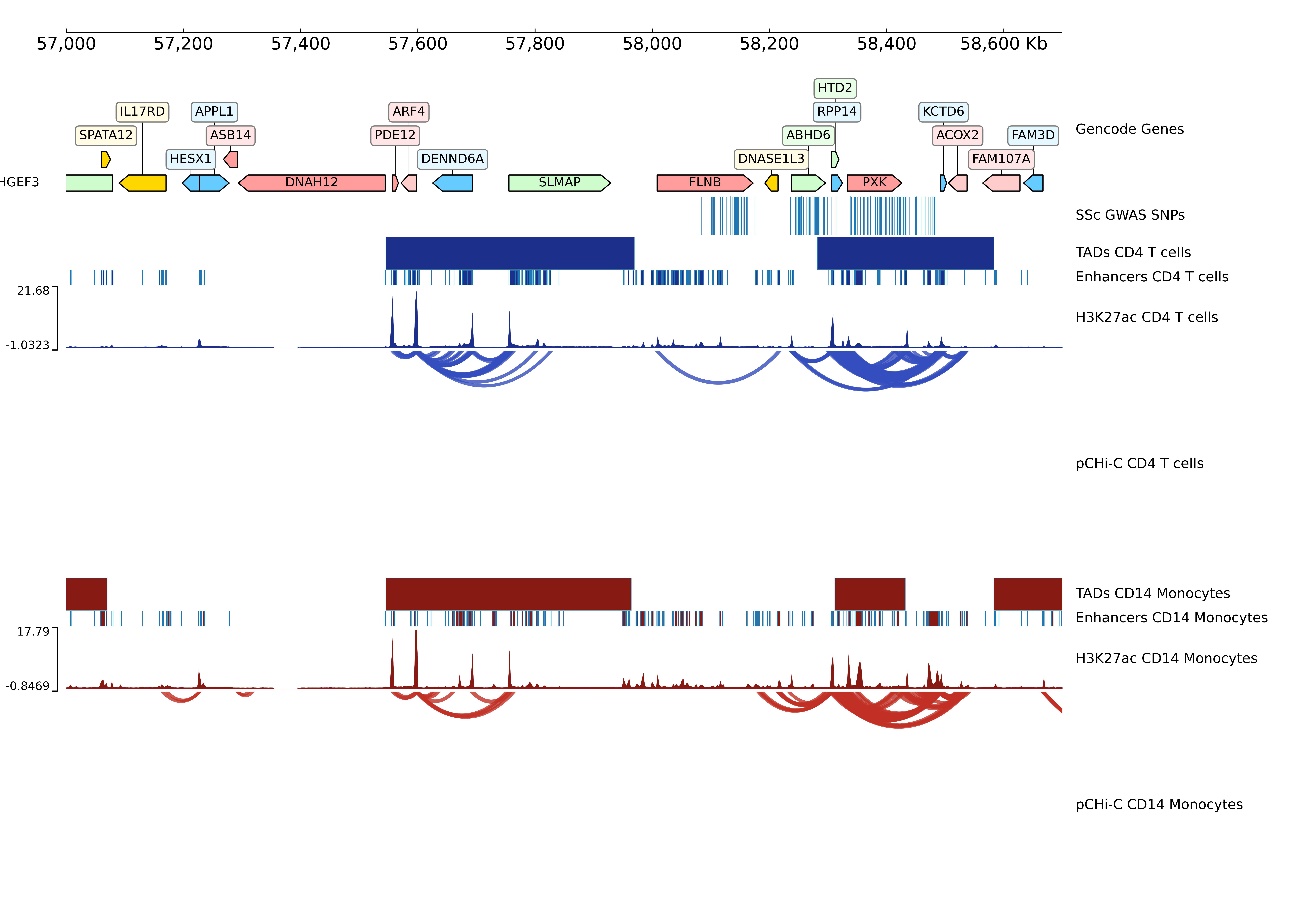

**Supplementary Figure 7.** Promoter capture Hi-C (pCHi-C) interactions in the rs4076852 *(FLNB -DNASE1L3-PXK)* locus. Genomic coordinates (GRCh38) are shown at the top of the panel. The tracks include Gencode Genes, systemic sclerosis (SSc)-associated SNPs from López-Isac et al (18) and their proxies (r2>0.8), topologically associating domains (TADs) (shown as bars), H3K27ac signal, enhancer regions as defined by chromHMM, and pCHi-C significant interactions (CHiCAGO score > 5) (shown as arcs) in CD4^+^ T cells (blue) and CD14^+^ monocytes (red).

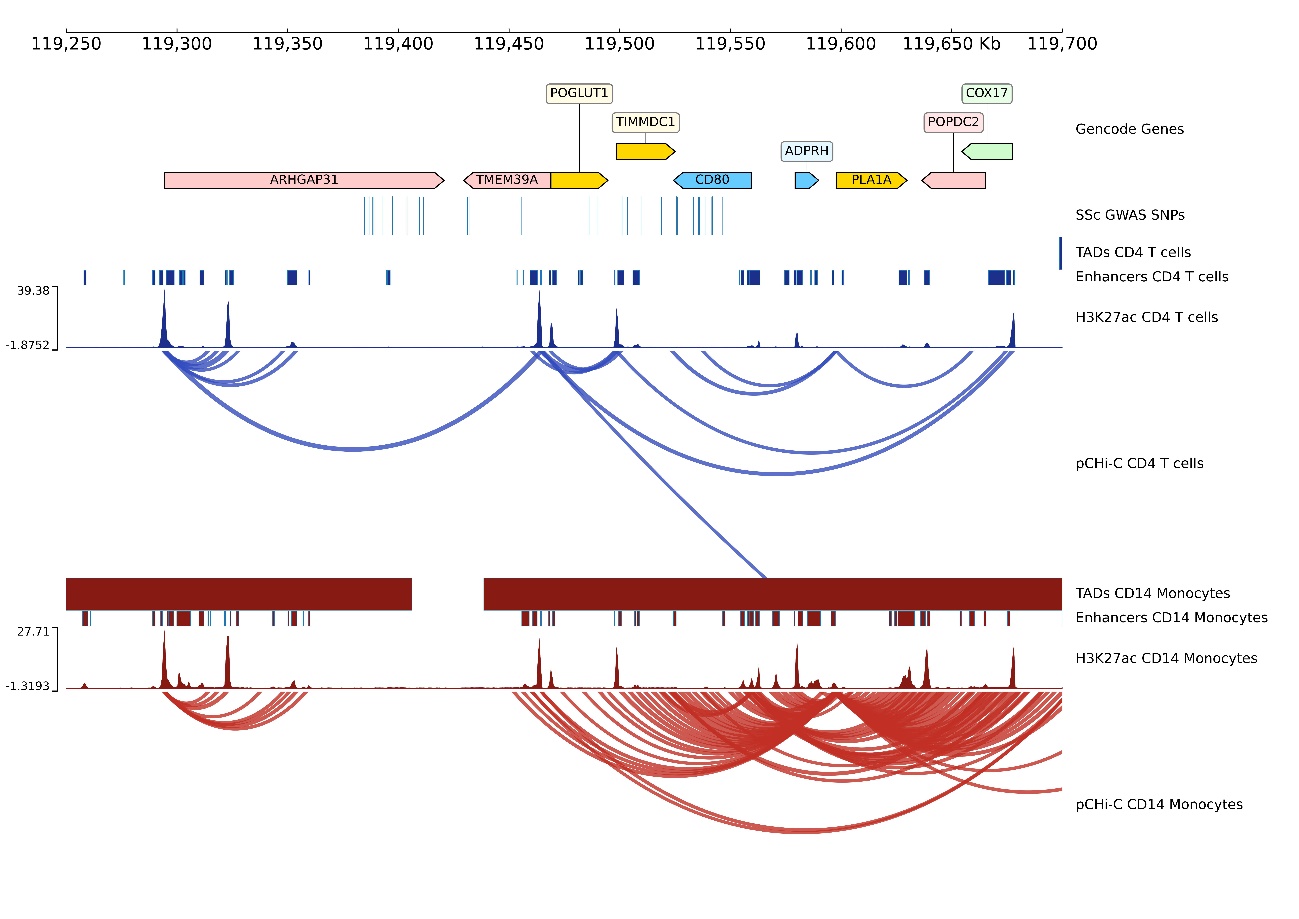

**Supplementary Figure 8.** Promoter capture Hi-C (pCHi-C) interactions in the rs9884090 (*POGLUT1-TIMMDC1-CD80- ARHGAP31*) locus. Genomic coordinates (GRCh38) are shown at the top of the panel. The tracks include Gencode Genes, systemic sclerosis (SSc)-associated SNPs from López-Isac et al (18) and their proxies (r2>0.8), topologically associating domains (TADs) (shown as bars), H3K27ac signal, enhancer regions as defined by chromHMM, and pCHi-C significant interactions (CHiCAGO score > 5) (shown as arcs) in CD4^+^ T cells (blue) and CD14^+^ monocytes (red).

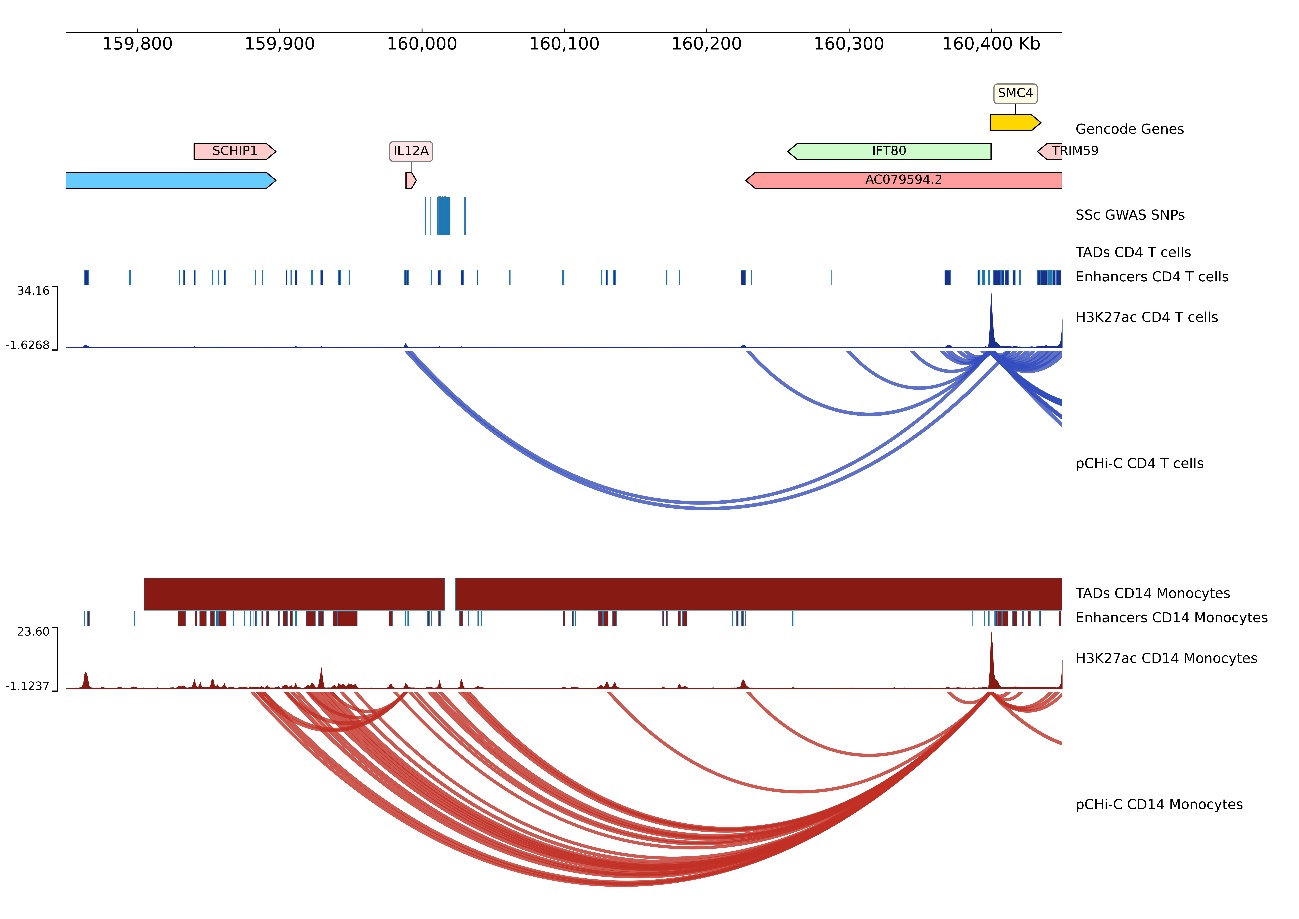

**Supplementary Figure 9.** Promoter capture Hi-C (pCHi-C) interactions in the rs589446 *(IL12A)* locus. Genomic coordinates (GRCh38) are shown at the top of the panel. The tracks include Gencode Genes, systemic sclerosis (SSc)-associated SNPs from López-Isac et al (18) and their proxies (r2>0.8), topologically associating domains (TADs) (shown as bars), H3K27ac signal, enhancer regions as defined by chromHMM, and pCHi-C significant interactions (CHiCAGO score > 5) (shown as arcs) in CD4^+^ T cells (blue) and CD14^+^ monocytes (red).

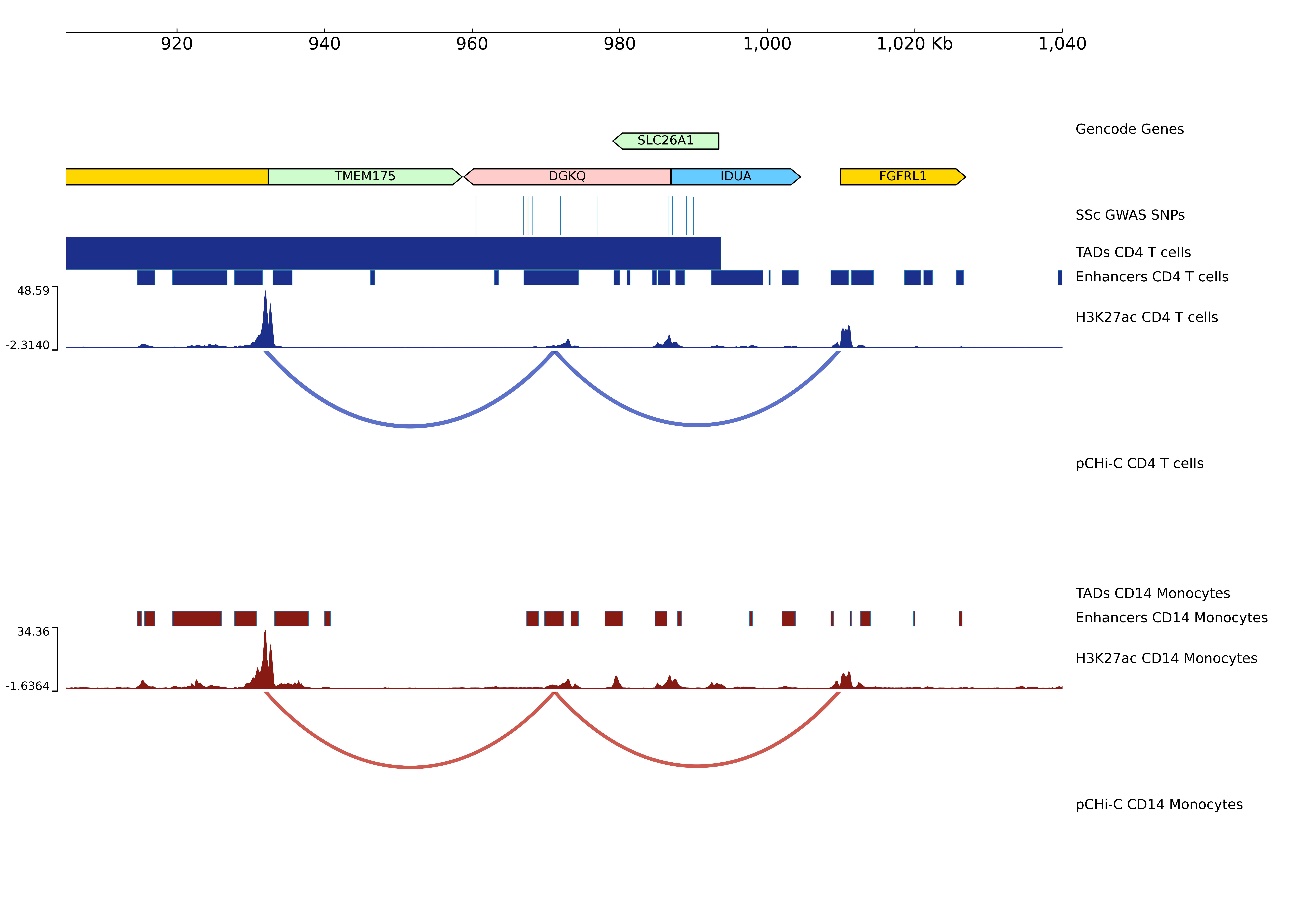
**Supplementary Figure 10.** Promoter capture Hi-C (pCHi-C) interactions in the rs11724804 *(DGKQ)* locus. Genomic coordinates (GRCh38) are shown at the top of the panel. The tracks include Gencode Genes, systemic sclerosis (SSc)-associated SNPs from López-Isac et al (18) and their proxies (r2>0.8), topologically associating domains (TADs) (shown as bars), H3K27ac signal, enhancer regions as defined by chromHMM, and pCHi-C significant interactions (CHiCAGO score > 5) (shown as arcs) in CD4^+^ T cells (blue) and CD14^+^ monocytes (red).

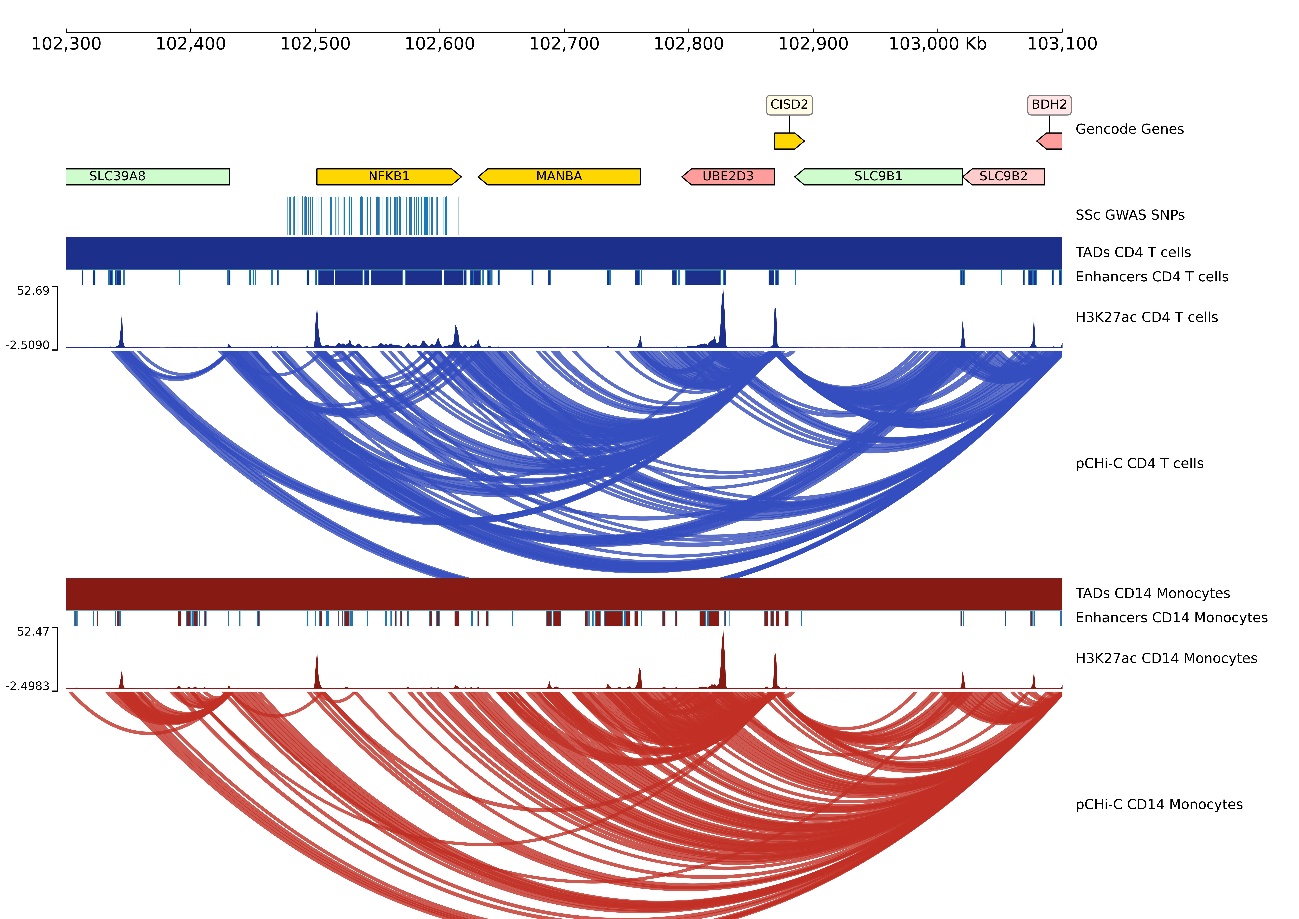

**Supplementary Figure 11.** Promoter capture Hi-C (pCHi-C) interactions in the rs230534 *(NFKB1)* locus. Genomic coordinates (GRCh38) are shown at the top of the panel. The tracks include Gencode Genes, systemic sclerosis (SSc)-associated SNPs from López-Isac et al (18) and their proxies (r2>0.8), topologically associating domains (TADs) (shown as bars), H3K27ac signal, enhancer regions as defined by chromHMM, and pCHi-C significant interactions (CHiCAGO score > 5) (shown as arcs) in CD4^+^ T cells (blue) and CD14^+^ monocytes (red).

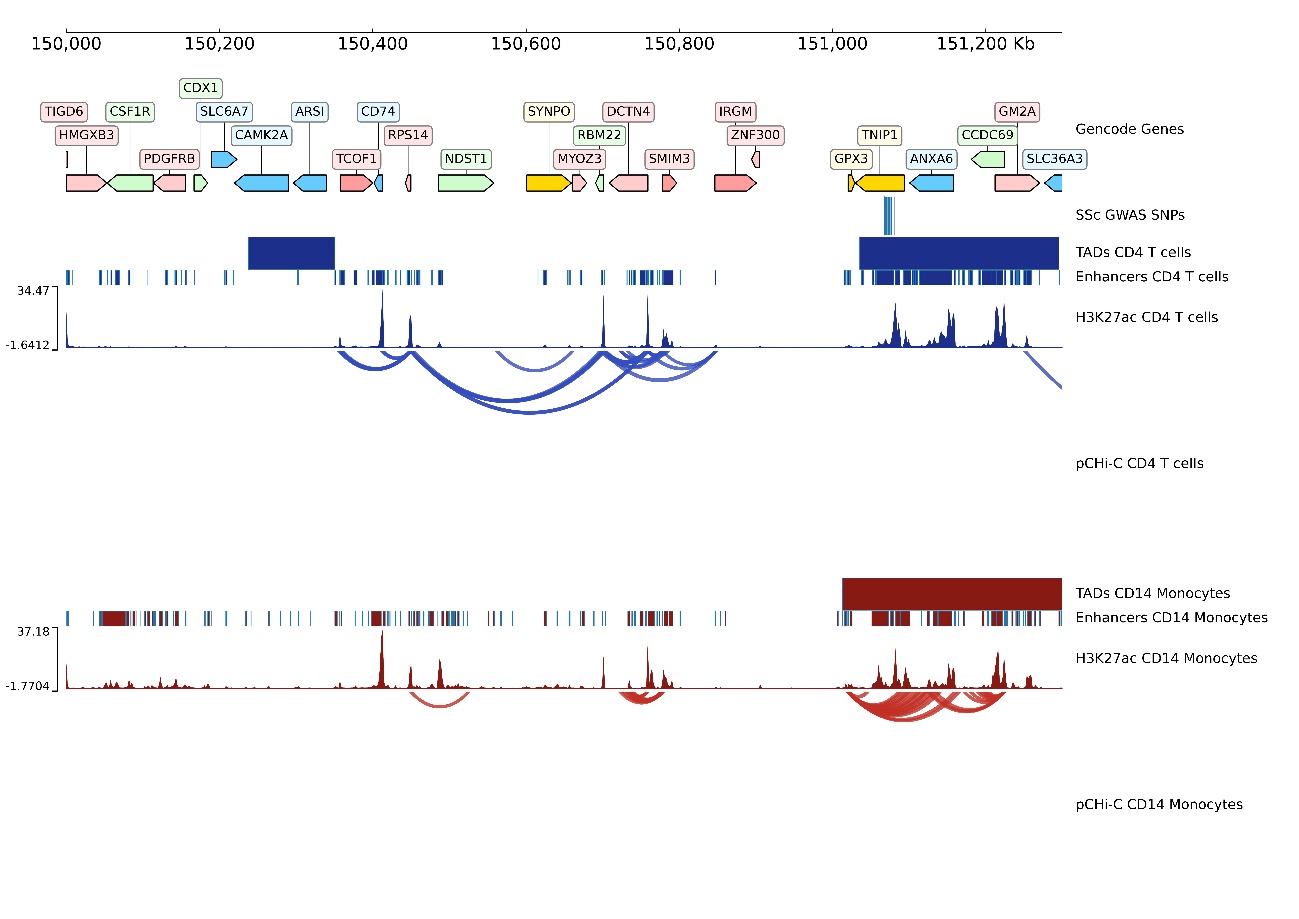

**Supplementary Figure 12.** Promoter capture Hi-C (pCHi-C) interactions in the rs3792783 *(TNIP1)* locus. Genomic coordinates (GRCh38) are shown at the top of the panel. The tracks include Gencode Genes, systemic sclerosis (SSc)-associated SNPs from López-Isac et al (18) and their proxies (r2>0.8), topologically associating domains (TADs) (shown as bars), H3K27ac signal, enhancer regions as defined by chromHMM, and pCHi-C significant interactions (CHiCAGO score > 5) (shown as arcs) in CD4^+^ T cells (blue) and CD14^+^ monocytes (red).

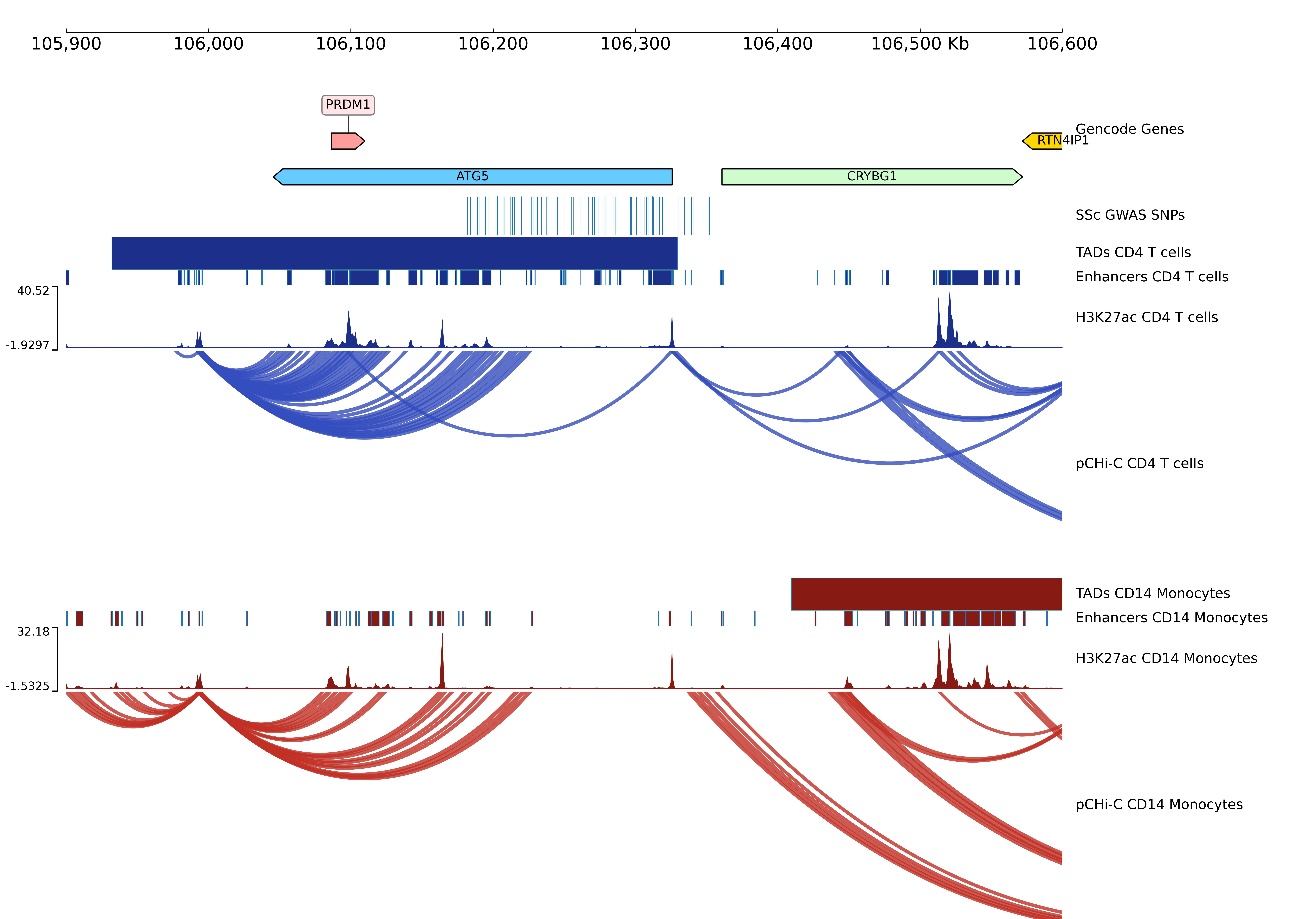

**Supplementary Figure 13.** Promoter capture Hi-C (pCHi-C) interactions and gene expression in the rs633724 *(ATG5)* locus. Genomic coordinates (GRCh38) are shown at the top of the panel. The tracks include Gencode Genes, systemic sclerosis (SSc)-associated SNPs from López-Isac et al (18) and their proxies (r2>0.8), topologically associating domains (TADs) (shown as bars), H3K27ac signal, enhancer regions as defined by chromHMM, and pCHi-C significant interactions (CHiCAGO score > 5) (shown as arcs) in CD4^+^ T cells (blue) and CD14^+^ monocytes (red).

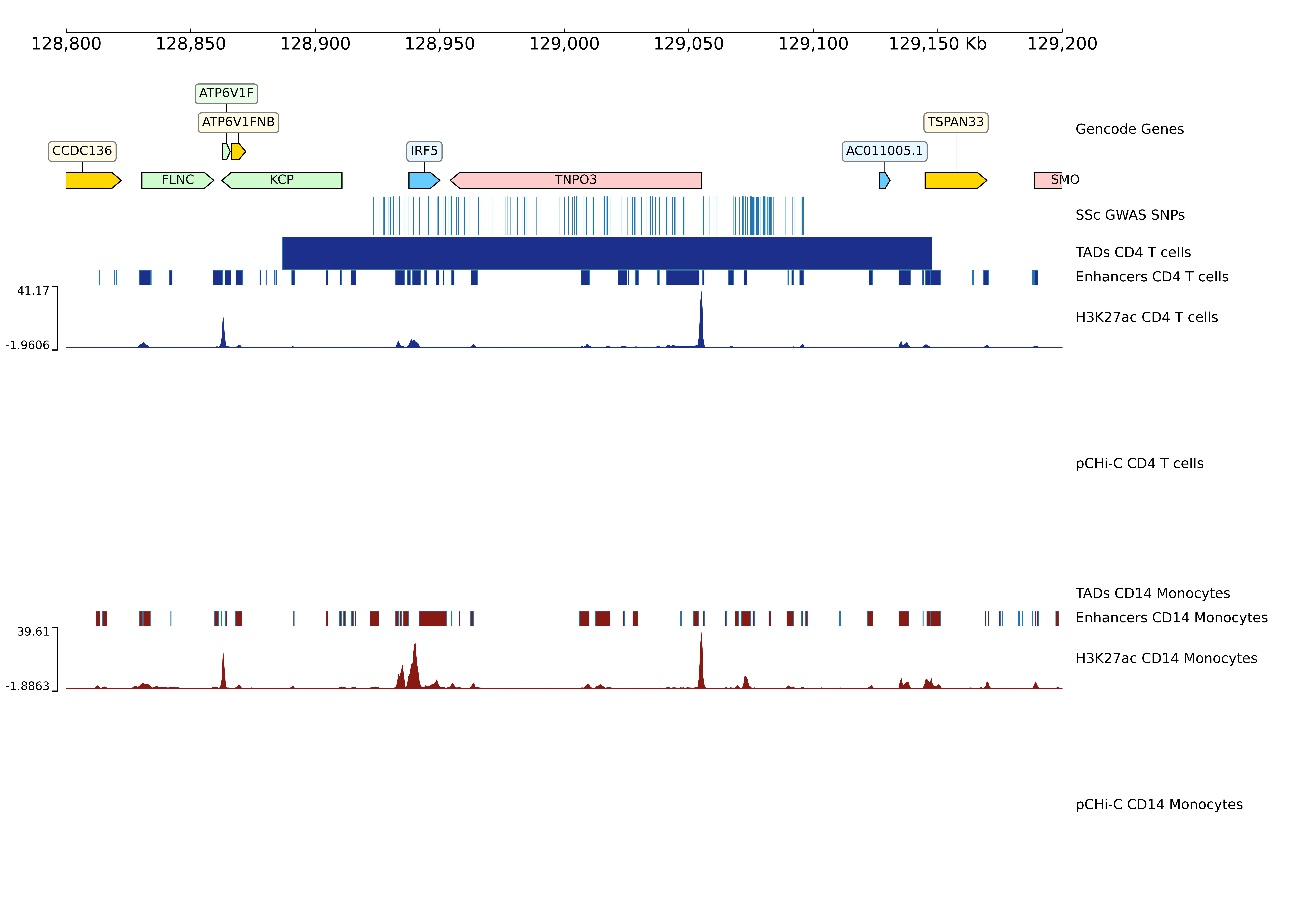

**Supplementary Figure 14.** Promoter capture Hi-C (pCHi-C) interactions in the rs36073657 *(IRF5-TNPO3)* locus. Genomic coordinates (GRCh38) are shown at the top of the panel. The tracks include Gencode Genes, systemic sclerosis (SSc)-associated SNPs from López-Isac et al (18) and their proxies (r2>0.8), topologically associating domains (TADs) (shown as bars), H3K27ac signal, enhancer regions as defined by chromHMM, and pCHi-C significant interactions (CHiCAGO score > 5) (shown as arcs) in CD4^+^ T cells (blue) and CD14^+^ monocytes (red).

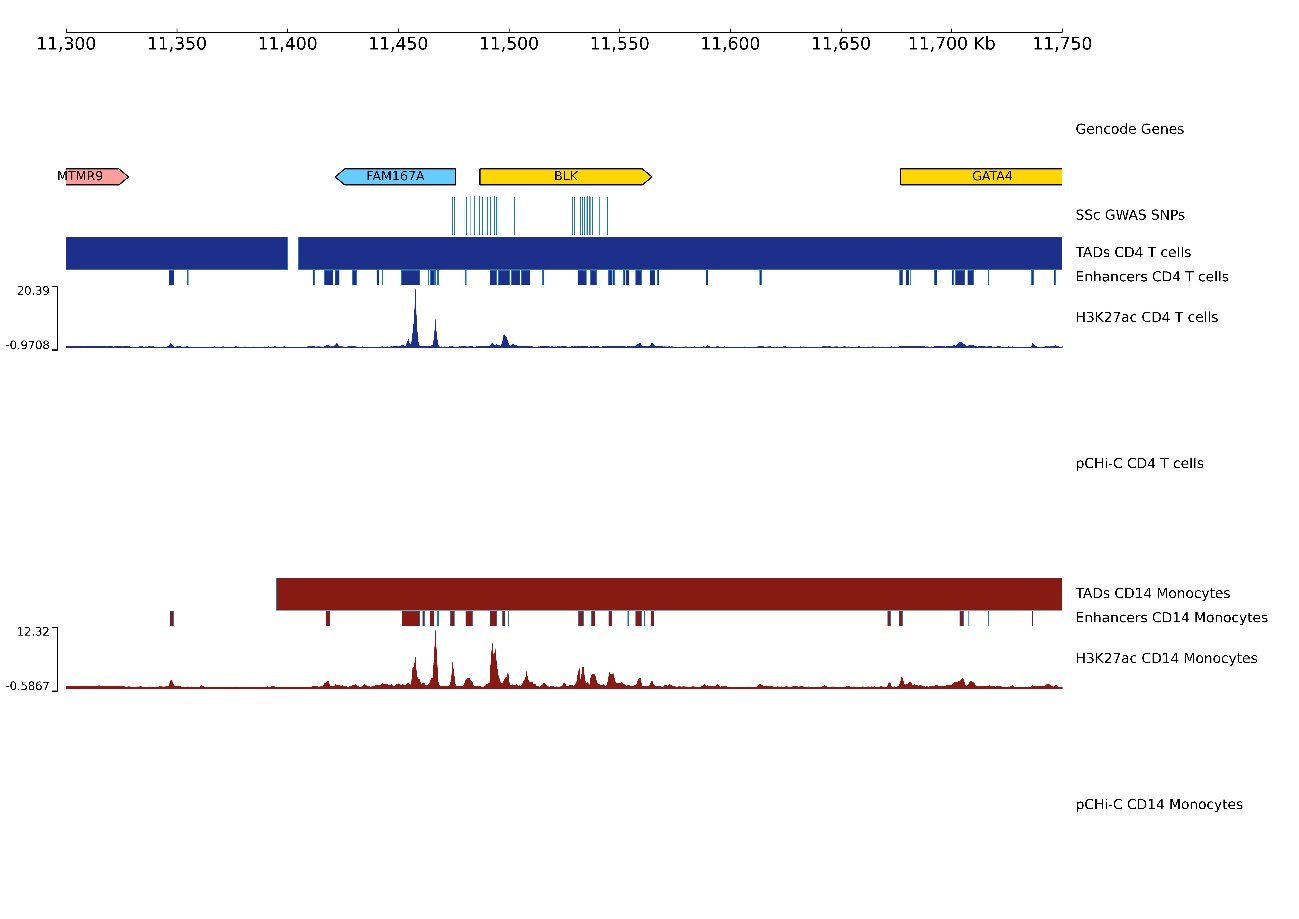

**Supplementary Figure 15.** Promoter capture Hi-C (pCHi-C) interactions in the rs2736340 *(FAM167A-BLK)* locus. Genomic coordinates (GRCh38) are shown at the top of the panel. The tracks include Gencode Genes, systemic sclerosis (SSc)-associated SNPs from López-Isac et al (18) and their proxies (r2>0.8), topologically associating domains (TADs) (shown as bars), H3K27ac signal, enhancer regions as defined by chromHMM, and pCHi-C significant interactions (CHiCAGO score > 5) (shown as arcs) in CD4^+^ T cells (blue) and CD14^+^ monocytes (red).

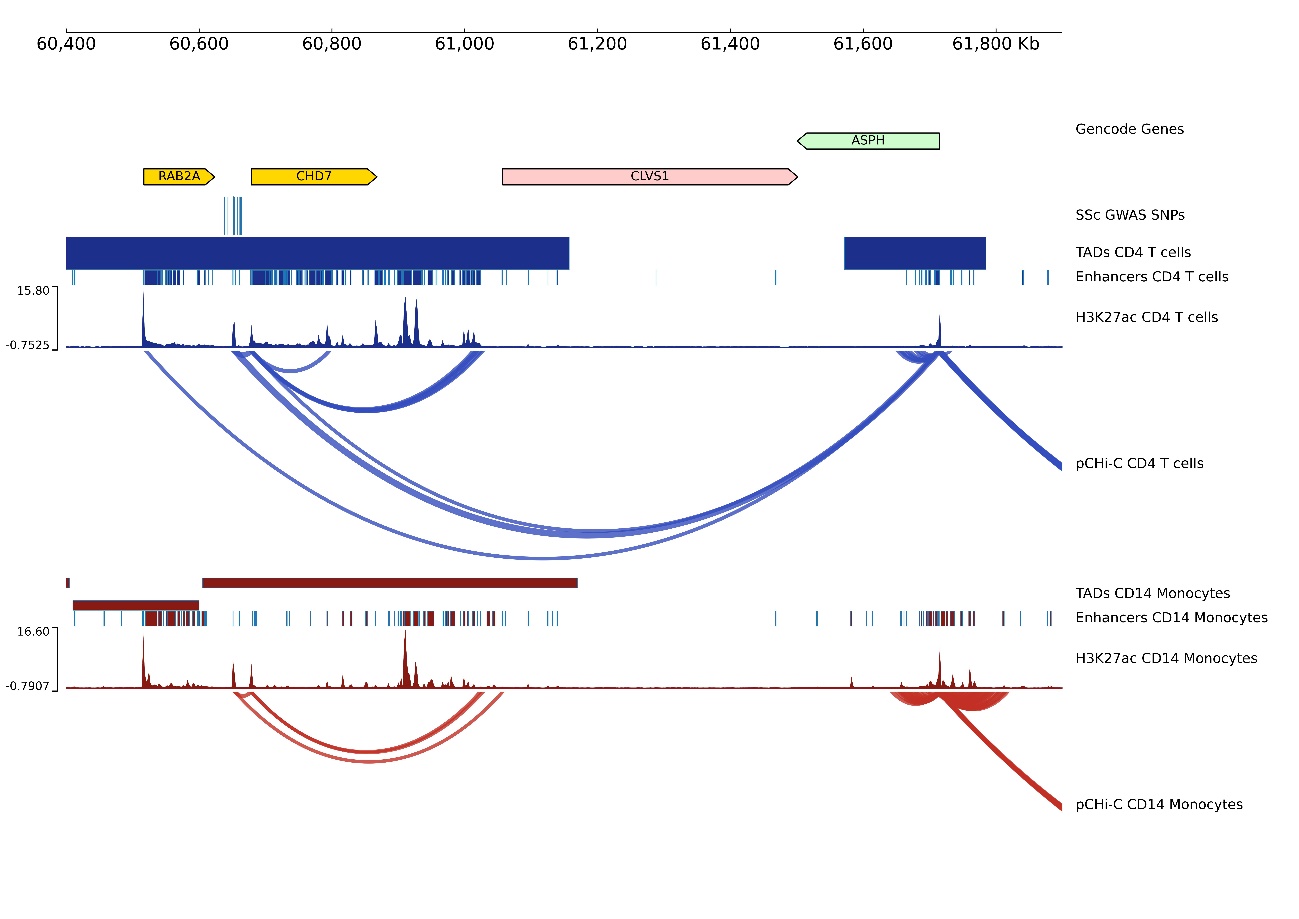

**Supplementary Figure 16.** Promoter capture Hi-C (pCHi-C) interactions in the rs685985 *(RAB2A-CHD7)* locus. Genomic coordinates (GRCh38) are shown at the top of the panel. The tracks include Gencode Genes, systemic sclerosis (SSc)-associated SNPs from López-Isac et al (18) and their proxies (r2>0.8), topologically associating domains (TADs) (shown as bars), H3K27ac signal, enhancer regions as defined by chromHMM, and pCHi-C significant interactions (CHiCAGO score > 5) (shown as arcs) in CD4^+^ T cells (blue) and CD14^+^ monocytes (red).

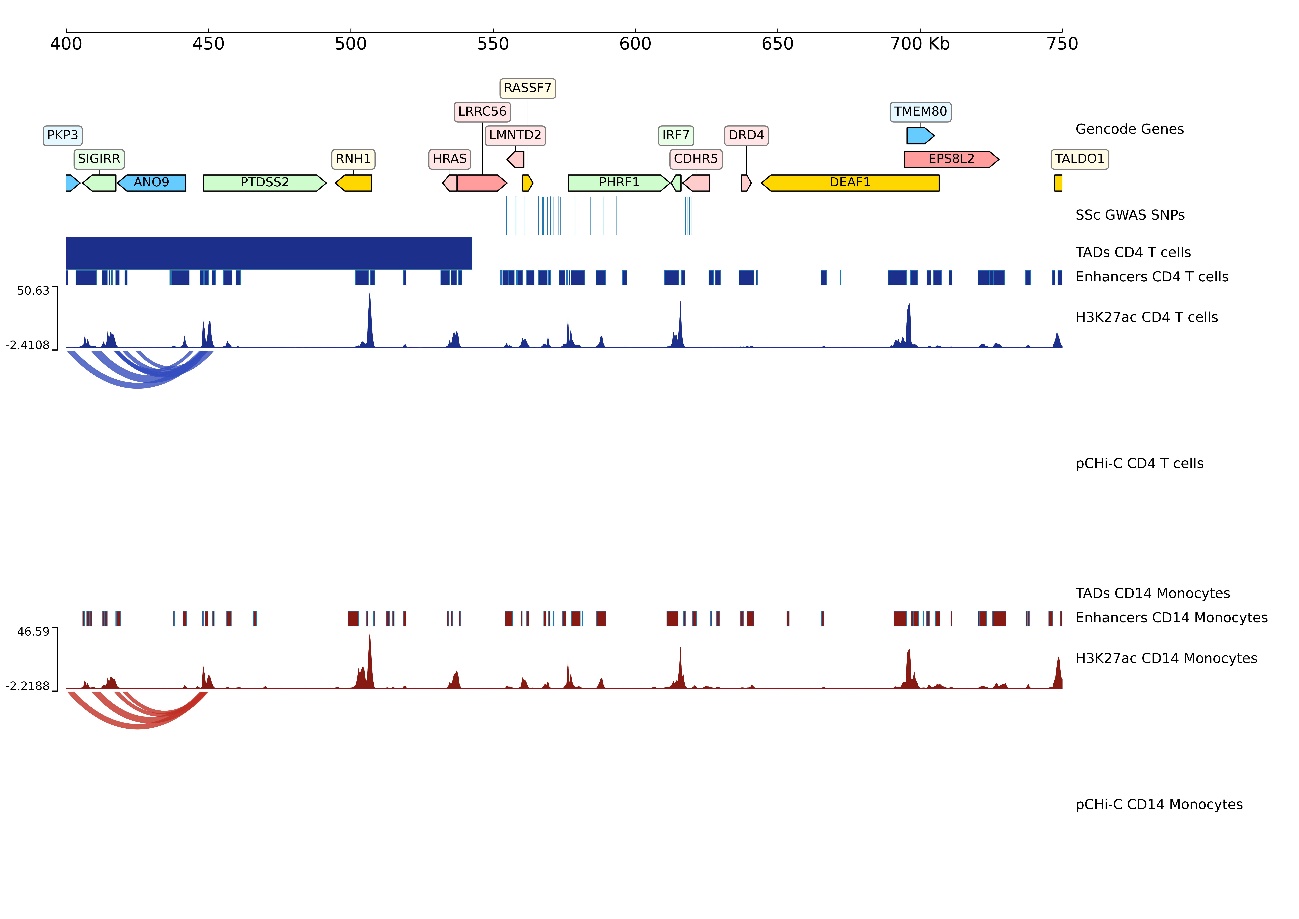

**Supplementary Figure 17.** Promoter capture Hi-C (pCHi-C) interactions in the rs6598008 *(CDHR5 -IRF7)* locus. Genomic coordinates (GRCh38) are shown at the top of the panel. The tracks include Gencode Genes, systemic sclerosis (SSc)-associated SNPs from López-Isac et al (18) and their proxies (r2>0.8), topologically associating domains (TADs) (shown as bars), H3K27ac signal, enhancer regions as defined by chromHMM, and pCHi-C significant interactions (CHiCAGO score > 5) (shown as arcs) in CD4^+^ T cells (blue) and CD14^+^ monocytes (red).

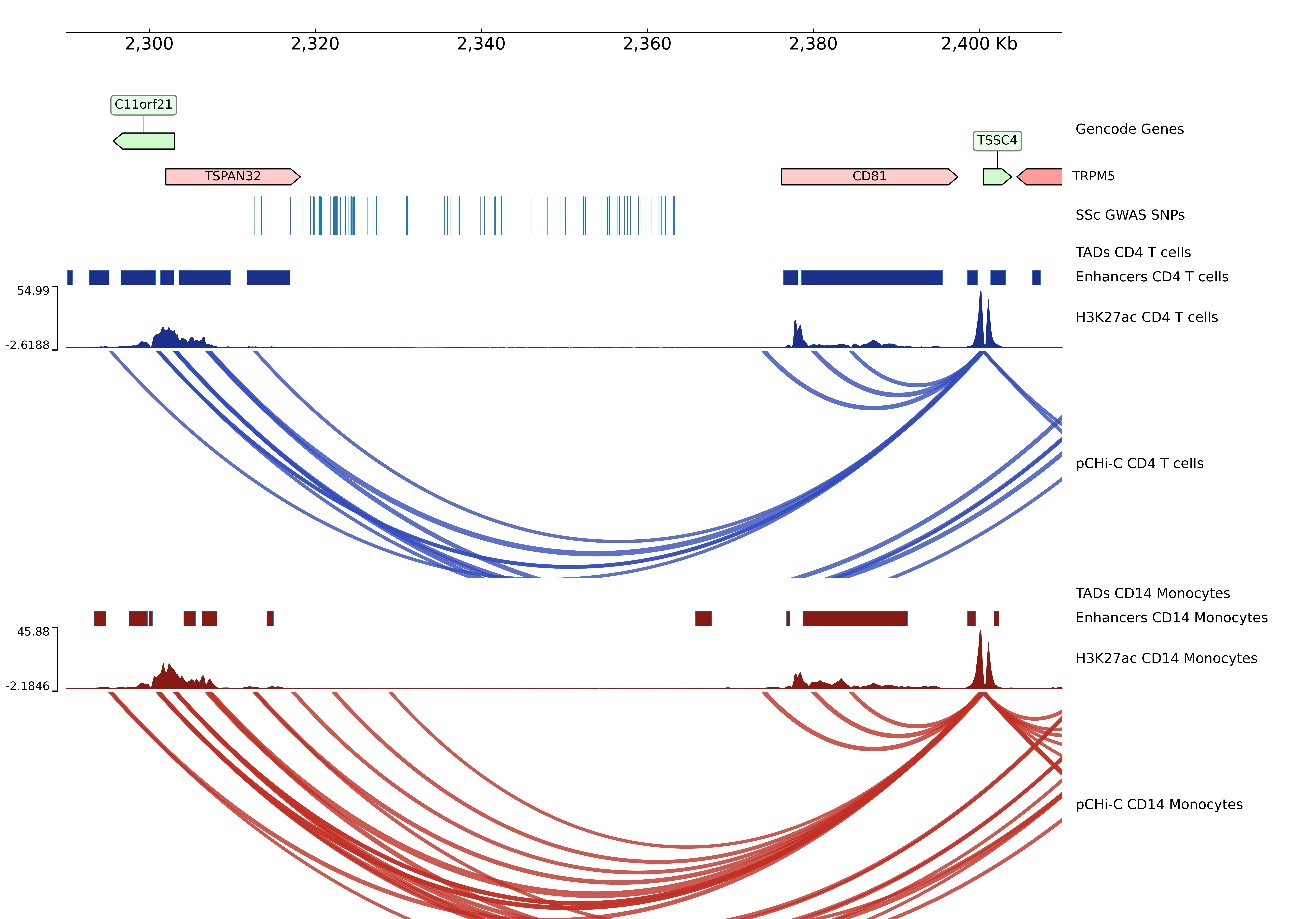

**Supplementary Figure 18.** Promoter capture Hi-C (pCHi-C) interactions in the rs2651804 *(TSPAN32,CD81-AS1)* locus. Genomic coordinates (GRCh38) are shown at the top of the panel. The tracks include Gencode Genes, systemic sclerosis (SSc)-associated SNPs from López-Isac et al (18) and their proxies (r2>0.8), topologically associating domains (TADs) (shown as bars), H3K27ac signal, enhancer regions as defined by chromHMM, and pCHi-C significant interactions (CHiCAGO score > 5) (shown as arcs) in CD4^+^ T cells (blue) and CD14^+^ monocytes (red).

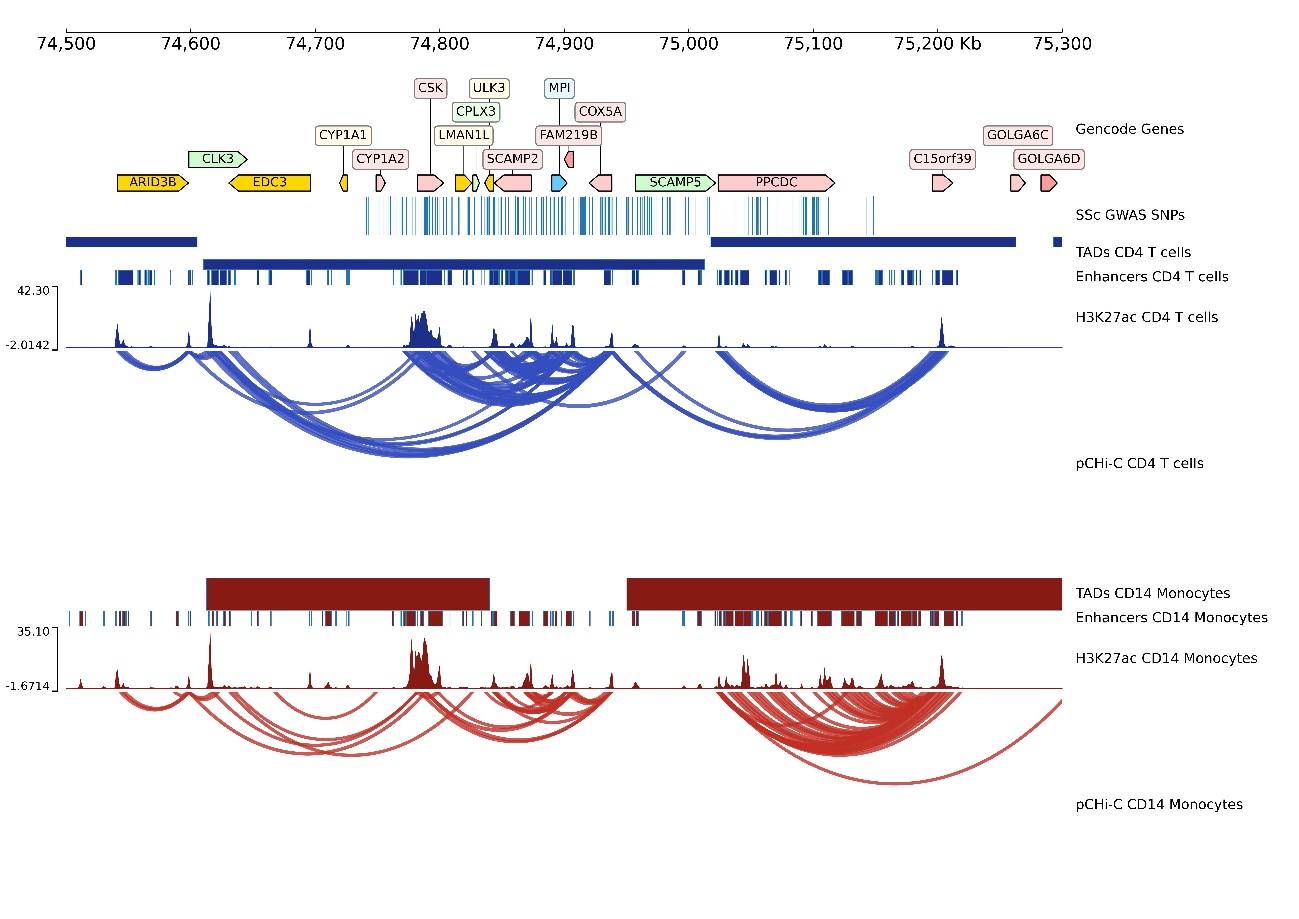

**Supplementary Figure 19.** Promoter capture Hi-C (pCHi-C) interactions in the rs1378942 *(CSK)* locus. Genomic coordinates (GRCh38) are shown at the top of the panel. The tracks include Gencode Genes, systemic sclerosis (SSc)-associated SNPs from López-Isac et al (18) and their proxies (r2>0.8), topologically associating domains (TADs) (shown as bars), H3K27ac signal, enhancer regions as defined by chromHMM, and pCHi-C significant interactions (CHiCAGO score > 5) (shown as arcs) in CD4^+^ T cells (blue) and CD14^+^ monocytes (red).

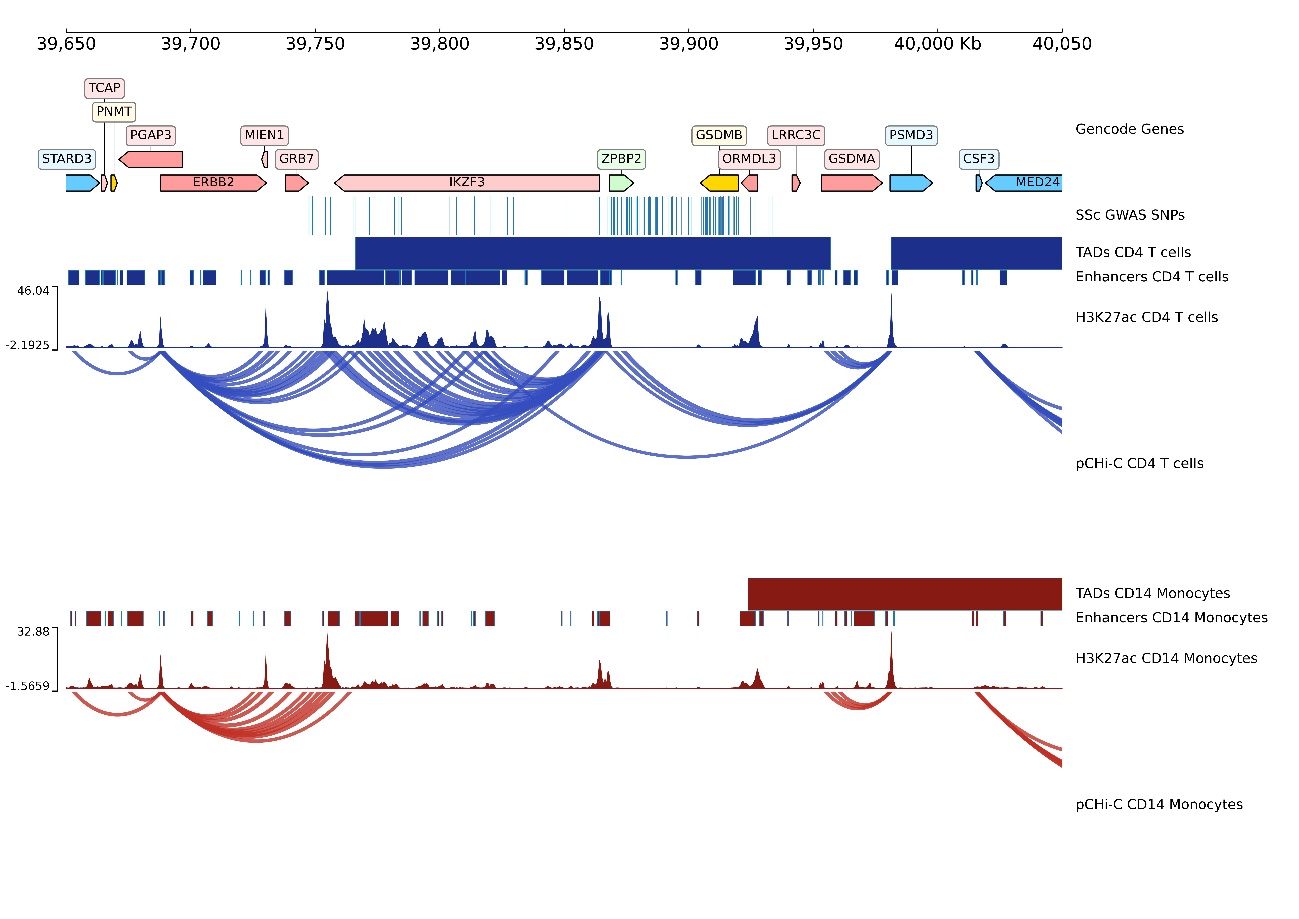

**Supplementary Figure 20.** Promoter capture Hi-C (pCHi-C) interactions in the rs883770 *(IKZF3-GSDMB)* locus. Genomic coordinates (GRCh38) are shown at the top of the panel. The tracks include Gencode Genes, systemic sclerosis (SSc)-associated SNPs from López-Isac et al (18) and their proxies (r2>0.8), topologically associating domains (TADs) (shown as bars), H3K27ac signal, enhancer regions as defined by chromHMM, and pCHi-C significant interactions (CHiCAGO score > 5) (shown as arcs) in CD4^+^ T cells (blue) and CD14^+^ monocytes (red).

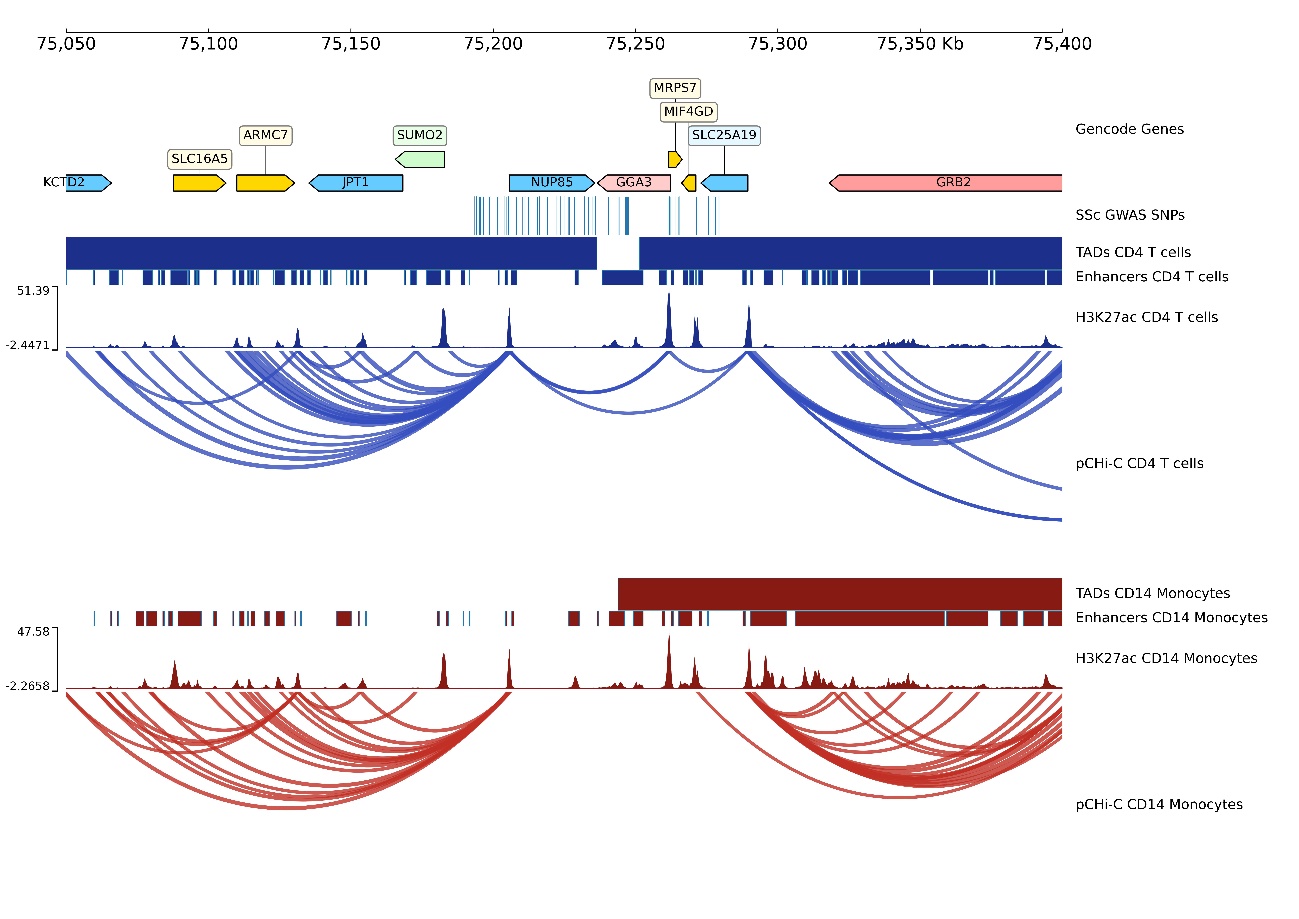

**Supplementary Figure 21.** Promoter capture Hi-C (pCHi-C) interactions in the rs1005714 *(NUP85-GRB2)* locus. Genomic coordinates (GRCh38) are shown at the top of the panel. The tracks include Gencode Genes, systemic sclerosis (SSc)-associated SNPs from López-Isac et al (18) and their proxies (r2>0.8), topologically associating domains (TADs) (shown as bars), H3K27ac signal, enhancer regions as defined by chromHMM, and pCHi-C significant interactions (CHiCAGO score > 5) (shown as arcs) in CD4^+^ T cells (blue) and CD14^+^ monocytes (red).

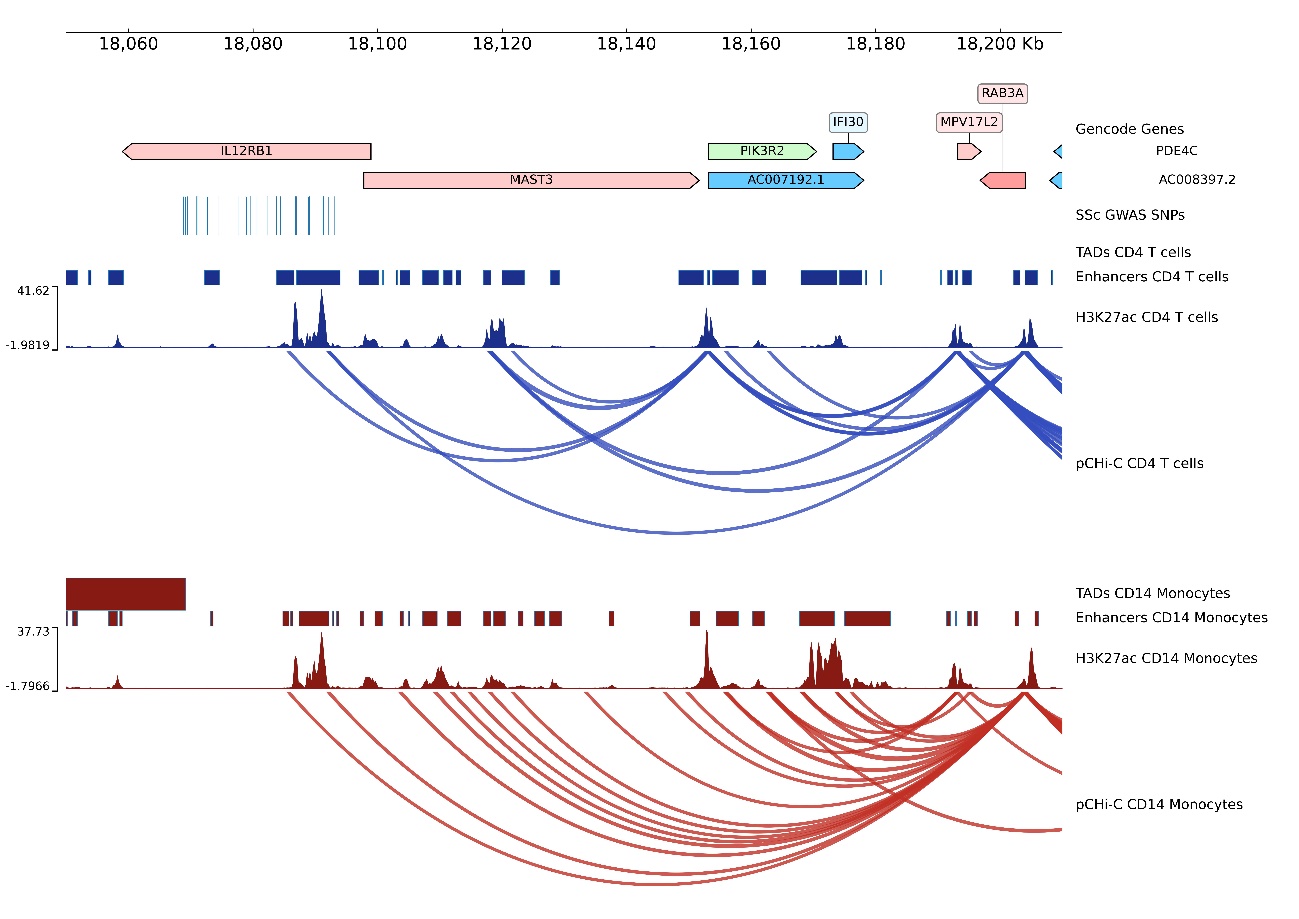

**Supplementary Figure 22.** Promoter capture Hi-C (pCHi-C) interactions in the rs2305743 *(IL12RB1)* locus. Genomic coordinates (GRCh38) are shown at the top of the panel. The tracks include Gencode Genes, systemic sclerosis (SSc)-associated SNPs from López-Isac et al (18) and their proxies (r2>0.8), topologically associating domains (TADs) (shown as bars), H3K27ac signal, enhancer regions as defined by chromHMM, and pCHi-C significant interactions (CHiCAGO score > 5) (shown as arcs) in CD4^+^ T cells (blue) and CD14^+^ monocytes (red).

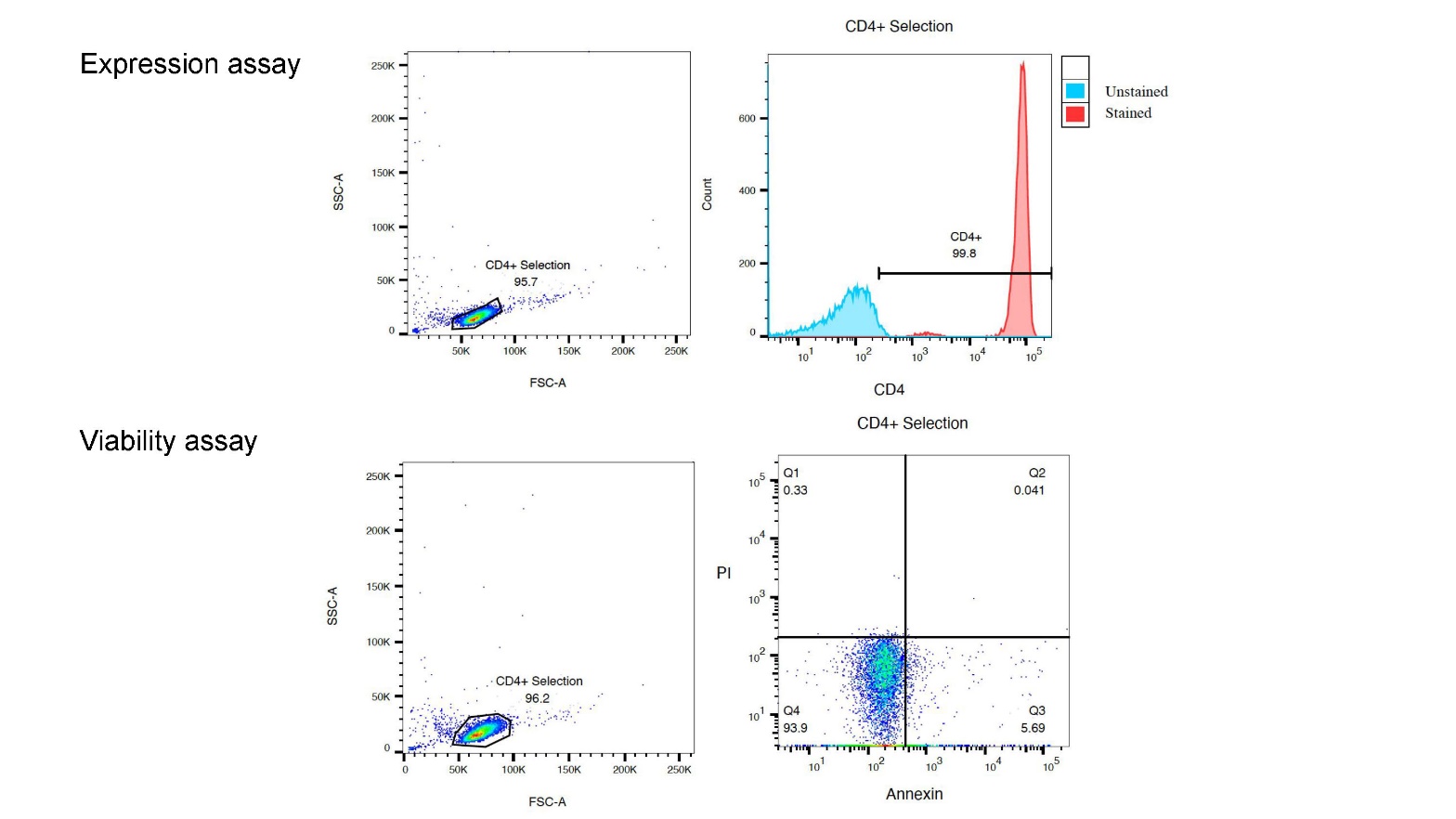

**Supplementary Figure 23.** Flow cytometry verifying the purity and viability of the cell isolation protocol for CD4+ T cells. Purity (determined by expression of CD4) was verified to be 99.8% of the selected cell population, which is 95.7% of the total counts. Viability (determined by Annexin-V and propidium iodide) was verified at 93.9%.

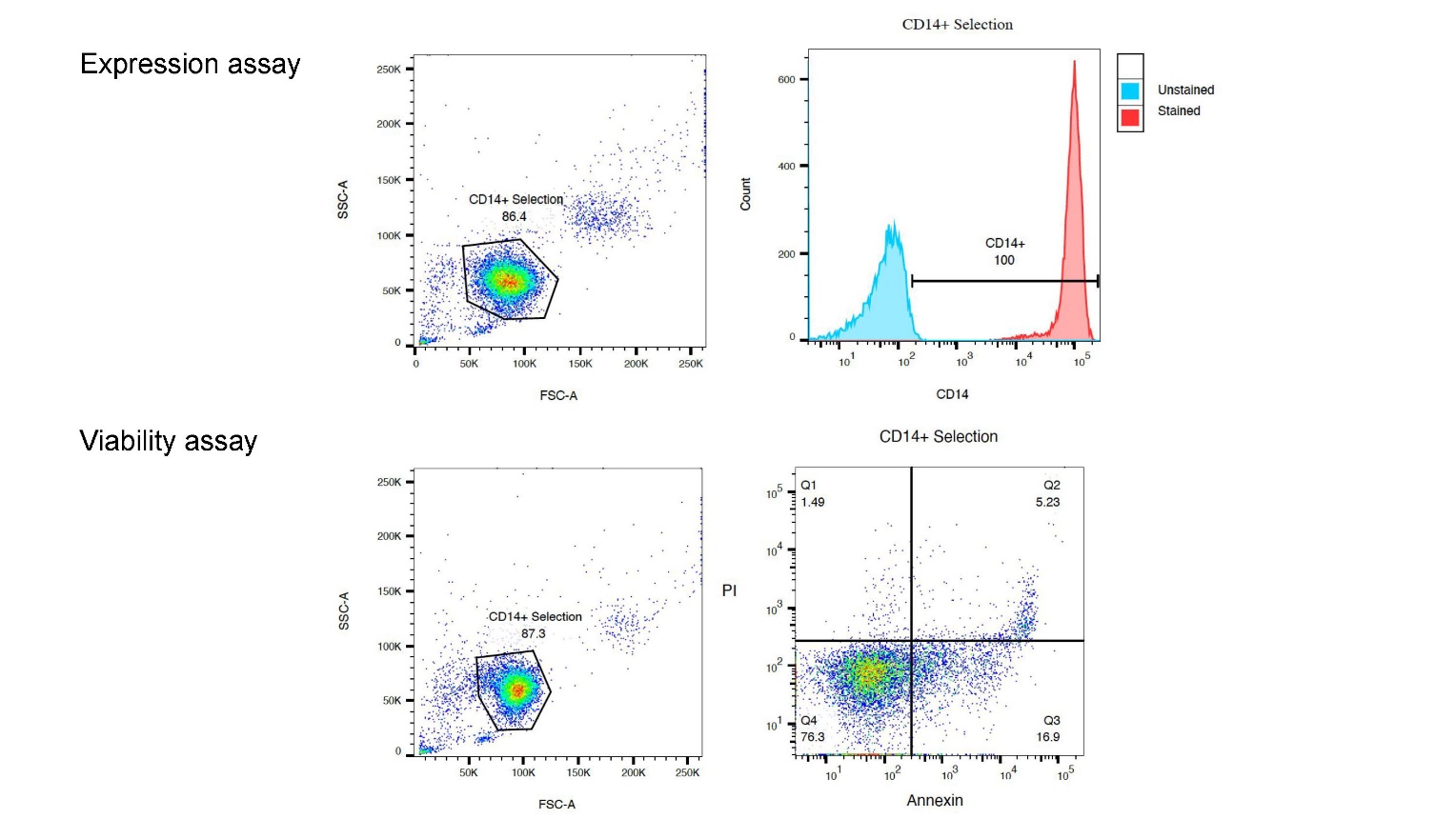

**Supplementary Figure 24.** Flow cytometry verifying the purity and viability of the cell isolation protocol for CD14+ Monocytes. Purity (determined by expression of CD14) was verified to be 100% of the selected cell population, which is 86.5% of the total counts. Viability (determined by Annexin-V and propidium iodide) was verified at 76.3%.

**Supplementary Tables**

**Supplementary Table 1.** Clinical and demographic data of systemic sclerosis patients and healthy controls.

| **Identifier** | **Disease status** | **Disease subtype** | **Age*** | **Sex** |
| --- | --- | --- | --- | --- |
| Patient 1 | SSc patient | Limited | 70-74 | Female |
| Patient 2 | SSc patient | Limited | 55-59 | Female |
| Patient 3 | SSc patient | Limited | 60-64 | Female |
| Patient 4 | SSc patient | Limited | 60-64 | Female |
| Patient 5 | SSc patient | Diffuse | 45-49 | Female |
| Patient 6 | SSc patient | Diffuse | 35-39 | Female |
| Patient 7 | SSc patient | Diffuse | 60-64 | Female |
| Patient 8 | SSc patient | Diffuse | 50-54 | Female |
| Patient 9 | SSc patient | Diffuse | 60-64 | Female |
| Patient 10 | SSc patient | Limited | 60-64 | Female |
| Control 1 | Healthy control | - | 25-29 | Male |
| Control 2 | Healthy control | - | 50-54 | Female |
| Control 3 | Healthy control | - | 50-54 | Female |
| Control 4 | Healthy control | - | 50-54 | Male |
| Control 5 | Healthy control | - | 55-59 | Female |

*Age of individuals when blood extraction was performed

**Supplementary Table 2**. Number of reads after HiCUP alignment and filtering of pCHi-C data.

| **Identifiers** | **Cell type** | **Total read pairs** | **Paired align** | **Valid rate (%)** | **Valid ditags** | **Unique ditags** | **Ditags duplication rate (%)** | **On-target ditags** | **On-target rate (%)** |
| --- | --- | --- | --- | --- | --- | --- | --- | --- | --- |
| Control 1 | CD4+ T cells | 512177179 | 289850386 | 82,72 | 239601506 | 82654878 | 65,5 | 11785577 | 14,25 |
|  | CD14+ monocytes | 598399541 | 407069986 | 75,44 | 306989949 | 58555629 | 80,93 | 33553452 | 57,3 |
| Control 2 | CD4+ T cells | 559118834 | 348499840 | 86,3 | 300595506 | 143426522 | 52,29 | 39474179 | 27,52 |
|  | CD14+ monocytes | 506954077 | 309516812 | 86,26 | 266869550 | 88746751 | 66,75 | 30752664 | 34,65 |
| Control 3 | CD4+ T cells | 463266373 | 282139234 | 84,26 | 237678643 | 95688469 | 59 | 31080028 | 32,48 |
|  | CD14+ monocytes | 483525548 | 285998823 | 86,95 | 248508586 | 123620799 | 50,25 | 36578861 | 29,59 |
| Control 4 | CD4+ T cells | 486749896 | 316379163 | 75,72 | 239415518 | 44815856 | 81,28 | 33944346 | 75,74 |
|  | CD14+ monocytes | 545086196 | 303120064 | 85,11 | 257739933 | 86916088 | 66,28 | 27013886 | 31,08 |
| Control 5 | CD4+ T cells | 505311506 | 348101258 | 71,63 | 249226756 | 32832588 | 86,83 | 27805005 | 84,68 |
|  | CD14+ monocytes | 450094153 | 251750604 | 85,84 | 215892785 | 81679085 | 62,17 | 24476551 | 29,97 |
| Patient 1 | CD4+ T cells | 596899626 | 334134066 | 85,61 | 285896447 | 76754039 | 73,15 | 28889836 | 37,64 |
|  | CD14+ monocytes | 434717431 | 248291417 | 84,83 | 210506596 | 82111642 | 60,99 | 27694919 | 33,73 |
| Patient 2 | CD4+ T cells | 469763189 | 260633656 | 85,46 | 222697560 | 69414218 | 68,83 | 30116005 | 43,38 |
|  | CD14+ monocytes | 460010412 | 253459164 | 84,73 | 212830646 | 46396538 | 78,2 | 20670637 | 44,55 |
| Patient 3 | CD4+ T cells | 484619244 | 266203618 | 84,23 | 223665186 | 51908654 | 76,79 | 25455852 | 49,04 |
|  | CD14+ monocytes | 557578828 | 306711062 | 83,88 | 257078113 | 73555230 | 76,39 | 30411030 | 41,34 |

**Supplementary Table 2**. Number of reads after HiCUP allignment and filtering of pCHi-C data (continuation).

| **Identifiers** | **Cell type** | **Total read pairs** | **Paired align** | **Valid rate (%)** | **Valid ditags** | **Unique ditags** | **Ditags duplication rate (%)** | **On-target ditags** | **On-target rate (%)** |
| --- | --- | --- | --- | --- | --- | --- | --- | --- | --- |
| Patient 4 | CD4+ T cells | 469382219 | 264830498 | 84,3 | 223370947 | 51140806 | 77,1 | 30838017 | 60,3 |
|  | CD14+ monocytes | 457312653 | 256003157 | 86,11 | 220493190 | 81500776 | 63,04 | 40125407 | 49,23 |
| Patient 5 | CD4+ T cells | 487944894 | 264223927 | 85,92 | 227010264 | 56124422 | 75,28 | 22843882 | 40,7 |
|  | CD14+ monocytes | 453928751 | 253398622 | 85,77 | 217236914 | 66179965 | 69,54 | 24938066 | 37,68 |
| Patient 6 | CD4+ T cells | 448621192 | 238163644 | 85,53 | 203602872 | 58129875 | 71,45 | 24449761 | 42,06 |
|  | CD14+ monocytes | 505600799 | 266272198 | 85,16 | 226569823 | 52073905 | 77,02 | 18910095 | 36,31 |
| Patient 7 | CD4+ T cells | 534934259 | 300936296 | 82,52 | 248289838 | 38360709 | 84,55 | 21885688 | 57,05 |
|  | CD14+ monocytes | 478697222 | 277297863 | 84,1 | 233041721 | 33566865 | 85,6 | 19188795 | 57,17 |
| Patient 8 | CD4+ T cells | 556928710 | 326202981 | 86,93 | 283705423 | 76151835 | 73,16 | 36277850 | 47,64 |
|  | CD14+ monocytes | 503474455 | 287223031 | 86,81 | 249346399 | 113587623 | 54,45 | 33538229 | 29,53 |
| Patient 9 | CD4+ T cells | 482366451 | 271674962 | 86,94 | 236198200 | 59733249 | 74,71 | 29862898 | 49,99 |
|  | CD14+ monocytes | 484859062 | 271899096 | 88,52 | 240703905 | 79402715 | 67,01 | 35202959 | 44,33 |
| Patient 10 | CD4+ T cells | 430784477 | 234590464 | 85,28 | 200541235 | 40835308 | 77,5 | 21661328 | 53,05 |
|  | CD14+ monocytes | 482295416 | 266056173 | 82,98 | 220458961 | 39118111 | 81,6 | 21557477 | 55,11 |

**Supplementary Table 3.** Number of reads from RNA-seq data.

| **Identifiers** | **Cell type** | **Read pairs (millions)** | **Unique reads (millions)** | **Duplication rate (%)** |
| --- | --- | --- | --- | --- |
| Control 2 | CD4+ T cells | 20,6 | 2,1 | 87,99 |
|  | CD14+ monocytes | 25,15 | 17,1 | 39,76 |
| Control 3 | CD4+ T cells | 29,52 | 4,87 | 80,44 |
|  | CD14+ monocytes | 24,97 | 18,26 | 37,63 |
| Control 4 | CD4+ T cells | 33,67 | 3,38 | 87,36 |
|  | CD14+ monocytes | 3,18 | 2,6 | 10,71 |
| Control 5 | CD4+ T cells | 37,23 | 3,51 | 88,00 |
|  | CD14+ monocytes | 43,37 | 21,44 | 48,95 |
| Patient 1 | CD4+ T cells | 40,12 | 5,59 | 82,93 |
|  | CD14+ monocytes | 37,03 | 16,59 | 52,37 |
| Patient 2 | CD4+ T cells | 33,18 | 2,78 | 89,23 |
|  | CD14+ monocytes | 35,17 | 2,22 | 91,36 |
| Patient 3 | CD4+ T cells | 28,28 | 2,52 | 88,67 |
|  | CD14+ monocytes | 36,81 | 8,04 | 74,23 |
| Patient 4 | CD4+ T cells | 57,28 | 9,84 | 78,77 |
|  | CD14+ monocytes | 31,84 | 3,29 | 87,05 |
| Patient 5 | CD4+ T cells | 35,37 | 15,24 | 53,80 |
|  | CD14+ monocytes | 37,53 | 16,6 | 52,52 |
| Patient 6 | CD4+ T cells | 38,79 | 5,2 | 83,42 |
|  | CD14+ monocytes | 37,22 | 9,85 | 68,80 |
| Patient 7 | CD4+ T cells | 36,38 | 3,05 | 89,01 |
|  | CD14+ monocytes | 36,23 | 16,24 | 53,17 |
| Patient 8 | CD4+ T cells | 20,04 | 0,85 | 93,60 |
|  | CD14+ monocytes | 35,35 | 8,98 | 70,72 |
| Patient 9 | CD4+ T cells | 27,97 | 1,27 | 93,24 |
|  | CD14+ monocytes | 32,66 | 3,57 | 86,49 |
| Patient 10 | CD4+ T cells | 29,5 | 2,32 | 89,61 |
|  | CD14+ monocytes | 32,37 | 17,42 | 46,70 |

**Supplementary Table 4.** Number of SSc GWAS SNPs and significant interactions in each locus.

| **Locus** | | | **Chr** | | **Start (bp)** | | **End (bp)** | | **N SSc GWAS SNPs** | | **N SSc GWAS SNPs + enhancer overlap** | | | |  | | **N significant interactions (SSc GWAS SNPs + Enhancer overlap)** | | |
| --- | --- | --- | --- | --- | --- | --- | --- | --- | --- | --- | --- | --- | --- | --- | --- | --- | --- | --- | --- |
|  |  |  |  |  |  |  |  |  |  |  | **CD4+** | | **CD14+** | |  | | **CD4+** | | **CD14+** |
| 1 | | | 1 | | 67326053 | | 67448804 | | 27 | | 10 | | 1 | |  | | 0 | | 0 |
| 2 | | | 1 | | 167445635 | | 167465040 | | 20 | | 20 | | 11 | |  | | 28 | | 0 |
| 3 | | | 1 | | 173337507 | | 173391947 | | 99 | | 3 | | 2 | |  | | 0 | | 0 |
| 4 | | | 2 | | 190642047 | | 190698201 | | 28 | | 18 | | 5 | |  | | 31 | | 4 |
| 5 | | | 2 | | 191035723 | | 191108308 | | 30 | | 21 | | 2 | |  | | 14 | | 0 |
| 6 | | | 3 | | 58084620 | | 58482701 | | 157 | | 17 | | 37 | |  | | 2 | | 19 |
| 7 | | | 3 | | 119384733 | | 119546340 | | 28 | | 1 | | 0 | |  | | 1 | | 0 |
| 8 | | | 3 | | 160002484 | | 160030580 | | 54 | | 14 | | 13 | |  | | 0 | | 13 |
| 9 | | | 4 | | 960523 | | 990021 | | 12 | | 5 | | 4 | |  | | 6 | | 6 |
| 10 | | | 4 | | 102477892 | | 102615256 | | 119 | | 97 | | 16 | |  | | 108 | | 10 |
| 11 | | | 5 | | 151064651 | | 151080486 | | 17 | | 6 | | 8 | |  | | 0 | | 0 |
| 12 | | | 6 | | 106181815 | | 106339294 | | 59 | | 14 | | 1 | |  | | 0 | | 0 |
| 13 | | | 7 | | 128933913 | | 129095960 | | 128 | | 26 | | 35 | |  | | 0 | | 0 |
| 14 | | | 8 | | 11474517 | | 11544554 | | 42 | | 13 | | 13 | |  | | 0 | | 0 |
| 15 | | | 8 | | 60638547 | | 60664239 | | 11 | | 1 | | 0 | |  | | 3 | | 0 |
| 16 | | | 11 | | 554659 | | 619789 | | 22 | | 9 | | 6 | |  | | 0 | | 0 |
| 17 | | | 11 | | 2311894 | | 2363262 | | 80 | | 3 | | 0 | |  | | 3 | | 0 |
| 18 | | | 11 | | 118704617 | | 118875175 | | 120 | | 31 | | 24 | |  | | 47 | | 4 |
| 19 | | | 15 | | 74739180 | | 75148328 | | 216 | | 68 | | 54 | |  | | 133 | | 38 |
| 20 | | | 16 | | 85932852 | | 85979945 | | 46 | | 21 | | 27 | |  | | 0 | | 12 |
| 21 | | | 17 | | 39747478 | | 39933464 | | 104 | | 22 | | 12 | |  | | 15 | | 1 |
| 22 | | | 17 | | 75193533 | | 75279345 | | 61 | | 16 | | 9 | |  | | 0 | | 0 |
| 23 | | | 19 | | 18068862 | | 18093031 | | 25 | | 9 | | 5 | |  | | 7 | | 2 |
|  |  |  | |  | |  | | 1505 | | 445 | | 285 | |  | | 398 | | 109 | |

| **Supplementary Table 5.** Protein-Protein interaction (PPI) network from genes with significant interactions in CD4^+^ T cells and CD14^+^ monocytes.  All PPI are calculated with STRINGS software, only those with highest confidence score (>0.9) were taking into account.  PPI within core genes are highlighted in bold. | |  |
| --- | --- | --- |
| **Genes** | **STRINGS PPI (>0.9 score)** | **N** |
| CD247 | B2M; CBL; CD2; CD28; CD3D; CD3E; CD3G; CD4; CD80; CD86; CD8A; CD8B; **CSK**; DBNL; FYB; FYN; GNAI1; GNAI2; GNAI3; GNAO1; GNAZ; GNB1; GNG2; GRAP2; HLA-A; HLA-DRA; HLA-DRB1; IL2; IL2RA; IL2RB; IL2RG; ITK; JUN; LAT; LCK; LCP2; MAP4K1; PAG1; PIK3CA; PRKACB; PRKACG; PRKAR1A; PRKAR1B; PRKAR2A; PRKAR2B; PRKCQ; PTPN11; PTPN6; PTPRC; RGS1; SH3BP2; SHC1; SLA2; SYK; TRAT1; VAV1; ZAP70 | 57 |
| CREG1 | ACLY; ACTR10; ACTR2; AGA; ANXA2; ARG1; ARSA; ARSB; AZU1; BPI; C3; C6orf120; CAP1; CCT2; CCT8; CECR1; CPPED1; CTSA; CTSC; CTSG; CYB5R3; DNAJC3; DPP7; DSN1; DYNC1H1; ELANE; FABP5; FAF2; FRK; FTL; FUCA1; FUCA2; GALNS; GCA; GDI2; GGH; GLA; GLB1; GM2A; GNS; GRN; GUSB; HEBP2; HEXB; HMHA1; HRNR; IMPDH1; IST1; LYZ; MAN2B1; MAPK1; MNDA; MPO; NAPRT; NHLRC3; NPC2; ORM2; PA2G4; PADI2; PLAC8; PRDX6; PRKCD; PRSS3P2; PRTN3; PSMD1; PTGES2; PYCARD; PYGB; RETN; RNASE2; RNASE3; RNASET2; S100A7; **SDCBP**; SERPINB3; STK11IP; TOLLIP; TRAPPC1; TTR; TUBB; TUBB4B; TXNDC5; UNC13D; VAT1; VCP | 85 |
| MFSD6 | MFSD12; **NEMP2**; ZNF384 | 3 |
| NEMP2 (TMEM194B) | C2orf88; **MFSD6** | 2 |
| HIBCH | ECHS1; EHHADH; HADH; HADHA; HIBADH | 5 |
| INPP1 | IMPA1; IMPA2; IMPAD1; INPP4A; INPP4B; INPP5A; INPP5B; INPP5F; INPP5J; INPP5K; INPPL1; ITPK1; OCRL; SYNJ1; SYNJ2 | 15 |
| STAT4 | AIP; ANXA2; ARF1; CA1; CAPZA1; CDC42; CFL1; CISH; CNN2; CREBBP; EBI3; ETV5; GSTA2; GSTO1; HLX; HNRNPA2B1; HNRNPDL; HNRNPF; HSPA9; IFNG; IFNL1; IFNLR1; IL10; IL10RB; IL12A; IL12B; IL12RB1; IL12RB2; IL18; IL18R1; IL18RAP; IL21; IL21R; IL23A; IL23R; IL27; IL27RA; IL2RG; IL6ST; JAK1; JAK2; JAK3; JUN; LCP1; LMNB1; MAPK14; MIF; MSN; MTAP; **NFKB1**; PAK2; PDCD4; PIAS2; PITPNA; PPIA; PSME2; RALA; RAP1B; RELA; RIPK2; RPLP0; SERPINB2; SNRPA1; SOD1; SOD2; SPHK2; STAT1; STAT2; STAT3; STAT5A; STAT5B; TALDO1; TCP1; TYK2 | 74 |
| NABP1 | ASUN; CCNK; CCNT1; CCNT2; CDK7; CDK9; CPSF3L; ELL; ELL2; ELL3; GTF2A1; GTF2A2; GTF2B; GTF2E1; GTF2E2; GTF2F1; GTF2F2; ICE1; ICE2; INIP; INTS1; INTS10; INTS12; INTS2; INTS3; INTS4; INTS5; INTS6; INTS7; INTS8; INTS9; NABP2; NCBP1; NCBP2; PCF11; PHAX; POLR2A; POLR2B; POLR2C; POLR2D; POLR2E; POLR2F; POLR2G; POLR2H; POLR2I; POLR2J; POLR2K; POLR2L; POU2F1; POU2F2; RPAP2; RPRD1A; RPRD1B; RPRD2; SNAPC1; SNAPC2; SNAPC3; SNAPC4; SNAPC5; SP1; SRRT; SSU72; SUPT4H1; SUPT5H; TAF11; TAF13; TAF5; TAF6; TAF8; TBP; VWA9; ZC3H8; ZNF143 | 73 |
| RPP14 | AGO2; BMS1; BYSL; CIRH1A; CLP1; CSNK1D; CSNK1E; DCAF13; DDX47; DDX49; DDX52; DHX37; DICER1; DIEXF; EMG1; FAU; FBL; FCF1; HEATR1; HSD17B10; IMP3; IMP4; KIAA0391; KRR1; LTV1; MPHOSPH10; NHP2L1; NOB1; NOC4L; NOL11; NOL6; NOP14; NOP56; NOP58; PDCD11; PNO1; POP1; POP4; POP5; POP7; PWP2; **PXK**; RCL1; RIOK1; RIOK2; RIOK3; RPP21; RPP25; RPP30; RPP38; RPP40; RPS10; RPS11; RPS12; RPS13; RPS14; RPS15; RPS15A; RPS16; RPS17; RPS18; RPS19; RPS2; RPS20; RPS21; RPS23; RPS24; RPS25; RPS26; RPS27; RPS27A; RPS27L; RPS28; RPS29; RPS3; RPS3A; RPS4X; RPS5; RPS6; RPS7; RPS8; RPS9; RPSA; RRP36; RRP7A; RRP9; TARBP2; TBL3; TSR1; UTP11L; UTP14A; UTP14C; UTP15; UTP18; UTP20; UTP3; UTP6; WBSCR22; WDR3; WDR36; WDR43; WDR46; WDR75 | 103 |
| KCTD6 | ANAPC1; ANAPC2; ANAPC5; ANKRD9; AREL1; ASB1; ASB14; ASB16; ASB17; ASB4; ASB5; ASB6; ASB7; ASB8; ATG7; BTBD1; BTRC; CBLB; CDC20; CDC23; CDC34; COMMD1; COMMD10; COMMD3; COMMD4; COMMD6; COMMD7; COMMD8; COMMD9; COPS2; COPS3; COPS4; COPS6; COPS7A; COPS7B; COPS8; CUL1; CUL3; DCUN1D1; DCUN1D3; DCUN1D5; DTX3L; FBXL12; FBXL13; FBXL14; FBXL15; FBXL19; FBXL20; FBXL3; FBXL4; FBXL5; FBXL8; FBXO10; FBXO11; FBXO17; FBXO2; FBXO21; FBXO22; FBXO27; FBXO30; FBXO31; FBXO32; FBXO4; FBXO40; FBXO7; FBXO9; FBXW11; FBXW12; FBXW2; FBXW5; FBXW7; FBXW8; FEM1A; FEM1B; FEM1C; FZR1; GLMN; HACE1; HECTD1; HECTD2; HECTD3; HECW2; HERC2; HERC3; HERC4; HERC5; HERC6; HUWE1; ITCH; KCTD7; KEAP1; KLHL11; KLHL20; KLHL22; KLHL25; KLHL3; KLHL41; LMO7; LNX1; LONRF1; LRR1; LRSAM1; MGRN1; MKRN1; NEDD4; NEDD4L; NEDD8; RBBP6; RBX1; RCHY1; RLIM; RNF111; RNF114; RNF123; RNF126; RNF138; RNF144B; RNF220; RNF25; RNF41; RNF6; RNF7; RPS27A; RUNX1; SH3RF1; SIAH1; SIAH2; SKP1; SKP2; SMURF1; SMURF2; SOCS1; SOCS3; SOCS5; SPSB1; SPSB4; STUB1; TCEB2; TRAF7; TRAIP; TRIM11; TRIM21; TRIM36; TRIM37; TRIM4; TRIM41; TRIM50; TRIM9; TRIP12; TULP4; UBA1; UBA3; UBA5; UBA52; UBA6; UBA7; UBB; UBC; UBE2A; UBE2B; UBE2C; UBE2D1; UBE2D2; **UBE2D3**; UBE2D4; UBE2E1; UBE2E2; UBE2E3; UBE2G1; UBE2G2; UBE2H; UBE2J1; UBE2K; UBE2L6; UBE2M; UBE2N; UBE2O; UBE2Q1; UBE2Q2; UBE2R2; UBE2S; UBE2U; UBE2V1; UBE2W; UBE2Z; UBE3A; UBE3B; UBE3C; UBE4A; UBOX5; UBR1; UNKL; VHL; WSB1; WSB2; WWP1; ZBTB16; ZNF645; ZNRF1; ZNRF2 | 200 |
| PXK | PHRF1; **RPP14** | 2 |
| TMEM39A | NA | 0 |
| POGLUT1 | NOTCH1; NOTCH2; NOTCH3; NOTCH4 | 4 |
| SMC4 | ASPM; AURKB; BUB1; BUB1B; CCNB1; CCNB2; CDC5L; CDC6; CDK1; CENPE; CSNK2A1; CSNK2A2; CSNK2B; ESCO2; ESPL1; H2AFB1; H2AFJ; H2AFV; H2AFX; H2AFZ; H2BFS; HIST1H2AC; HIST1H2AD; HIST1H2AJ; HIST1H2BA; HIST1H2BB; HIST1H2BD; HIST1H2BH; HIST1H2BJ; HIST1H2BK; HIST1H2BL; HIST1H2BM; HIST1H2BN; HIST1H2BO; HIST2H2AC; HIST2H2BE; HIST3H3; KIF11; KIF15; MAD2L1; MCM2; MCM3; MCM4; MCM5; MCM7; MCPH1; MKI67; NCAPD2; NCAPD3; NCAPG; NCAPG2; NCAPH; NCAPH2; NDC80; NIPBL; NUF2; PDS5A; PLK1; POLA1; RAD21; RB1; RFC1; RRM1; SET; SMC1A; SMC2; SMC3; SMC5; SMC6; TOP2A; TTK | 71 |
| IFT80 | CLUAP1; DYNC2H1; DYNC2LI1; DYNLL1; DYNLL2; DYNLRB1; DYNLRB2; HSPB11; IFT122; IFT140; IFT172; IFT20; IFT22; IFT27; IFT43; **IFT46**; IFT52; IFT57; IFT74; IFT81; IFT88; KIF17; KIF3A; KIF3B; KIF3C; KIFAP3; TCTE3; TCTEX1D1; TCTEX1D2; TRAF3IP1; TTC21B; TTC26; TTC30A; TTC30B; WDR19; WDR34; WDR35; WDR60 | 38 |
| GAK | AAK1; ACTR2; ACTR3; ADRB2; AGFG1; AGTR1; AMPH; AP1B1; AP1G1; AP1M1; AP1M2; AP1S1; AP1S2; AP1S3; AP2A1; AP2A2; AP2B1; AP2M1; AP2S1; AP3B1; AP3S1; AP4B1; AP4E1; APOB; ARF1; ARPC1A; ARPC2; ARPC3; ARPC4; ARPC5; ARRB1; ARRB2; AVP; AVPR2; BIN1; BLOC1S3; BLOC1S4; BLOC1S6; CBL; CD3D; CD3G; CD4; CFTR; CHRM2; CLHC1; CLINT1; CLTA; CLTB; CLTC; CLTCL1; CPD; CTTN; DAB2; DNAJC6; DNM1; DNM2; DNM3; DTNBP1; DVL2; EGF; EGFR; EPN1; EPN2; EPS15; EPS15L1; FNBP1; FNBP1L; FTH1; FTL; FZD4; GAPVD1; HGS; HIP1; HIP1R; HSPA8; HSPA9; IGF2R; IL7R; ITSN1; ITSN2; KIAA0319; LDLR; LDLRAP1; LRP2; M6PR; NAPA; NECAP1; NECAP2; OCRL; PACSIN1; PACSIN2; PACSIN3; PICALM; PIK3C2A; PUM1; RAB5A; RAB5B; RAB5C; REPS1; REPS2; RPS27A; SCARB2; SGIP1; SH3D19; SH3GL1; SH3GL2; SH3GL3; SH3KBP1; SLC18A3; SLC2A8; SNAP91; SNAPIN; SNX18; SNX2; SNX5; SNX9; SORT1; STAM; STAM2; STON1; STON2; SYNJ1; SYNJ2; SYT1; SYT11; SYT2; SYT8; SYT9; TACR1; TBC1D8B; TF; TFRC; TGOLN2; TPD52; TPD52L1; TRIP10; TXNDC5; UBA52; UBB; UBC; VAMP2; VAMP3; VAMP4; VAMP7; VAMP8; VPS35; WASL; WNT5A; YIPF6 | 149 |
| TMEM175 | NA | 0 |
| FGFRL1 | FGF10; FGF18; FGF2; FGF22; FGF23; FGF3; FGF4; SPRED1; SPRED2 | 9 |
| SLC39A8 | SLC11A2; SLC30A1; SLC30A10; SLC30A2; SLC30A4; SLC30A5; SLC30A6; SLC30A7; SLC39A1; SLC39A11; SLC39A14; SLC39A2; SLC39A3; SLC39A9 | 14 |
| NFKB1 | ACAA1; AGER; AKT1; ANAPC10; ANAPC11; ANAPC2; APP; ARG1; B2M; BCL10; BCL2A1; BCL3; BIRC3; BTRC; CARD11; CASP8; CD40; CD40LG; CDC23; CDC27; CDC34; CEBPB; CHUK; CREBBP; CTSD; CUL1; CXCL1; CXCL8; DDX58; DOCK2; EP300; EPO; EPOR; ERC1; ERP44; FBXW11; FRK; FZR1; HDAC2; HDAC3; HMGB1; HP; HRAS; IFIH1; IKBKB; IKBKG; IL12B; IL12RB1; IL18; IL18R1; IL1A; IL1B; IL1R1; IL23A; IL23R; IL6; IRAK1; IRAK2; ITGAM; ITGB2; JAK2; JUP; KPNB1; KRAS; LCK; LCN2; LRRC7; LYN; LYZ; MALT1; MAP2K1; MAP2K4; MAP3K1; MAP3K7; MAP3K8; MAPK1; MAPK14; MAPK3; MAPK8; MAVS; MTOR; MTPN; MYD88; NFKB2; NFKBIA; NFKBIB; NFKBIE; NFKBIZ; NGF; NKIRAS1; NKIRAS2; NLRP3; NR3C1; NRAS; OLFM4; PDPK1; PGLYRP1; PIK3CA; PIK3R1; PPARG; PRKACA; PRKACB; PRKACG; PRKCA; PRKCD; PRKCI; PRKCZ; PSMA1; PSMA2; PSMA3; PSMA4; PSMA5; PSMA6; PSMA7; PSMA8; PSMB1; PSMB10; PSMB11; PSMB2; PSMB3; PSMB4; PSMB5; PSMB6; PSMB7; PSMB8; PSMB9; PSMC1; PSMC2; PSMC3; PSMC4; PSMC5; PSMC6; PSMD1; PSMD10; PSMD11; PSMD12; PSMD13; PSMD14; PSMD2; **PSMD3**; PSMD4; PSMD6; PSMD7; PSMD8; PSMD9; PSME2; PSME4; PTPN6; RAF1; RAN; RBX1; REL; RELA; RELB; RIPK1; RIPK3; RPS27A; RPS6KA5; S100A12; S100B; SAA1; SETD6; SIRT1; SKP1; SPTAN1; SQSTM1; SRC; **STAT4**; SYK; TAB1; TAB2; TAB3; TBK1; TERT; TIMP2; TLR2; TLR4; TNF; TNFRSF1A; TNIP2; TOLLIP; TRADD; TRAF1; TRAF2; TRAF6; TRIM25; TXN; TYK2; UBA52; UBB; UBC; UBE2D1; UBE2D2; UBE2E1; UBE2N; UBE2V1; VCL; XPO1; YWHAE; ZBP1 | 200 |
| UBE2D3 | AGXT; ANAPC10; ANAPC11; ANAPC13; ANAPC2; ANAPC4; ANAPC5; AREL1; ARIH2; ASB1; ASB12; ASB14; ASB6; ATG7; BARD1; BCL10; BIRC2; BIRC3; BRCA1; BTBD1; BTRC; CBL; CBLB; CCNF; CDC16; CDC20; CDC23; CDC26; CDC27; CDC34; CUL1; CUL2; CUL3; CUL5; DCUN1D1; DDX58; DET1; DTX3L; DZIP3; FBXL12; FBXL13; FBXL14; FBXL15; FBXL16; FBXL19; FBXL22; FBXL3; FBXL4; FBXL5; FBXL7; FBXO2; FBXO32; FBXO7; FBXO9; FBXW11; FBXW8; FZR1; GAN; HACE1; HECTD1; HECTD2; HECTD3; HECW2; HERC1; HERC2; HERC3; HERC4; HERC5; HERC6; HIF1A; HMGCL; HUWE1; IKBKG; ITCH; KBTBD8; **KCTD6**; KCTD7; KEAP1; KLHL13; KLHL2; KLHL20; KLHL21; KLHL22; KLHL25; KLHL3; KLHL41; KLHL5; KLHL9; LNX1; LRSAM1; LTN1; MGRN1; MIB2; MYLIP; NEDD4; NEDD4L; NEDD8; PAOX; PARK2; PEX1; PEX10; PEX12; PEX13; PEX14; PEX2; PEX6; PJA1; PJA2; RBCK1; RBX1; RCHY1; RIPK1; RLIM; RNF111; RNF114; RNF115; RNF125; RNF126; RNF130; RNF138; RNF14; RNF144B; RNF181; RNF19A; RNF19B; RNF217; RNF220; RNF25; RNF4; RNF41; RNF6; RNF7; RPS27A; SH3RF1; SIAH1; SIAH2; SKP1; SMAD1; SMAD2; SMAD3; SMAD4; SMAD5; SMAD9; SMURF1; SMURF2; STUB1; TLR3; TLR4; TRAF7; TRAIP; TRIM11; TRIM21; TRIM32; TRIM37; TRIM39; TRIM50; TRIM63; TRIM69; TRIM71; TRIP12; UBA1; UBA3; UBA5; UBA52; UBA6; UBA7; UBB; UBC; UBE2A; UBE2B; UBE2C; UBE2F; UBE2G1; UBE2G2; UBE2H; UBE2J2; UBE2K; UBE2M; UBE2N; UBE2O; UBE2R2; UBE2S; UBE2U; UBE2V1; UBE2V2; UBE3A; UBE3B; UBE3C; UBE3D; UBE4A; UBOX5; UBR1; UBR2; UBXN7; UFL1; WSB1; WWP1; ZFAND6; ZNRF1; ZNRF2 | 200 |
| CISD2 | NA | 0 |
| SLC9B1 | NA | 0 |
| BDH2 | AACS; HMGCL; HMGCLL1; OXCT1; OXCT2 | 5 |
| ASPH | ARNT; COPS5; CREB1; EP300; FKBP1B; HIF1A; JUN; RYR1; RYR2; RYR3; TRDN | 11 |
| SDCBP | ACLY; ACTR10; ACTR2; AGA; ANXA2; ARG1; ARSA; ARSB; AZU1; BPI; C3; C6orf120; CAP1; CCT2; CCT8; CECR1; CPPED1; **CREG1**; CTSA; CTSC; CTSG; CYB5R3; DNAJC3; DPP7; DSN1; DYNC1H1; EFNB1; EFNB2; EFNB3; ELANE; EPHB1; EPHB2; EPHB3; EPHB4; EPHB6; FABP5; FAF2; FRK; FTL; FUCA1; FUCA2; GALNS; GCA; GDI2; GGH; GLA; GLB1; GM2A; GNS; GRN; GUSB; HEBP2; HEXB; HMHA1; HRNR; IL5; IL5RA; IMPDH1; IST1; LYZ; MAN2B1; MAPK1; MNDA; MPO; NAPRT; NFASC; NHLRC3; NPC2; ORM2; PA2G4; PADI2; PLAC8; PRDX6; PRKCD; PRSS3P2; PRTN3; PSMD1; PTGES2; PYCARD; PYGB; RETN; RNASE2; RNASE3; RNASET2; S100A7; SDC1; SDC2; SDC4; SERPINB3; STK11IP; TOLLIP; TRAPPC1; TTR; TUBB; TUBB4B; TXNDC5; UNC13D; VAT1; VCP | 99 |
| CHD7 | NLK; PPARG; SETDB1 | 3 |
| TSSC4 | NA | 0 |
| CXCR5 | ACKR3; ADCY1; ADCY2; ADCY3; ADCY4; ADCY5; ADCY6; ADCY7; ADCY8; ADCY9; ADORA1; ADORA3; ADRA2A; ADRA2C; AGT; AGTR2; ANXA1; APLN; APP; BDKRB1; C3; C3AR1; C5; C5AR1; CASR; CCL1; CCL13; CCL16; CCL19; CCL20; CCL21; CCL25; CCL27; CCL28; CCL4; CCL4L1; CCL5; CCR1; CCR10; CCR2; CCR3; CCR4; CCR5; CCR7; CCR8; CCR9; CHRM2; CHRM4; CNR1; CNR2; CORT; CX3CL1; CX3CR1; CXCL1; CXCL10; CXCL11; CXCL12; CXCL13; CXCL16; CXCL2; CXCL3; CXCL5; CXCL6; CXCL8; CXCL9; CXCR1; CXCR2; CXCR3; CXCR4; CXCR6; DRD2; DRD3; DRD4; FPR3; GABBR1; GABBR2; GAL; GALR1; GALR2; GALR3; GNAI1; GNAI2; GNAI3; GNAT3; GNB1; GNB2; GNB3; GNB4; GNB5; GNG10; GNG11; GNG12; GNG13; GNG2; GNG3; GNG4; GNG5; GNG7; GNG8; GNGT1; GNGT2; GPER1; GPR18; GPR183; GPR31; GPR37; GPR37L1; GPR55; GPSM2; GPSM3; GRM2; GRM3; GRM4; GRM6; GRM7; GRM8; HCAR1; HCAR2; HCAR3; HEBP1; HRH3; HRH4; HTR1A; HTR1B; HTR1D; HTR1E; HTR1F; HTR5A; INSL5; KNG1; LPAR1; LPAR5; MCHR1; MCHR2; MTNR1A; MTNR1B; NMS; NMU; NMUR1; NMUR2; NPB; NPBWR1; NPBWR2; NPW; NPY; NPY1R; NPY2R; NPY4R; OPRD1; OPRK1; OPRL1; OXER1; OXGR1; P2RY12; P2RY13; P2RY14; P2RY4; PCP2; PENK; PF4; PMCH; PNOC; POMC; PPBP; PSAP; PTGDR2; PTGER3; PYY; RXFP3; RXFP4; S1PR1; S1PR3; S1PR4; S1PR5; SAA1; SST; SSTR1; SSTR2; SSTR4; SSTR5; SUCNR1; TAS1R1; TAS1R2; TAS1R3; TAS2R1; TAS2R10; TAS2R13; TAS2R16; TAS2R19; TAS2R3; TAS2R31; TAS2R4; TAS2R40; TAS2R41; TAS2R42; TAS2R5; TAS2R60; TAS2R7; TAS2R8; TAS2R9 | 200 |
| UPK2 | UPK1A; UPK1B; UPK3A | 3 |
| DDX6 | AGO1; AGO2; AGO3; AGO4; CNOT1; CNOT4; DCP1A; DCP1B; DCP2; EDC3; EDC4; EIF4E; EIF4ENIF1; LIMD1; LSM1; LSM14A; LSM14B; LSM2; LSM3; LSM4; LSM5; LSM6; LSM7; PATL1; PATL2; XRN1 | 26 |
| IFT46 | ARL13B; CLUAP1; DYNC2H1; DYNC2LI1; DYNLL1; DYNLL2; DYNLRB1; DYNLRB2; HSPB11; IFT122; IFT140; IFT172; IFT20; IFT22; IFT27; IFT43; IFT52; IFT57; IFT74; **IFT80**; IFT81; IFT88; KIF17; KIF3A; KIF3B; KIF3C; KIFAP3; TCTE3; TCTEX1D1; TCTEX1D2; TRAF3IP1; TTC21B; TTC26; TTC30A; TTC30B; WDR19; WDR34; WDR35; WDR60 | 39 |
| ARCN1 | ACTR10; ACTR1A; ANK1; ANK2; ANK3; ARF1; ARF3; ARF4; ARF5; ARFGAP1; ARFGAP2; ARFGAP3; BET1; BET1L; BNIP1; CAPZA1; CAPZA2; CAPZA3; CAPZB; CD55; CD59; CDC5L; CENPE; COG1; COG2; COG3; COG4; COG5; COG6; COG7; COG8; COPA; COPB1; COPB2; COPE; COPG1; COPG2; COPZ1; COPZ2; DCTN1; DCTN2; DCTN3; DCTN4; DCTN5; DCTN6; DYNC1H1; DYNC1I1; DYNC1I2; DYNC1LI1; DYNC1LI2; DYNLL1; DYNLL2; FOLR1; GBF1; GOLGA2; GOLGB1; GORASP1; GOSR1; GOSR2; INS; KDELR1; KDELR2; KDELR3; KIF11; KIF12; KIF13B; KIF15; KIF16B; KIF18A; KIF18B; KIF19; KIF1A; KIF1B; KIF1C; KIF20A; KIF20B; KIF21A; KIF21B; KIF22; KIF23; KIF25; KIF26A; KIF26B; KIF27; KIF2A; KIF2B; KIF2C; KIF3A; KIF3B; KIF3C; KIF4A; KIF4B; KIF5A; KIF5B; KIF6; KIF9; KIFAP3; KIFC1; KIFC2; KLC1; KLC2; KLC3; KLC4; NAPA; NAPB; NAPG; NBAS; NSF; RAB1A; RAB1B; RACGAP1; RINT1; SEC22B; SPTA1; SPTAN1; SPTB; SPTBN1; SPTBN2; SPTBN4; SPTBN5; STX18; STX5; SURF4; TMED10; TMED2; TMED3; TMED7; TMED9; TMEM115; USE1; USO1; YKT6; ZW10 | 133 |
| BCL9L | APC; ASH2L; BCL9; BTRC; CTBP1; CTNNB1; H2AFB1; H2AFJ; H2AFV; H2AFX; H2AFZ; H2BFS; HDAC1; HIST1H2AC; HIST1H2AD; HIST1H2AJ; HIST1H2BA; HIST1H2BB; HIST1H2BD; HIST1H2BH; HIST1H2BJ; HIST1H2BK; HIST1H2BL; HIST1H2BM; HIST1H2BN; HIST1H2BO; HIST2H2AC; HIST2H2BE; HIST3H3; KMT2D; LDB1; LEF1; MEN1; PYGO1; PYGO2; RBBP5; TCF4; TCF7; TCF7L1; TCF7L2; TLE1; TLE2; TLE3; TLE4 | 44 |
| CSK | ACTA1; ACTN1; ACTN2; ACTN3; ACTN4; AGGF1; AGK; AGTRAP; AKAP9; AP3B1; APBB1IP; ARAF; ARRB1; ARRB2; ATG7; B2M; BCAR1; BCL2L11; BCL6; BCR; BLK; BRAF; CAPN1; **CD247**; CD274; CD3D; CD3E; CD3G; CD4; CD79A; CD79B; CD8A; CD8B; CLCN6; CNKSR1; CNKSR2; CREBBP; CRKL; DOK1; EGF; EGFR; ESRP1; FAM114A2; FAM131B; FCGR2B; FGA; FGB; FGG; FGR; FLT1; FLT4; FN1; FXR1; FYN; GAB1; HCK; HLA-A; HLA-DPA1; HLA-DPB1; HLA-DQA1; HLA-DQA2; HLA-DQB1; HLA-DQB2; HLA-DRA; HLA-DRB1; HLA-DRB5; HRAS; IL17RD; IQGAP1; ITGA1; ITGA2B; ITGB1; ITGB3; KDM7A; KDR; KIAA1549; KRAS; KSR1; KSR2; LCK; LMNA; LYN; MAP2K1; MAP2K2; MAPK1; MAPK3; MARK3; MPRIP; NOLC1; NRAS; PAG1; PAPD7; PAPSS1; PDCD1; PDCD1LG2; PEBP1; PECAM1; PIK3CA; PIK3R1; PRKACB; PRKACG; PRKAR1A; PRKAR1B; PRKAR2A; PRKAR2B; PTK2; PTPN11; PTPN22; PTPN6; PXN; QKI; RAF1; RAP1A; RAP1B; RASA1; RHOA; SHC1; SND1; SRC; TLN1; TNS1; TRAK1; TRIM24; VCL; VEGFA; VWF; YES1; YWHAB; ZAP70; ZC3HAV1; ZYX | 131 |
| CLK3 | NA | 0 |
| ULK3 | GLI1; GLI2; GLI3; IST1; SUFU | 5 |
| SCAMP2 | PLD1; PLD2 | 2 |
| MPI | C12orf5; FBP1; FBP2; GFPT1; GFPT2; GNPDA1; GNPDA2; GPI; HK1; HK2; HK3; HKDC1; PFKFB1; PFKFB2; PFKFB3; PFKFB4; PFKL; PFKM; PFKP; PMM1; PMM2 | 21 |
| FAM219B | NA | 0 |
| COX5A | ATP5C1; ATP5D; ATP5F1; ATP5G3; ATP5H; ATP5J2; COA6; COX4I1; COX4I2; COX5B; COX6A1; COX6A2; COX6B1; COX6B2; COX6C; COX7A2L; COX7B; COX7C; COX8A; CYC1; CYCS; MT-CO1; MT-CO2; MT-CO3; NDUFA12; NDUFA4; NDUFA5; NDUFA6; NDUFA7; NDUFA8; NDUFA9; NDUFAB1; NDUFB10; NDUFB11; NDUFB5; NDUFB6; NDUFB7; NDUFB8; NDUFB9; NDUFS3; NDUFS4; NDUFS6; NDUFS8; NDUFV2; PMPCB; UQCR10; UQCR11; UQCRB; UQCRC1; UQCRC2; UQCRFS1; UQCRH; UQCRHL; UQCRQ | 54 |
| C15orf39 | NA | 0 |
| PPCDC | COASY; ENPP1; ENPP3; PANK1; PANK2; PANK3; PANK4; PPCS | 8 |
| IRF8 | ADAR; B2M; BST2; CD44; CIITA; EGR1; FCGR1A; FCGR1B; GBP1; GBP2; GBP3; GBP4; GBP5; GBP6; GBP7; HLA-A; HLA-B; HLA-C; HLA-DPA1; HLA-DPB1; HLA-DQA1; HLA-DQA2; HLA-DQB1; HLA-DQB2; HLA-DRA; HLA-DRB1; HLA-DRB5; HLA-E; HLA-F; HLA-G; ICAM1; IFI27; IFI30; IFI35; IFI6; IFIT1; IFIT2; IFIT3; IFITM1; IFITM2; IFITM3; IP6K2; IRF9; ISG15; ISG20; MAX; MID1; MT2A; MX1; MX2; MYC; NCAM1; OAS1; OAS2; OAS3; OASL; PML; PSMB8; PTAFR; RNASEL; RSAD2; SAMHD1; SP100; SPI1; STAT2; TRIM10; TRIM14; TRIM17; TRIM2; TRIM21; TRIM22; TRIM25; TRIM26; TRIM29; TRIM3; TRIM31; TRIM35; TRIM38; TRIM45; TRIM46; TRIM48; TRIM5; TRIM6; TRIM62; TRIM68; TRIM8; VCAM1; XAF1; ZBTB17 | 89 |
| IKZF3 | MAPK1;IL2RB;IL2;KRAS;IKZF4;MAPK3;RASA1;SOCS3;LCK;JAK1;SOS1;NRAS;IL2RG;JAK3;SHC1;IL2RA | 16 |
| ERBB2 (HER2) | ADAM17; AREG; ARHGEF11; ARHGEF12; BTC; CBL; CD44; CDC37; CTNNB1; CUL5; DIAPH1; DOCK7; EGF; EGFR; ERBB2IP; ERBB3; ERBB4; EREG; ERRFI1; ESR1; FYN; GAB1; GRB2; GRB7; HBEGF; HRAS; HSP90AA1; IL6; IL6R; JAK2; KRAS; LRPPRC; MAP2K1; MAPK8; MATK; MEMO1; MUC1; MUC4; NRAS; NRG1; NRG2; NRG3; NRG4; PGR; PIK3CA; PIK3CB; PIK3R1; **PIK3R2**; PLCG1; PLCG2; PLXNB1; PRKACA; PTEN; PTK2; PTK6; PTPN11; PTPN12; PTPN18; RHOA; RHOB; RHOC; RND1; RNF41; ROCK1; ROCK2; RPS27A; SEMA4D; SHC1; SHC2; SOS1; SRC; STAT1; STAT3; STUB1; TFAP2A; TFAP2B; TFAP2C; TFF1; TFF2; TFF3; TGFA; TP53; UBA52; UBB; UBC; VEGFA; YES1; YY1 | 88 |
| PSMD3 | ADRM1; AJUBA; AMER1; ANAPC1; ANAPC10; ANAPC11; ANAPC15; ANAPC16; ANAPC2; ANAPC4; ANAPC5; ANAPC7; APC; AURKA; AURKB; AXIN1; AXIN2; BIRC2; BIRC3; BTRC; BUB1B; BUB3; CCNA1; CCNA2; CCND1; CCNE1; CCNE2; CD40LG; CDC16; CDC20; CDC23; CDC25A; CDC26; CDC27; CDC5L; CDC6; CDK1; CDK2; CDKN1A; CDKN1B; CFTR; CKS1B; CSNK1A1; CTNNB1; CUL1; CUL2; CUL3; DHH; DVL1; DVL2; DVL3; EGLN1; EGLN3; EPAS1; FBXL7; FBXO5; FZR1; GLI1; GLI2; GLI3; GMNN; GSK3B; GTSE1; HECW1; HIF1A; HIF3A; HIVEP3; IHH; ITCH; KLHL12; LIMD1; LTA; LTB; LTBR; MAD2L1; MAP3K14; MAPK6; NEDD8; NF1; **NFKB1**; NFKB2; NFKBIA; NFKBIB; NFKBIE; NOTCH4; NUMB; OAZ1; OAZ2; OAZ3; ODC1; ORC1; PAK2; PLK1; PPP2CA; PPP2CB; PPP2R1A; PPP2R1B; PPP2R5A; PPP2R5B; PPP2R5C; PPP2R5D; PPP2R5E; PRICKLE1; PSMA1; PSMA2; PSMA3; PSMA4; PSMA5; PSMA6; PSMA7; PSMA8; PSMB1; PSMB10; PSMB11; PSMB2; PSMB3; PSMB4; PSMB5; PSMB6; PSMB7; PSMB8; PSMB9; PSMC1; PSMC2; PSMC3; PSMC4; PSMC5; PSMC6; PSMD1; PSMD10; PSMD11; PSMD12; PSMD13; PSMD14; PSMD2; PSMD4; PSMD5; PSMD6; PSMD7; PSMD8; PSMD9; PSME1; PSME2; PSME3; PSME4; PSMF1; PTEN; PTTG1; RBX1; REL; RELA; RELB; RFWD2; RNF146; RPN1; RPN2; RPS27A; RUNX3; SHFM1; SHH; SKP1; SKP2; SMURF2; SPOP; SPOPL; SPRED1; SPRED2; SPRED3; SUFU; TCEB1; TCEB2; TNF; TNFRSF11A; TNFRSF12A; TNFRSF13C; TNFRSF1B; TNFSF11; TNFSF12; TNFSF13B; TNFSF14; TNKS; TNKS2; TP53; TP73; TRAF2; TRAF3; UBA52; UBB; UBC; UBD; UBE2C; UBE2D1; UBE2E1; UBLCP1; UCHL5; USP14; VHL; WTIP; WWP1; ZSWIM8 | 200 |
| PIK3R2 | ADRA1A; ADRA1B; ADRA1D; AGT; AGTR1; AKT1; AKT2; AKT3; ANGPT1; ANXA1; APP; ARHGEF1; AVPR1A; AVPR1B; AXL; B2M; BDKRB1; BLNK; BRS3; BTK; CBL; CCKAR; CCKBR; CD28; CD2AP; CDC42; CDIPT; CRK; CRKL; CSF2; CSF2RA; EDN1; EDN2; EDN3; EDNRA; EDNRB; EGFR; **ERBB2**; ERBB3; ERBB4; ESR1; ESR2; F2RL1; F2RL2; FCER1G; FFAR4; FGF1; FGF10; FGF16; FGF18; FGF19; FGF2; FGF20; FGF22; FGF3; FGF4; FGF6; FGF9; FGFR4; FRS2; FYN; GAB1; GAB2; GHSR; GNA11; GNAQ; GNB1; GNB2; GNB3; GNB4; GNB5; GNG11; GNG13; GNG4; GNG7; GNGT1; GNRHR; GPR39; GPR65; GPRC6A; GRAP2; GRM1; GRM5; GRPR; HCK; HCRTR1; HRH1; HTR2B; HTR2C; IGF1R; IL2; IL3; INPP4B; INPP5F; IRS1; IRS2; JAK1; JAK2; JAK3; KDR; KIRREL; KISS1R; KIT; LCK; LCP2; LPAR1; LPAR3; LPAR6; LTB4R; LYN; MCHR1; MCHR2; MLNR; MTOR; NGF; NMBR; NMUR1; NMUR2; NPFFR1; NPFFR2; NPSR1; NTRK1; NTSR1; NTSR2; OPN4; OXTR; P2RY1; P2RY6; PDGFB; PDGFRA; PDGFRB; PDPK1; PIK3C2B; PIK3C2G; PIK3CA; PIK3CB; PIK3CD; PIK3CG; PIK3R1; PIK3R3; PIK3R5; PIK3R6; PIKFYVE; PIP4K2B; PIP5K1A; PIP5K1B; PIP5K1C; PLCG1; PLCG2; PROKR2; PTEN; PTGER1; PTGFR; PTPN11; RAC1; RAC2; RAC3; RET; RHOA; RHOB; RHOBTB1; RHOBTB2; RHOC; RHOD; RHOF; RHOG; RHOH; RHOJ; RHOQ; RHOT1; RHOT2; RHOU; RHOV; SACM1L; SH2D2A; SHC1; SOS1; SRC; SYK; TBXA2R; TEK; TREM2; TRHR; UTS2R; VAV1; VAV2; VAV3; XCR1; YES1; YWHAZ | 190 |
| RAB3A | APBA1; BZRAP1; CASK; CHM; CHML; CPLX1; DNAJC5; GAD1; GAD2; GDI1; GDI2; HSPA8; LIN7A; LIN7B; LIN7C; PPFIA1; PPFIA2; PPFIA3; PPFIA4; RAB10; RAB11A; RAB11B; RAB12; RAB13; RAB14; RAB15; RAB17; RAB18; RAB19; RAB1A; RAB1B; RAB20; RAB21; RAB22A; RAB23; RAB24; RAB25; RAB26; RAB27A; RAB27B; RAB29; RAB2A; RAB2B; RAB30; RAB31; RAB32; RAB33A; RAB33B; RAB34; RAB35; RAB36; RAB37; RAB38; RAB39A; RAB39B; RAB3B; RAB3C; RAB3D; RAB3IL1; RAB40A; RAB40B; RAB40C; RAB41; RAB42; RAB43; RAB44; RAB4A; RAB4B; RAB5A; RAB5B; RAB5C; RAB6A; RAB6B; RAB7A; RAB7B; RAB8A; RAB8B; RAB9A; RAB9B; RABGGTA; RABGGTB; RIMS1; RIMS2; RPH3A; SLC17A7; SLC18A2; SLC18A3; SLC32A1; SNAP25; STX1A; SYN1; SYN2; SYN3; SYT1; UNC13B; VAMP2 | 96 |

**Supplementary Table 6.** Number of significant pCHi-C interactions and captured promoters identified by group.

| **Cell type** | **Group** | **N of biological replicates** | **Significant interactions*** | **N of captured promoters** |
| --- | --- | --- | --- | --- |
| CD4+ T cells | Total | 15 | 81624 | 8193 |
|  | Patients | 10 | 67573 | 7356 |
|  | Controls | 5 | 89794 | 9315 |
| CD14+ monocytes | Total | 15 | 74853 | 7024 |
|  | Patients | 10 | 64333 | 6654 |
|  | Controls | 5 | 67041 | 7908 |

*Significant interactions are defined as those with CHICAGO score > 5

**Supplementary Table 7**. Differentially expressed genes in SSc vs. controls CD4^+^ T cells.

| **Gene** | **log_2_FC** | **log_2_CPM** | **FDR** |
| --- | --- | --- | --- |
| *GPR15* | -2,86 | 5,53 | 4,57E-05 |
| *NUAK2* | -3,44 | 6,33 | 4,57E-05 |
| *LRRN3* | -2,53 | 5,57 | 2,08E-04 |
| *RPS4Y1* | -11,63 | 5,30 | 2,08E-04 |
| *DDX3Y* | -11,47 | 5,13 | 2,08E-04 |
| *KDM5D* | -11,10 | 4,76 | 2,08E-04 |
| *USP9Y* | -10,75 | 4,41 | 2,14E-04 |
| *UTY* | -9,26 | 2,90 | 6,12E-04 |
| *MOSPD2* | -1,07 | 4,48 | 7,00E-04 |
| *GPR55* | -1,38 | 3,69 | 3,25E-03 |
| *PRKY* | -8,53 | 4,06 | 8,17E-03 |
| *PLXNB2* | 1,93 | 2,64 | 1,89E-02 |
| *ZNF208* | -0,88 | 4,24 | 1,92E-02 |
| *B3GAT1* | 4,72 | 3,94 | 2,39E-02 |
| *C1orf21* | 4,45 | 3,55 | 2,88E-02 |
| *TMEM184C* | -0,63 | 4,53 | 2,88E-02 |
| *ITGAX* | 2,94 | 2,77 | 2,88E-02 |
| *FGFBP2* | 5,63 | 5,29 | 3,03E-02 |
| *HLA-DPB1* | 1,28 | 4,91 | 3,05E-02 |
| *IL6R* | -0,50 | 8,21 | 3,38E-02 |
| *CALR* | 0,43 | 8,39 | 3,84E-02 |
| *FCRL6* | 4,51 | 4,69 | 3,87E-02 |
| *TRPS1* | 0,60 | 5,03 | 3,87E-02 |
| *GSE1* | 0,72 | 6,18 | 3,87E-02 |
| *ASF1A* | -0,48 | 5,21 | 3,88E-02 |
| *GZMH* | 5,05 | 6,00 | 4,24E-02 |
| *RAB31* | 1,86 | 2,14 | 4,33E-02 |
| *BEX3* | 0,92 | 4,44 | 6,33E-02 |
| *TP53INP2* | 1,59 | 3,27 | 6,46E-02 |
| *NBPF15* | -0,52 | 7,13 | 6,83E-02 |
| *ADGRG1* | 4,32 | 5,44 | 6,83E-02 |
| *VPS9D1* | 0,47 | 5,40 | 6,83E-02 |
| *TP53INP1* | 0,66 | 7,26 | 6,83E-02 |
| *ZNF419* | -0,79 | 3,87 | 6,83E-02 |
| *SNTB1* | 0,87 | 5,28 | 6,83E-02 |
| *ZEB2* | 3,43 | 5,48 | 6,83E-02 |
| *VAV3* | 1,15 | 4,52 | 6,83E-02 |
| *C12orf75* | 1,12 | 4,68 | 6,83E-02 |
| *FGR* | 2,93 | 4,32 | 6,83E-02 |
| *YBX3* | 1,42 | 3,77 | 6,99E-02 |
| *ZMIZ1* | 0,65 | 6,07 | 7,00E-02 |
| *YIPF4* | -0,48 | 5,84 | 7,39E-02 |
| *FCRL3* | 1,07 | 5,99 | 7,39E-02 |
| *FADS2* | 3,26 | 3,14 | 7,39E-02 |
| *LGR6* | 4,52 | 2,51 | 7,39E-02 |
| *CD36* | 5,11 | 0,98 | 7,39E-02 |
| *ADAM28* | 1,62 | 2,24 | 7,39E-02 |
| *MS4A1* | 1,52 | 3,59 | 7,39E-02 |
| *VCAN* | 2,25 | 2,40 | 7,39E-02 |
| *TMEM119* | 1,85 | 1,87 | 8,43E-02 |
| *NECTIN2* | 4,41 | 0,39 | 8,43E-02 |
| *CDC42BPB* | -0,69 | 4,49 | 8,83E-02 |
| *GNLY* | 3,78 | 6,66 | 8,83E-02 |
| *ITGAM* | 2,40 | 5,55 | 8,83E-02 |
| *CD74* | 0,67 | 8,65 | 9,17E-02 |
| *IL7R* | -0,41 | 11,12 | 9,17E-02 |
| *DUSP6* | 1,53 | 2,87 | 9,23E-02 |
| *PIK3AP1* | 1,72 | 2,79 | 9,23E-02 |
| *FBLN5* | -1,24 | 4,31 | 9,23E-02 |
| *GFOD1* | 1,54 | 3,14 | 9,23E-02 |
| *COL5A3* | -1,04 | 4,24 | 9,52E-02 |
| *GNB4* | 2,15 | 1,55 | 9,52E-02 |

*CPM* Counts per million, *FC* fold change, *FDR* false discovery rate.

**Supplementary Table 8**. Differentially expressed genes in SSc vs. controls CD14^+^ T cells.

| **Genes** | **log_2_FC** | **log_2_CPM** | **FDR** |
| --- | --- | --- | --- |
| *SEMA6B* | -3,14 | 3,26 | 8,26E-05 |
| *CLEC10A* | -1,03 | 6,08 | 7,16E-04 |
| *STAG3* | -1,93 | 0,13 | 2,78E-03 |
| *RPS4Y1* | -12,01 | 3,77 | 8,56E-03 |
| *KDM5D* | -11,29 | 2,79 | 9,34E-03 |
| *CTTNBP2* | -2,29 | 1,75 | 9,34E-03 |
| *DDX3Y* | -9,83 | 3,60 | 1,68E-02 |
| *CSRNP2* | -0,57 | 4,25 | 1,68E-02 |
| *UTY* | -9,74 | 0,80 | 1,68E-02 |
| *CHAMP1* | -0,59 | 4,15 | 1,68E-02 |
| *TBCC* | 0,62 | 4,51 | 1,68E-02 |
| *RHPN1* | -1,29 | 1,18 | 2,67E-02 |
| *USP9Y* | -8,53 | -0,31 | 4,17E-02 |
| *ACCS* | -0,99 | 5,00 | 4,17E-02 |
| *TIGD2* | -0,92 | 1,94 | 4,23E-02 |
| *ZNF552* | -0,75 | 2,75 | 4,23E-02 |
| *ZNF613* | -1,02 | 2,07 | 4,23E-02 |
| *CYP4F22* | 2,40 | 2,85 | 4,23E-02 |
| *FAM118B* | -0,56 | 4,54 | 4,93E-02 |
| *PRKY* | -8,03 | 0,51 | 5,27E-02 |
| *SUOX* | -0,47 | 4,33 | 5,27E-02 |
| *FPR3* | -1,37 | 3,85 | 5,27E-02 |
| *AHRR* | -4,28 | 2,37 | 5,27E-02 |
| *ZNF2* | -1,12 | 1,48 | 5,27E-02 |
| *CMTM6* | 0,40 | 8,51 | 5,27E-02 |
| *CAD* | -0,54 | 4,51 | 5,27E-02 |
| *BBS2* | -0,48 | 5,48 | 5,28E-02 |
| *DBR1* | -0,67 | 3,90 | 5,28E-02 |
| *NUAK2* | -1,92 | 5,91 | 5,82E-02 |
| *RAB5IF* | 0,51 | 4,85 | 5,82E-02 |
| *SLAMF7* | 1,31 | 5,75 | 5,82E-02 |
| *POLH* | -0,61 | 3,24 | 5,82E-02 |
| *MSH6* | -0,54 | 4,64 | 5,82E-02 |
| *LGALS1* | 0,49 | 8,01 | 5,82E-02 |
| *MRC1* | -1,21 | 2,43 | 6,59E-02 |
| *GADD45B* | 0,86 | 6,27 | 6,59E-02 |
| *GBP2* | 0,75 | 8,52 | 6,59E-02 |
| *KLF10* | 1,12 | 7,94 | 6,59E-02 |
| *RBM12B* | -0,56 | 4,32 | 6,59E-02 |
| *RNF149* | 0,48 | 8,41 | 6,59E-02 |
| *TMEM79* | -0,72 | 3,21 | 6,59E-02 |
| *UBALD2* | 0,70 | 6,57 | 6,59E-02 |
| *RSL24D1* | 0,37 | 6,76 | 6,70E-02 |
| *APOL1* | 0,64 | 5,54 | 6,70E-02 |
| *STX11* | 0,53 | 8,61 | 6,70E-02 |
| *ZNF793* | -2,26 | -0,96 | 6,70E-02 |
| *CLU* | 2,02 | 4,12 | 6,70E-02 |
| *ZNF737* | -1,11 | 1,39 | 6,70E-02 |
| *TMEM86A* | -0,81 | 3,19 | 6,70E-02 |
| *BRD8* | -0,69 | 5,22 | 7,23E-02 |
| *ZNF577* | -0,64 | 3,57 | 7,23E-02 |
| *FXR2* | -0,55 | 4,97 | 7,41E-02 |
| *ZC3H13* | -0,49 | 6,43 | 7,53E-02 |
| *SMURF1* | 0,52 | 5,51 | 7,92E-02 |
| *FKBP1C* | 1,31 | 2,38 | 7,98E-02 |
| *GPR174* | 1,65 | 2,08 | 8,91E-02 |
| *MFHAS1* | 0,95 | 1,70 | 8,91E-02 |
| *EHHADH* | -0,89 | 1,45 | 9,44E-02 |
| *ZNF160* | -0,60 | 4,76 | 9,81E-02 |
| *RAP1B* | 0,44 | 7,44 | 9,81E-02 |
| *CDKN2D* | 0,77 | 6,74 | 9,81E-02 |
| *PID1* | -0,77 | 6,57 | 9,81E-02 |
| *ZNF132* | -1,47 | 0,59 | 9,92E-02 |

*CPM* Counts per million, *FC* fold change, *FDR* false discovery rate.

**Supplementary Table 9**. Gene set enrichment analysis of differentially expressed genes in SSc vs. controls CD4^+^ T cells.

| **Source** | **Term name** | **Term id** | **Adjusted *p-*value** | **Genes** |
| --- | --- | --- | --- | --- |
| GO:BP | cell migration | GO:0016477 | 1,78E-03 | *GPR15,USP9Y,MOSPD2,PLXNB2,ITGAX,IL6R,CALR,ADGRG1,TP53INP1,ZEB2,VAV3,FGR,ZMIZ1,LGR6,VCAN,CDC42BPB,ITGAM,CD74* |
| GO:BP | positive regulation of immune system process | GO:0002684 | 5,04E-03 | *MOSPD2,HLA-DPB1, IL6R, CALR, VAV3, FGR, ZMIZ1, FCRL3, CD36, MS4A1, NECTIN2,ITGAM,CD74,IL7R* |
| GO:BP | positive regulation of leukocyte activation | GO:0002696 | 6,96E-03 | *HLA-DPB1,VAV3,FGR,ZMIZ1,FCRL3,NECTIN2,ITGAM,CD74,IL7R* |
| GO:BP | cell motility | GO:0048870 | 7,85E-03 | *GPR15,USP9Y,MOSPD2,PLXNB2,ITGAX,IL6R,CALR,ADGRG1,TP53INP1,ZEB2,VAV3,FGR,ZMIZ1,LGR6,VCAN,CDC42BPB,ITGAM,CD74* |
| GO:BP | localization of cell | GO:0051674 | 7,85E-03 | *GPR15,USP9Y,MOSPD2,PLXNB2,ITGAX,IL6R,CALR,ADGRG1,TP53INP1,ZEB2,VAV3,FGR,ZMIZ1,LGR6,VCAN,CDC42BPB,ITGAM,CD74* |
| GO:BP | positive regulation of cell activation | GO:0050867 | 8,77E-03 | *HLA-DPB1,VAV3,FGR,ZMIZ1,FCRL3,NECTIN2,ITGAM,CD74,IL7R* |
| GO:BP | leukocyte activation | GO:0045321 | 1,27E-02 | *MOSPD2,ITGAX,HLA-DPB1, IL6R, RAB31, VAV3, FGR, ZMIZ1, FCRL3, CD36, MS4A1,NECTIN2,ITGAM,CD74,IL7R* |
| GO:BP | positive regulation of response to stimulus | GO:0048584 | 1,80E-02 | *MOSPD2,GPR55,HLA-DPB1, IL6R, CALR, ADGRG1, ZEB2, VAV3, FGR, ZMIZ1, FCRL3,LGR6,CD36,MS4A1,NECTIN2,ITGAM,CD74,IL7R,DUSP6,PIK3AP1* |
| GO:BP | positive regulation of gene expression | GO:0010628 | 1,82E-02 | *PLXNB2,ITGAX,HLA-DPB1, IL6R, CALR, ASF1A, TP53INP1, FGR, YBX3, ZMIZ1, CD36,TMEM119,CD74,IL7R* |
| GO:BP | B cell proliferation | GO:0042100 | 2,42E-02 | *VAV3,FCRL3,MS4A1,CD74,IL7R* |
| GO:BP | locomotion | GO:0040011 | 3,29E-02 | *GPR15,USP9Y,MOSPD2,PLXNB2,ITGAX,IL6R,CALR,ADGRG1,TP53INP1,ZEB2,VAV3,FGR,ZMIZ1,LGR6,VCAN,CDC42BPB,ITGAM,CD74* |
| GO:BP | immune effector process | GO:0002252 | 4,67E-02 | *KDM5D,MOSPD2,ITGAX,IL6R,RAB31,VAV3,FGR,FCRL3,CD36,NECTIN2,ITGAM, CD74,IL7R* |
| GO:BP | cell adhesion | GO:0007155 | 4,93E-02 | *PLXNB2,ITGAX,HLA-DPB1, CALR, ADGRG1, VAV3, ZMIZ1, CD36, VCAN, NECTIN2, ITGAM,CD74,IL7R,FBLN5,COL5A3* |
| GO:CC | side of membrane | GO:0098552 | 6,91E-05 | *ITGAX,HLA-DPB1,IL6R,CALR,FCRL6,FGR,CD36,MS4A1,ITGAM,CD74,IL7R,GNB4* |
| GO:CC | cell surface | GO:0009986 | 5,13E-04 | *PLXNB2,ITGAX,HLA-DPB1, IL6R, CALR, FCRL6, FCRL3, CD36, MS4A1, NECTIN2, ITGAM,CD74,IL7R* |
| GO:CC | external side of plasma membrane | GO:0009897 | 9,00E-04 | *ITGAX,IL6R,CALR,FCRL6,CD36,MS4A1,ITGAM,CD74,IL7R* |
| GO:CC | endocytic vesicle membrane | GO:0030666 | 6,00E-03 | *HLA-DPB1,CALR,RAB31,CD36,CD74,IL7R* |
| GO:CC | integral component of lumenal side of endoplasmic reticulum membrane | GO:0071556 | 1,53E-02 | *HLA-DPB1,CALR,CD74* |
| GO:CC | lumenal side of endoplasmic reticulum membrane | GO:0098553 | 1,53E-02 | *HLA-DPB1,CALR,CD74* |
| GO:CC | endocytic vesicle | GO:0030139 | 1,69E-02 | *HLA-DPB1,CALR,RAB31,CD36,GNLY,CD74,IL7R* |
| GO:CC | cell periphery | GO:0071944 | 1,95E-02 | *GPR15,LRRN3,MOSPD2,GPR55,PLXNB2,ITGAX,HLA-DPB1, IL6R, CALR, FCRL6, RAB31,ADGRG1,SNTB1,VAV3,FGR,YIPF4,FCRL3,FADS2,LGR6,CD36,ADAM28,MS4A1,VCAN,TMEM119,NECTIN2,CDC42BPB,ITGAM,CD74,IL7R,PIK3AP1,FBLN5,COL5A3,GNB4* |
| GO:CC | lumenal side of membrane | GO:0098576 | 3,56E-02 | *HLA-DPB1,CALR,CD74* |
| KEGG | Hematopoietic cell lineage | KEGG:04640 | 6,45E-05 | *HLA-DPB1,IL6R,CD36,MS4A1,ITGAM,IL7R* |
| WP | Apoptosis-related network due to altered Notch3 in ovarian cancer | WP:WP2864 | 1,02E-02 | *BEX3,VAV3,YBX3,IL7R* |

Only significant terms with adjusted *p-*value < 0,05 were included.

*GO:BP* Gene ontology biological process, *GO:CC* Gene ontology cellular component, *KEGG* KEGG pathways, *REAC* Reactome pathways, *WK* WikiPathways.

**Supplementary Table 10**. Gene set enrichment analysis of overexpressed genes in CD4^+^ T cells.

| **Source** | **Term name** | **Term id** | **Adjusted *p-*value** | **Genes** |
| --- | --- | --- | --- | --- |
| GO:MF | T cell receptor binding | GO:0042608 | 4,46E-04 | *CD3E,LCK,CD3G* |
| GO:MF | phosphotransferase activity, alcohol group as acceptor | GO:0016773 | 8,56E-04 | *ITK,CAMK4,LCK,ZAP70,SBK1,PRKCQ,TRIB2,TXK,OBSCN,PRKCH,TNIK,DGKA,DYRK2,AAK1,PASK,PIM2,STK17A,TTN,PRKCA,PDK1,PRKACB,ACVR1C,ITPKB,CDC42BPG,MAST4,HKDC1,SPEG,MAPK13,LTBP4,EPHA4,DGKH,LMTK3,MAP3K9,DGKE,PRKCZ,PIK3C2B,PGM2L1,TGFBR3,ACVR2B,EPHA1,NPR2,KALRN* |
| GO:MF | kinase activity | GO:0016301 | 1,03E-03 | *ITK,CAMK4,LCK,ZAP70,SBK1,PRKCQ,CARD11,TRIB2,TXK,OBSCN,PRKCH,TNIK,DGKA,DYRK2,AAK1,PASK,PIM2,STK17A,ALDH18A1,TTN,PRKCA,PKIA,PDK1,PRKACB,AK5,ACVR1C,ITPKB,CDC42BPG,MAST4,HKDC1,SPEG,MAPK13,LTBP4,EPHA4,DGKH,LMTK3,MAP3K9,DGKE,PRKCZ,PIK3C2B,PGM2L1,TGFBR3,ACVR2B,EPHA1,NPR2,KALRN* |
| GO:MF | beta-catenin binding | GO:0008013 | 1,80E-03 | *LEF1,TCF7,AXIN2,RORA* |
| GO:MF | interleukin-2 receptor activity | GO:0004911 | 4,65E-03 | *IL2RA,IL2RB,IL2RG* |
| GO:BP | T cell activation | GO:0042110 | 1,42E-26 | *LEF1,CD40LG,BCL11B,DPP4,CTLA4,TCF7,ITK,GATA3,CAMK4,IL7R,CD3E,CD28,CD6,SIRPG,RORA,CD5,CD27,IL2RA,LCK,ZAP70,CD3G,CD3D,CCR7,PRKCQ,RASGRP1,NLRC3,TESPA1,RHOH,CARD11,LAX1,LY9,NFATC2,SLAMF6,CD2* |
| GO:BP | T cell differentiation | GO:0030217 | 1,89E-23 | *LEF1,BCL11B,CTLA4,TCF7,ITK,GATA3,CAMK4,IL7R,CD3E,CD28,RORA,CD27,IL2RA,LCK,ZAP70,CD3G,CD3D,CCR7,RASGRP1,TESPA1,RHOH,CARD11,LY9,NFATC2,SLAMF6,CD2* |
| GO:BP | lymphocyte activation | GO:0046649 | 2,21E-22 | *LEF1,CD40LG,BCL11B,DPP4,CTLA4,TCF7,ITK,GATA3,CAMK4,IL7R,CD3E,CD28,CD6,SIRPG,RORA,CD5,CD27,IL2RA,LCK,ZAP70,CD3G,CD3D,CCR7,PRKCQ,RASGRP1,NLRC3,TESPA1,RHOH,CARD11,ITM2A,LAX1,LY9,NFATC2,SLAMF6,SLAMF1,CD2* |
| GO:BP | lymphocyte differentiation | GO:0030098 | 6,06E-22 | *LEF1,CD40LG,BCL11B,CTLA4,TCF7,ITK,GATA3,CAMK4,IL7R,CD3E,CD28,RORA,CD27,IL2RA,LCK,ZAP70,CD3G,CD3D,CCR7,RASGRP1,TESPA1,RHOH,CARD11,ITM2A,LY9,NFATC2,SLAMF6,CD2* |
| GO:BP | positive regulation of leukocyte cell-cell adhesion | GO:1903039 | 1,27E-20 | *LEF1,CD40LG,DPP4,CTLA4,GATA3,IL7R,CD3E,CD28,CD6,SIRPG,CD5,CD27,IL2RA,LCK,ZAP70,CCR7,PRKCQ,RASGRP1,SKAP1,TESPA1,RHOH,CARD11* |
| GO:CC | immunological synapse | GO:0001772 | 3,00E-11 | *CD3E,CD28,CD6,LCK,ZAP70,PRKCQ,SKAP1,RHOH,CARD11* |
| GO:CC | plasma membrane | GO:0005886 | 1,19E-09 | *RNF157,CD40LG,DPP4,CTLA4,ITK,VSIG1,TRAT1,CACNA1I,IL7R,MAL,CD3E,CD28,CD6,SIRPG,IGSF9B,CD5,CD27,RASGRF2,IL2RA,LCK,ZAP70,CD3G,ANK3,ITGA6,AQP3,CD3D,CCR7,CD96,ANO9,CD247,PRKCQ,CCR4,RASGRP1,PATJ,TNFRSF25,AMIGO1,TRABD2A,KLRB1,S1PR1,APBA2,SKAP1,LRATD2,RHOH,ENO2,CARMIL2,CARD11,PLCG1,ITM2A,ATP8B2,LAX1,LY9,TSPAN18,TXK,SLC38A1,SLAMF6,SLAMF1,CD2,OBSCN,NMT2,APBB1,ITPR3,CD226,THEM4,LTB,PRKCH,TRAF5,TNIK,SIDT1,DGKA,PLA2G6,CLEC2D* |
| GO:CC | cell surface | GO:0009986 | 1,00E-07 | *RNF157,CD40LG,DPP4,CTLA4,ITK,VSIG1,TRAT1,CACNA1I,IL7R,MAL,CD3E,CD28,CD6,SIRPG,IGSF9B,CD5,CD27,RASGRF2,IL2RA,LCK,ZAP70,SPOCK2,CD3G,ANK3,ITGA6,AQP3,CD3D,CCR7,CD96,ANO9,CD247,PRKCQ,CCR4,RASGRP1,PATJ,TNFRSF25,AMIGO1,TRABD2A,KLRB1,S1PR1,APBA2,SKAP1,LRATD2,RHOH,ENO2,CARMIL2,CARD11,PLCG1,ITM2A,ATP8B2,LAX1,LY9,TSPAN18,TXK,SLC38A1,SEPTIN1,SLAMF6,SLAMF1,CD2,OBSCN,NMT2,APBB1,ITPR3,CD226,THEM4,LTB,PRKCH,TRAF5,TNIK,LTBP3,SIDT1,DGKA,PLA2G6,CLEC2D* |
| GO:CC | alpha-beta T cell receptor complex | GO:0042105 | 3,95E-07 | *CD40LG,CTLA4,IL7R,CD3E,CD28,CD6,CD5,CD27,IL2RA,LCK,ZAP70,CD3G,CD3D,CCR7,CCR4,S1PR1,CARMIL2,SLC38A1,CD2,CD226,TRAF5,CLEC2D,TRAF1* |
| GO:CC | T cell receptor complex | GO:0042101 | 5,07E-06 | *CD40LG,CTLA4,IL7R,CD3E,CD28,CD6,CD5,CD27,IL2RA,CD3G,CD3D,CCR7,CCR4* |
| KEGG | T cell receptor signaling pathway | KEGG:04660 | 2,51E-11 | *CD40LG,CTLA4,ITK,CD3E,CD28,LCK,ZAP70,CD3G,CD3D,CD247,PRKCQ,RASGRP1,CARD11,PLCG1,NFATC2* |
| KEGG | Th1 and Th2 cell differentiation | KEGG:04658 | 1,91E-08 | *GATA3,CD3E,IL2RA,LCK,ZAP70,CD3G,CD3D,CD247,PRKCQ,STAT4,PLCG1,NFATC2* |
| KEGG | Th17 cell differentiation | KEGG:04659 | 3,40E-08 | *GATA3,CD3E,RORA,IL2RA,LCK,ZAP70,CD3G,CD3D,CD247,PRKCQ* |
| KEGG | PD-L1 expression and PD-1 checkpoint pathway in cancer | KEGG:05235 | 2,72E-07 | *CD3E,CD28,LCK,ZAP70,CD3G,CD3D,CD247,PRKCQ,RASGRP1* |
| KEGG | Primary immunodeficiency | KEGG:05340 | 2,42E-06 | *CD40LG,IL7R,CD3E,LCK,ZAP70,CD3D* |
| REAC | Generation of second messenger molecules | REAC:R-HSA-202433 | 1,52E-06 | *ITK,CD3E,LCK,ZAP70,CD3G,CD3D* |
| REAC | Translocation of ZAP-70 to Immunological synapse | REAC:R-HSA-202430 | 4,56E-06 | *CD3E,LCK,ZAP70,CD3G,CD3D* |
| REAC | TCR signaling | REAC:R-HSA-202403 | 6,40E-06 | *ITK,TRAT1,CD3E,LCK,ZAP70,CD3G,CD3D,PRKCQ,CARD11,PLCG1* |
| REAC | Binding of TCF/LEF:CTNNB1 to target gene promoters | REAC:R-HSA-4411364 | 8,07E-05 | *LEF1,TCF7,AXIN2* |
| REAC | Costimulation by the CD28 family | REAC:R-HSA-388841 | 8,58E-05 | *CTLA4,CD3E,CD28,LCK,CD3G,CD3D* |
| WP | T-Cell antigen Receptor (TCR) Signaling Pathway | WP:WP69 | 2,43E-10 | *ITK,GATA3,CD3E,CD28,LCK,ZAP70,CD3G,CD3D,CD247,PRKCQ,SKAP1,CARD11,PLCG1,NFATC2* |
| WP | T-Cell antigen Receptor (TCR) pathway during Staphylococcus aureus infection | WP:WP3863 | 1,44E-08 | *CD40LG,CTLA4,ITK,CD28,LCK,ZAP70,CD3D,PRKCQ,CARD11,PLCG1,NFATC2* |
| WP | T-Cell Receptor and Co-stimulatory Signaling | WP:WP2583 | 9,93E-08 | *CTLA4,ITK,CD28,LCK,ZAP70,RASGRP1,PLCG1,NFATC2,DYRK2* |
| WP | Cancer immunotherapy by PD-1 blockade | WP:WP4585 | 1,67E-05 | *CD3E,LCK,ZAP70,CD3G,CD3D* |
| WP | Inflammatory Response Pathway | WP:WP453 | 3,01E-05 | *CD40LG,CD28,IL2RA,LCK,ZAP70* |

Only first five terms of each source with adjusted *p-*value < 0,05 were included.

*GO:MF* Gene ontology molecular function, *GO:BP* Gene ontology biological process, *GO:CC* Gene ontology cellular component, *KEGG* KEGG pathways, *REAC* Reactome pathways, *WK* WikiPathways.

**Supplementary Table 11**. Gene set enrichment analysis of overexpressed genes in CD14^+^ monocytes.

| **Source** | **Term name** | **Term id** | **Adjusted *p-*value** | **Genes** |
| --- | --- | --- | --- | --- |
| GO:MF | immune receptor activity | GO:0140375 | 4,93E-11 | *FPR2,LILRB2,FPR1,CCR1,CSF3R,C5AR1,IL13RA1,FCER1G,CTSH,IFNGR2,IFNGR1,C3AR1,CSF2RB,HLA-DRA, CCR2,HLA-DPA1,CD74,C5AR2,FCAR,IL17RA,PRLR,IL17RC,FCGR1B,FPR3,CR1,CXCR2,HLA-DRB1, CCRL2, IL31RA,FLT3,FCGR2B,IL1R2,CX3CR1,CXCR1,GFRA2,HLA-DQA1,CMKLR1* |
| GO:MF | identical protein binding | GO:0042802 | 4,91E-10 | *WLS,SERPINA1,CSF1R,TLR8,LILRB2,MEFV,SLC11A1,NLRC4,CDA,TLR4,MARCKS,CREB5,TYROBP,ADAMTSL4,LRRK2,PYGL,TGFBI,KYNU,HMOX1,CST3,FCER1G,IRAK3,LTBR,CARD9,MGST1,VEGFA,GAS7,TLR2,PIK3AP1,LRRK1,CTBP2,DAPK1,LMO2,BTK,TYMP,HHEX,KCTD12,RXRA,NLRP3,INSR,PECAM1,NPL,PSAP,GCA,APLP2,IRF5,WARS,PAK1,CTNNA1,NAGA,SLC31A1,RNF135,TKT,CCDC88A,DPYSL2,PYCARD,AGTRAP,BRI3,IDH1,NACC2,DPYD,TNFRSF1A,HEXB,MFSD1,CASP1,VSIR,CEBPA,GLB1,ANXA2,STAC3,NUDT16,IMPDH1,HVCN1,GLIPR2,DGAT2,ARID3A,FBP1,MYD88,SHTN1,IMPA2,BCL2L2,CAT,ADA2,PRDX3,PADI2,TLR1,ATG7,TLR6,MAFB,HOMER3,SNX2,G6PD,LACTB,HSBP1,GLA,HGF,BEST1,CCR2,S100A11,SDCBP,MCEMP1,CD74,BCL6,TPD52L2,YWHAE,LYZ,RILPL2,PADI4,FTL,JAK2,TRIM8,P2RX7,TESC,SRGAP2,APP,BCL2A1,CEBPB,CLCN5,S100Z,APAF1,MGST2,SCARB2,ABCD1,SUMF1,VIM,S100A6,ENG,UHRF1BP1L,MICU1,RENBP,SAT1,GLUL,FCHO2,MID1IP1,P2RX1,ANXA4,AMPD2,LGALS1,NECTIN1,DNTTIP1,PDGFC,JAML,SCARB1,TOR2A,CPQ,GYG1,STK3,TP53I3,CTSC,TPCN2,EXT1,PLEK,STEAP3,SDC3,BTBD3,TUBA1A,PHC2,SH2B2,SOD2,MYBPC3,SNX8,ABI3,PHETA1,SMIM3,TNFSF10,ALDOA,NAMPT,PLXNA2,OPLAH,MPST,BCAT1,NR6A1,PTGS2,EPHB2,SNCA,CHKA,HIP1,CIDEB,BCL11A,KCTD15,PTX3,AKIRIN2,SMAD1,NECTIN2,TRPM4,RIPK2,SNX33,APCDD1,MYO7A,PPM1H,CCL3,SKIL,JUP,RBBP8,AHR,KCNJ2,XRCC4,NRP2,BAIAP2,ANXA1,TMEM38A,ALDH4A1,IER5,PLCB1,ATF3,NOL3,MGLL,S100P,PTK6,TCF4,IFIT3,ID1,SLAMF8,NFKBIA,MLC1,THBS1,NTSR1,SH3BP4,TMCC3,ZNF618,ADRB2,MMP9,MME,JUN* |
| GO:MF | lipid binding | GO:0008289 | 1,29E-09 | *CD36,CD14,RBP7,S100A8,CD1D,TLR4,CD300LF,PTAFR,WDFY3,ADAP2,LYN,FES,NCF1,FGD2,TLR2,RASGRP4,PRAM1,BTK,RXRA,GAB2,PSAP,SNX10,CCDC88A,ARAP1,SNX30,S100A9,ANXA2,MYOF,DENND1A,ANXA5,STX3,DYSF,TPP1,ARAP3,SBF2,OSBPL11,S1PR3,PPT1,PLA2G7,CPNE2,NPC2,TSPO,TLR1,DGKG,TLR6,SNX2,MCOLN1,PICALM,GSTP1,SDCBP,GSN,ADAP1,RARA,VDR,IQGAP1,P2RX7,PCTP,DAPP1,NRGN,NCF4,SNX11,SCARB2,SGK1,ARHGAP26,SNX21,FCHO2,CAPG,STARD8,ANXA4,PLA2G4A,SCARB1,JAG1,CPNE8,WIPI1,MYO1E,SNX18,PLD1,OPN3,LPAR1,SNX8,LY96,FFAR2,PAQR7,CD1C,SPHK1,SNCA,BPI,HIP1,SESTD1,OXER1,RUFY4,OSBPL1A,SNX33,RPH3A,KCNJ2,RUBCNL,SH3PXD2B,CD300A,ANXA1,ALOX15B,PLCB1,OSBPL5,ABCA1,CHPT1,THBS1,MME,PLA2G4C* |
| GO:MF | carbohydrate binding | GO:0030246 | 9,96E-09 | *CLEC4E,VCAN,ASGR2,FCN1,CLEC7A,HK3,SIGLEC7,CD33,PYGL,CD93,SIGLEC10,CD302,SIGLEC9,LGALS3,TALDO1,MANBA,CLEC4D,PLOD1,ASGR1,FBP1,HK2,MAN2B1,HLA-DRA, G6PD, CLEC10A, CLEC5A, GAA, SIGLEC14,CLEC12B,ENG,CLEC12A,LGALS1,NECTIN1,LGALS9,GALK1,ALDOA,SIGLEC1,CLEC4A,CLEC6A,PTX3,LGALS2,CLEC1A,MGAM,HLA-DRB1,FBXO6,LGALS12,CLEC11A,FUOM,SLC2A8,CLEC4F,CHI3L1,FCER2* |
| GO:MF | pattern recognition receptor activity | GO:0038187 | 2,46E-08 | *CLEC4E,FCN1,CD36,CD14,TLR8,CLEC7A,TLR4,TLR7,PTAFR,TLR2,NOD2,MARCO,SCARB1,LY96* |
| GO:BP | myeloid leukocyte activation | GO:0002274 | 4,31E-90 | *S100A12,SERPINA1,FPR2,FCN1,CD36,CD14,TLR8,LILRB2,CYBB,SLC11A1,FPR1,ALDH3B1,FCGR2A,S100A8,CDA,MNDA,HK3,NFAM1,LILRA2,TLR4,QPCT,SIRPA,TYROBP,LRRK2,CD33,SPI1,LILRB4,CD300LF,TLR7,PTAFR,BST1,C5AR1,PYGL,CD93,HMOX1,CST3,FCER1G,LILRB3,LTBR,LYN,MGST1,FGL2,FES,DOK3,SYK,RAB31,ADGRE2,TLR2,PLAUR,PRAM1,LAT2,BTK,AIF1,TIMP2,GRN,SIGLEC9,SIRPB1,CPPED1,ALOX5,CTSS,CTSZ,ADAM9,TRPM2,PECAM1,GAB2,CTSH,PSAP,GCA,CFP,RAB3D,CYFIP1,PRKCD,LGALS3,GNS,CREG1,PYCARD,ASAH1,BRI3,IDH1,CTSD,PRCP,NOTCH2,HEXB,QSOX1,MANBA,S100A9,FUCA2,ITGAX,LTA4H,CEBPA,IFNGR2,GLB1,ANXA2,IFNGR1,IMPDH1,HAVCR2,METTL7A,HVCN1,CLEC4D,DYSF,COTL1,LAMP2,CAT,ADA2,STXBP2,CKAP4,CRISPLD2,NPC2,MAN2B1,GM2A,PADI2,C3AR1,TLR1,RNASE2,ATG7,ITGB2,OSCAR,TLR6,TNFRSF1B,ATP6V0A1,ARSB,GSTP1,GLA,CCR2,CTSB,CD63,PSEN1,MILR1,S100A11,SDCBP,GSN,MCEMP1,ATP11A,CD74,CFD,ATP8B4,CTSA,HSPA6,GLIPR1,CLEC5A,LYZ,FTL,JAK2,IQGAP1,GAA,APP,SERPINB1,AGPAT2,APAF1,FCAR,RAB44,SIGLEC14,PLEKHO2,HPSE,CLEC12A,P2RX1,PLA2G4A,CMTM6,ADGRE3,IL15,GYG1,CTSC,RAB24,LGALS9,VNN1,FUCA1,NEU1,CEACAM3,PLSCR1,PLD1,RETN,TMEM106A,CXCL8,ANPEP,SYNGR1,FGR,ALDOA,NAMPT,IL18,FTH1,SPHK1,SNCA,BPI,C1QA,ITGAM,PTX3,C3,NECTIN2,HP,CR1,CXCR2,LRG1,CCL3,MGAM,JUP,HYAL2,MMP25,MOSPD2,CD300A,CYSTM1,BATF2,FOLR3,IL31RA,BATF3,CXCL1,VSIG4,FCGR2B,S100P,F2RL1,TNFAIP6,FCGR3B,THBS1,MPO,CXCR1,MMP9,CHI3L1,MME,ADGRG3,SLCO4C1,PPBP* |
| GO:BP | cell activation involved in immune response | GO:0002263 | 3,06E-79 | *S100A12,SERPINA1,FPR2,FCN1,CD36,CD14,LILRB2,CYBB,SLC11A1,FPR1,ALDH3B1,FCGR2A,CLEC7A,S100A8,CDA,MNDA,HK3,NFAM1,LILRA2,TLR4,QPCT,SIRPA,TYROBP,CD86,CD33,PTAFR,BST1,C5AR1,PYGL,CD93,HMOX1,CST3,FCER1G,LILRB3,LYN,MGST1,FGL2,FES,DOK3,SYK,RAB31,ADGRE2,TLR2,LRP1,PLAUR,SEMA4A,PRAM1,LAT2,BTK,HLA-DMB, TIMP2, TNFSF13, GRN, SIGLEC9, SIRPB1, CPPED1, PLCG2, NLRP3, ALOX5, CTSS,CTSZ,TRPM2,PECAM1,GAB2,CTSH,PSAP,GCA,CFP,RAB3D,CYFIP1,PRKCD,LGALS3,GNS,CREG1,PYCARD,ASAH1,BRI3,IDH1,CTSD,PRCP,NOTCH2,HEXB,QSOX1,MANBA,S100A9,FUCA2,LOXL3,ITGAX,LTA4H,GAPT,GLB1,ANXA2,IMPDH1,HAVCR2,METTL7A,HVCN1,CLEC4D,DYSF,COTL1,LAMP2,CAT,ADA2,STXBP2,CKAP4,CRISPLD2,NPC2,MAN2B1,GM2A,PADI2,C3AR1,RNASE2,ATG7,ITGB2,OSCAR,HLA-DRA, TNFRSF1B, ATP6V0A1,ARSB,GSTP1,GLA,CCR2,CTSB,CD63,PSEN1,MILR1,S100A11,SDCBP,GSN,MCEMP1,ATP11A,CD74,BCL6,CFD,RARA,ATP8B4,SWAP70,CD244,CTSA,HSPA6,GLIPR1,CLEC5A,LYZ,HLX,FTL,ICAM1,IQGAP1,GAA,APP,SERPINB1,AGPAT2,APAF1,FCAR,RAB44,SIGLEC14,PLEKHO2,HPSE,CLEC12A,P2RX1,CMTM6,ADGRE3,LGALS1,GYG1,CTSC,RAB24,LGALS9,CD180,VNN1,FUCA1,NEU1,CEACAM3,PLD1,RETN,ANPEP,SYNGR1,FGR,ALDOA,IL18,CD1C,FTH1,BPI,ITGAM,PTX3,C3,HP,CR1,RIPK2,CXCR2,LRG1,CCL3,MGAM,JUP,MMP25,MOSPD2,HLA-DRB1, CD40, CD300A, ANXA1, CYSTM1, FOLR3, NFKBID, CXCL1, FCGR2B, S100P, F2RL1, TNFAIP6, FCGR3B,MPO,BCL3,CXCR1,SH2D1B,MMP9,CHI3L1,MME,ADGRG3,SLCO4C1,PPBP* |
| GO:BP | leukocyte activation involved in immune response | GO:0002366 | 4,27E-78 | *S100A12,SERPINA1,FPR2,FCN1,CD36,CD14,LILRB2,CYBB,SLC11A1,FPR1,ALDH3B1,FCGR2A,CLEC7A,S100A8,CDA,MNDA,HK3,NFAM1,LILRA2,TLR4,QPCT,SIRPA,TYROBP,CD86,CD33,PTAFR,BST1,C5AR1,PYGL,CD93,HMOX1,CST3,FCER1G,LILRB3,LYN,MGST1,FGL2,FES,DOK3,SYK,RAB31,ADGRE2,TLR2,PLAUR,SEMA4A,PRAM1,LAT2,BTK,HLA-DMB, TIMP2, TNFSF13, GRN, SIGLEC9, SIRPB1, CPPED1, PLCG2, NLRP3, ALOX5, CTSS, CTSZ,TRPM2,PECAM1,GAB2,CTSH,PSAP,GCA,CFP,RAB3D,CYFIP1,PRKCD,LGALS3,GNS,CREG1,PYCARD,ASAH1,BRI3,IDH1,CTSD,PRCP,NOTCH2,HEXB,QSOX1,MANBA,S100A9,FUCA2,LOXL3,ITGAX,LTA4H,GAPT,GLB1,ANXA2,IMPDH1,HAVCR2,METTL7A,HVCN1,CLEC4D,DYSF,COTL1,LAMP2,CAT,ADA2,STXBP2,CKAP4,CRISPLD2,NPC2,MAN2B1,GM2A,PADI2,C3AR1,RNASE2,ATG7,ITGB2,OSCAR,HLA-DRA, TNFRSF1B, ATP6V0A1,ARSB,GSTP1,GLA,CCR2,CTSB,CD63,PSEN1,MILR1,S100A11,SDCBP,GSN,MCEMP1,ATP11A,CD74,BCL6,CFD,RARA,ATP8B4,SWAP70,CD244,CTSA,HSPA6,GLIPR1,CLEC5A,LYZ,HLX,FTL,ICAM1,IQGAP1,GAA,SERPINB1,AGPAT2,APAF1,FCAR,RAB44,SIGLEC14,PLEKHO2,HPSE,CLEC12A,P2RX1,CMTM6,ADGRE3,LGALS1,GYG1,CTSC,RAB24,LGALS9,CD180,VNN1,FUCA1,NEU1,CEACAM3,PLD1,RETN,ANPEP,SYNGR1,FGR,ALDOA,IL18,CD1C,FTH1,BPI,ITGAM,PTX3,C3,HP,CR1,RIPK2,CXCR2,LRG1,CCL3,MGAM,JUP,MMP25,MOSPD2,HLA-DRB1, CD40, CD300A, ANXA1, CYSTM1, FOLR3, NFKBID, CXCL1, FCGR2B, S100P, F2RL1, TNFAIP6, FCGR3B,MPO,BCL3,CXCR1,SH2D1B,MMP9,CHI3L1,MME,ADGRG3,SLCO4C1,PPBP* |
| GO:BP | leukocyte activation | GO:0045321 | 3,14E-77 | *S100A12,SERPINA1,FPR2,FCN1,CD36,CD14,TLR8,LILRB2,CYBB,SLC11A1,FPR1,ALDH3B1,FCGR2A,CLEC7A,S100A8,CD1D,CDA,MNDA,HK3,NFAM1,LILRA2,TLR4,QPCT,SIRPA,TYROBP,LRRK2,CD86,CD33,SPI1,LILRB4,CD300LF,TLR7,PTAFR,BST1,C5AR1,PYGL,CD93,HMOX1,CST3,FCER1G,LILRB3,LTBR,LYN,MGST1,FGL2,FES,DOK3,SYK,RAB31,ADGRE2,TLR2,PLAUR,SEMA4A,PRAM1,LAT2,BTK,MEF2C,HLA-DMB, AIF1, TIMP2, HHEX, TNFSF13,GRN,LST1,SIGLEC9,SIRPB1,CPPED1,PLCG2,NLRP3,ALOX5,SRC,CTSS,CTSZ,ADAM9,TRPM2,PECAM1,GAB2,CTSH,PSAP,GCA,CFP,RAB3D,CYFIP1,PRKCD,PAK1,LGALS3,GNS,DUSP3,CREG1,PYCARD,ASAH1,BRI3,IDH1,CTSD,PRCP,NOTCH2,HEXB,QSOX1,MANBA,S100A9,FUCA2,LOXL3,ITGAX,LTA4H,NOD2,VSIR,GAPT,CEBPA,IFNGR2,GLB1,ANXA2,IFNGR1,IMPDH1,SKAP2,HAVCR2,METTL7A,HVCN1,CLEC4D,DYSF,COTL1,MYD88,LAMP2,CAT,ADA2,STXBP2,CKAP4,IRF8,CRISPLD2,NPC2,MAN2B1,GM2A,PADI2,C3AR1,TLR1,RNASE2,ATG7,ITGB2,OSCAR,TLR6,MAFB,FKBP1A,HLA-DRA, WDFY4, TNFRSF1B, ATP6V0A1, ARSB, GSTP1, GLA, TNFSF13B,CCR2,CTSB,CD63,PSEN1,MILR1,S100A11,SDCBP,GSN,MCEMP1,HLA-DPA1, SLC46A2, ATP11A, CD74,BCL6,CFD,RARA,ATP8B4,SWAP70,CD244,CTSA,HSPA6,GLIPR1,PIK3R6,CLEC5A,LYZ,HLX,FTL,JAK2,PRELID1,ICAM1,IQGAP1,GAA,LYL1,APP,POU2F2,SERPINB1,CEBPB,AGPAT2,APAF1,FCAR,RAB44,SIGLEC14,PLEKHO2,HPSE,ZMIZ1,CLEC12A,P2RX1,MERTK,PLA2G4A,CMTM6,ADGRE3,LGALS1,IL15,JAML,GYG1,CTSC,RAB24,LGALS9,CD180,VNN1,FUCA1,CD151,NEU1,HLA-DPB1, CEACAM3, PLSCR1, PLD1, IL1B, RETN, EGR1, TMEM106A,GBA,CXCL8,ANPEP,MMP14,SYNGR1,FGR,ALDOA,SOCS6,TNFAIP8L2,NAMPT,IL18,CLEC4A,HFE,CD83,CD1C,FTH1,SPHK1,SNCA,BPI,C1QA,EGR3,HDAC9,ITGAM,PTX3,C3,NECTIN2,HP,CR1,RIPK2,CXCR2,LRG1,CCL3,MGAM,JUP,CDKN1A,SH3RF1,HYAL2,KLF6,MMP25,AHR,MOSPD2,HLA-DRB1, CD40, CD300A, ANXA1,CYSTM1,BATF2,FOLR3,IL31RA,BATF3,FLT3,NFKBID,CXCL1,VSIG4,FCGR2B,S100P,MAP3K8,SOX4,SLAMF8,F2RL1,TNFAIP6,FCGR3B,THBS1,CTSL,MPO,CCL2,BCL3,CXCR1,SH2D1B,HMGB3,BANK1,MMP9,CHI3L1,MME,ADGRG3,SLCO4C1,PPBP* |
| GO:BP | leukocyte degranulation | GO:0043299 | 7,48E-76 | *S100A12,SERPINA1,FPR2,FCN1,CD36,CD14,LILRB2,CYBB,SLC11A1,FPR1,ALDH3B1,FCGR2A,S100A8,CDA,MNDA,HK3,NFAM1,HCK,QPCT,SIRPA,TYROBP,CD33,PTAFR,BST1,C5AR1,PYGL,CD93,HMOX1,CST3,FCER1G,LILRB3,LYN,MGST1,FGL2,FES,DOK3,SYK,RAB31,ADGRE2,TLR2,PLAUR,PRAM1,LAT2,BTK,TIMP2,GRN,SIGLEC9,SIRPB1,CPPED1,ALOX5,CTSS,CTSZ,TRPM2,PECAM1,GAB2,CTSH,PSAP,GCA,CFP,RAB3D,CYFIP1,PRKCD,LGALS3,GNS,CREG1,PYCARD,ASAH1,BRI3,IDH1,CTSD,PRCP,HEXB,QSOX1,MANBA,S100A9,FUCA2,ITGAX,LTA4H,GLB1,ANXA2,IMPDH1,METTL7A,HVCN1,CLEC4D,COTL1,LAMP2,CAT,ADA2,STXBP2,CKAP4,CRISPLD2,NPC2,MAN2B1,GM2A,PADI2,C3AR1,RNASE2,ATG7,ITGB2,OSCAR,TNFRSF1B,ATP6V0A1,ARSB,GSTP1,GLA,CCR2,CTSB,CD63,PSEN1,MILR1,S100A11,SDCBP,GSN,MCEMP1,ATP11A,CFD,ATP8B4,CTSA,HSPA6,GLIPR1,CLEC5A,LYZ,FTL,IQGAP1,GAA,SERPINB1,AGPAT2,APAF1,FCAR,RAB44,SIGLEC14,PLEKHO2,HPSE,CLEC12A,P2RX1,CMTM6,ADGRE3,GYG1,CTSC,RAB24,LGALS9,VNN1,FUCA1,NEU1,CEACAM3,PLD1,RETN,ANPEP,SYNGR1,FGR,ALDOA,FTH1,BPI,ITGAM,PTX3,C3,HP,CR1,CXCR2,LRG1,CCL3,MGAM,JUP,MMP25,MOSPD2,CD300A,CYSTM1,FOLR3,CXCL1,FCGR2B,S100P,F2RL1,TNFAIP6,FCGR3B,MPO,CXCR1,MMP9,CHI3L1,MME,ADGRG3,SLCO4C1,PPBP* |
| GO:CC | intracellular vesicle | GO:0097708 | 2,26E-61 | *WLS,S100A12,SERPINA1,FPR2,FCN1,CD36,CD14,STAB1,CD163,TLR8,LILRB2,MEFV,CYBB,SLC11A1,FPR1,ALDH3B1,FCGR2A,S100A8,CD1D,CDA,MNDA,HK3,NFAM1,TLR4,HCK,QPCT,SIRPA,TYROBP,LRRK2,MPEG1,SCIMP,CD33,SORT1,TLR7,PTAFR,BST1,C5AR1,PYGL,CD93,CST3,FCER1G,LILRB3,MGST1,FGL2,FES,VEGFA,DOK3,SYK,NCF1,RAB32,MCTP1,RAB31,FGD2,TLR2,LRP1,PLAUR,KIF13A,SLC15A3,RNASE6,BTK,SLC31A2,HLA-DMB,DMXL2,TIMP2,AOAH,GRN,SIGLEC9,SIRPB1,CPPED1,STX11,CORO1C,ALOX5,INSR,SRC,CTSS,CTSZ,TRPM2,PECAM1,CTSH,PSAP,FCGRT,GCA,APLP2,CFP,RAB3D,CYFIP1,PRKCD,LGALS3,CTNNA1,SLC31A1,SNX10,AP1S2,GNS,CREG1,UNC93B1,CCDC88A,RHOQ,PYCARD,RP2,ATP6V1B2,AGTRAP,ASAH1,BRI3,ATP6V0D1,IDH1,CTSD,ATP6V0B,PRCP,HEXB,QSOX1,CHP1,ARAP1,NUMB,MANBA,S100A9,FUCA2,ITGAX,LTA4H,NOD2,IFNGR2,GLB1,ANXA2,MYOF,DENND1A,ADAM15,HLA-DMA, IMPDH1, HAVCR2, METTL7A, HVCN1, STX3, ARRB1,CLEC4D,DYSF,COTL1,TPP1,SBF2,OSBPL11,FKBP15,MYD88,LAMP2,PPT1,TOR4A,CAT,ANKRD50,ADA2,STXBP2,PRDX3,BMF,PRKACA,VAMP3,PLIN3,ZYX,CKAP4,CRISPLD2,NPC2,CLIC4,MAN2B1,GM2A,PADI2,C3AR1,RUSC2,TLR1,RNASE2,ATG7,ITGB2,OSCAR,TLR6,SNX2,HLA-DRA, WDFY4, MCOLN1, PICALM, TNFRSF1B,ATP6V0A1,ARSB,GSTP1,GLA,HGF,CTSB,FIG4,AP2A1,CD63,PSEN1,S100A11,SDCBP,GSN,TMEM150B,DENND3,MCEMP1,HLA-DPA1, TBC1D2, GOLIM4, ATP11A, C9ORF72, CD74, CFD, VPS37C, PLD3, ATP8B4, YWHAE,CTSA,HSPA6,TMEM127,GLIPR1,RGS19,CLEC5A,LYZ,LAPTM4A,ADPRH,RNF13,CXORF21,STX7,NCF2,FTL,JAK2,IQGAP1,GAA,SRGAP2,APP,NRGN,ARRDC4,NCF4,SERPINB1,MARCO,CLCN5,AGPAT2,APAF1,FCAR,RAB11FIP1,RAB44,RHOB,SIGLEC14,SNX11,PLEKHO2,SCARB2,STEAP4,AP2S1,RCBTB2,DIAPH2,HPSE,LHFPL2,VIM,TCN2,UHRF1BP1L,DLG4,DSC2,STS,SNX21,CLEC12A,FCHO2,CDC42EP4,FCGR1A,CAPG,P2RX1,RAB20,TIMP1,CMTM6,ADGRE3,TBC1D12,ITSN1,RAB34,GABARAP,IL15,CLTCL1,SLC1A5,SCARB1,GYG1,CTSC,PRLR,RAB24,WIPI1,TPCN2,VNN1,RASGEF1B,FUCA1,MYO1E,NEU1,SPRED1,STEAP3,HLA-DPB1, CEACAM3, ROGDI,SNX18,FCGR1B,PLD1,TUBA1A,RAB39A,LPAR1,SNX8,RETN,PHETA1,SRGN,ANPEP,MMP14,SYNGR1,FGR,ALDOA,FZD2,DENND6B,LY96,SIGLEC1,DAGLA,HFE,CD1C,FTH1,SPHK1,SNCA,BPI,HIP1,RRAGD,ITGAM,PTX3,SCARF1,BACE1,RUFY4,C3,F13A1,HP,OSBPL1A,CR1,RHOC,SNX33,ZDHHC1,DIPK2A,CXCR2,FSTL3,RILP,MYO7A,LRG1,SKIL,MGAM,RPH3A,JUP,HYAL2,MMP25,MOSPD2,RUBCNL,ACRBP,HLA-DRB1, HBEGF, BAIAP2,SCCPDH,MSR1,CD300A,ANXA1,CYSTM1,FOLR3,PLD4,NME8,DAB2,CXCL1,IFITM3,SERPING1,NRP1,SPIRE1,GHRL,SLC17A9,S100P,CALCRL,ABCA1,F2RL1,MLC1,TNFAIP6,FCGR3B,SLC2A8,THBS1,SH3BP4,CTSL,MPO,HLA-DRB5, EREG, CXCR1, TP53INP2, SPRED2, ADRB2, MMP9, CHI3L1, MME, SPARC, ADGRG3,SLCO4C1,CD9,HLA-DQA1,HLA-DQB1,PPBP* |
| GO:CC | cytoplasmic vesicle | GO:0031410 | 4,62E-61 | *WLS,S100A12,SERPINA1,FPR2,FCN1,CD36,CD14,STAB1,CD163,TLR8,LILRB2,MEFV,CYBB,SLC11A1,FPR1,ALDH3B1,FCGR2A,S100A8,CD1D,CDA,MNDA,HK3,NFAM1,TLR4,HCK,QPCT,SIRPA,TYROBP,LRRK2,MPEG1,SCIMP,CD33,SORT1,TLR7,PTAFR,BST1,C5AR1,PYGL,CD93,CST3,FCER1G,LILRB3,MGST1,FGL2,FES,VEGFA,DOK3,SYK,NCF1,RAB32,MCTP1,RAB31,FGD2,TLR2,LRP1,PLAUR,KIF13A,SLC15A3,RNASE6,BTK,SLC31A2,HLA-DMB,DMXL2,TIMP2,AOAH,GRN,SIGLEC9,SIRPB1,CPPED1,STX11,CORO1C,ALOX5,INSR,SRC,CTSS,CTSZ,TRPM2,PECAM1,CTSH,PSAP,FCGRT,GCA,APLP2,CFP,RAB3D,CYFIP1,PRKCD,LGALS3,CTNNA1,SLC31A1,SNX10,AP1S2,GNS,CREG1,UNC93B1,CCDC88A,RHOQ,PYCARD,RP2,ATP6V1B2,AGTRAP,ASAH1,BRI3,ATP6V0D1,IDH1,CTSD,ATP6V0B,PRCP,HEXB,QSOX1,CHP1,ARAP1,NUMB,MANBA,S100A9,FUCA2,ITGAX,LTA4H,NOD2,IFNGR2,GLB1,ANXA2,MYOF,DENND1A,ADAM15,HLA-DMA, IMPDH1, HAVCR2, METTL7A, HVCN1, STX3, ARRB1,CLEC4D,DYSF,COTL1,TPP1,SBF2,OSBPL11,FKBP15,MYD88,LAMP2,PPT1,TOR4A,CAT,ANKRD50,ADA2,STXBP2,PRDX3,BMF,PRKACA,VAMP3,PLIN3,ZYX,CKAP4,CRISPLD2,NPC2,CLIC4,MAN2B1,GM2A,PADI2,C3AR1,RUSC2,TLR1,RNASE2,ATG7,ITGB2,OSCAR,TLR6,SNX2,HLA-DRA, WDFY4, MCOLN1, PICALM, TNFRSF1B,ATP6V0A1,ARSB,GSTP1,GLA,HGF,CTSB,FIG4,AP2A1,CD63,PSEN1,S100A11,SDCBP,GSN,TMEM150B,DENND3,MCEMP1,HLA-DPA1, TBC1D2, GOLIM4, ATP11A, C9ORF72, CD74, CFD, VPS37C, PLD3, ATP8B4, YWHAE,CTSA,HSPA6,TMEM127,GLIPR1,RGS19,CLEC5A,LYZ,LAPTM4A,ADPRH,RNF13,CXORF21,STX7,NCF2,FTL,JAK2,IQGAP1,GAA,SRGAP2,APP,NRGN,ARRDC4,NCF4,SERPINB1,MARCO,CLCN5,AGPAT2,APAF1,FCAR,RAB11FIP1,RAB44,RHOB,SIGLEC14,SNX11,PLEKHO2,SCARB2,STEAP4,AP2S1,RCBTB2,DIAPH2,HPSE,LHFPL2,VIM,TCN2,UHRF1BP1L,DLG4,DSC2,STS,SNX21,CLEC12A,FCHO2,CDC42EP4,FCGR1A,CAPG,P2RX1,RAB20,TIMP1,CMTM6,ADGRE3,TBC1D12,ITSN1,RAB34,GABARAP,IL15,CLTCL1,SLC1A5,SCARB1,GYG1,CTSC,PRLR,RAB24,WIPI1,TPCN2,VNN1,RASGEF1B,FUCA1,MYO1E,NEU1,SPRED1,STEAP3,HLA-DPB1, CEACAM3, ROGDI,SNX18,FCGR1B,PLD1,TUBA1A,RAB39A,LPAR1,SNX8,RETN,PHETA1,SRGN,ANPEP,MMP14,SYNGR1,FGR,ALDOA,FZD2,DENND6B,LY96,SIGLEC1,DAGLA,HFE,CD1C,FTH1,SPHK1,SNCA,BPI,HIP1,RRAGD,ITGAM,PTX3,SCARF1,BACE1,RUFY4,C3,F13A1,HP,OSBPL1A,CR1,RHOC,SNX33,ZDHHC1,DIPK2A,CXCR2,FSTL3,RILP,MYO7A,LRG1,SKIL,MGAM,RPH3A,JUP,HYAL2,MMP25,MOSPD2,RUBCNL,ACRBP,HLA-DRB1, HBEGF, BAIAP2,SCCPDH,MSR1,CD300A,ANXA1,CYSTM1,FOLR3,PLD4,DAB2,CXCL1,IFITM3,SERPING1,NRP1,SPIRE1,GHRL,SLC17A9,S100P,CALCRL,ABCA1,F2RL1,MLC1,TNFAIP6,FCGR3B,SLC2A8,THBS1,SH3BP4,CTSL,MPO,HLA-DRB5,EREG,CXCR1,TP53INP2,SPRED2,ADRB2,MMP9,CHI3L1,MME,SPARC,ADGRG3,SLCO4C1,CD9,HLA-DQA1,HLA-DQB1,PPBP* |
| GO:CC | secretory granule | GO:0030141 | 6,88E-56 | *S100A12,SERPINA1,FPR2,FCN1,CD36,CD14,LILRB2,CYBB,SLC11A1,FPR1,ALDH3B1,FCGR2A,S100A8,CDA,MNDA,HK3,NFAM1,QPCT,SIRPA,TYROBP,CD33,PTAFR,BST1,C5AR1,PYGL,CD93,CST3,FCER1G,LILRB3,MGST1,FGL2,VEGFA,DOK3,RAB31,TLR2,LRP1,PLAUR,DMXL2,TIMP2,GRN,SIGLEC9,SIRPB1,CPPED1,ALOX5,CTSS,CTSZ,TRPM2,PECAM1,CTSH,PSAP,GCA,APLP2,CFP,RAB3D,CYFIP1,PRKCD,LGALS3,CTNNA1,SNX10,GNS,CREG1,PYCARD,ASAH1,BRI3,IDH1,CTSD,PRCP,HEXB,QSOX1,MANBA,S100A9,FUCA2,ITGAX,LTA4H,GLB1,ANXA2,ADAM15,IMPDH1,METTL7A,HVCN1,STX3,CLEC4D,COTL1,LAMP2,TOR4A,CAT,ADA2,STXBP2,BMF,PRKACA,VAMP3,CKAP4,CRISPLD2,NPC2,MAN2B1,GM2A,PADI2,C3AR1,RNASE2,ATG7,ITGB2,OSCAR,TNFRSF1B,ATP6V0A1,ARSB,GSTP1,GLA,HGF,CTSB,CD63,PSEN1,S100A11,SDCBP,GSN,MCEMP1,ATP11A,CFD,ATP8B4,CTSA,HSPA6,GLIPR1,CLEC5A,LYZ,STX7,NCF2,FTL,IQGAP1,GAA,APP,SERPINB1,AGPAT2,APAF1,FCAR,RAB44,SIGLEC14,PLEKHO2,RCBTB2,HPSE,LHFPL2,CLEC12A,P2RX1,TIMP1,CMTM6,ADGRE3,GYG1,CTSC,RAB24,VNN1,FUCA1,NEU1,CEACAM3,PLD1,RETN,SRGN,ANPEP,SYNGR1,FGR,ALDOA,FTH1,SNCA,BPI,ITGAM,PTX3,C3,F13A1,HP,CR1,CXCR2,FSTL3,LRG1,SKIL,MGAM,RPH3A,JUP,MMP25,MOSPD2,ACRBP,BAIAP2,SCCPDH,CD300A,CYSTM1,FOLR3,CXCL1,SERPING1,GHRL,SLC17A9,S100P,TNFAIP6,FCGR3B,THBS1,CTSL,MPO,CXCR1,MMP9,CHI3L1,MME,SPARC,ADGRG3,SLCO4C1,CD9,PPBP* |
| GO:CC | vesicle | GO:0031982 | 5,66E-51 | *WLS,HNMT,S100A12,SERPINA1,FPR2,FCN1,CD36,CD14,STAB1,CD163,TLR8,LILRB2,MEFV,CYBB,SLC11A1,FPR1,ALDH3B1,FCGR2A,S100A8,CD1D,CDA,MNDA,HK3,NFAM1,TLR4,HCK,QPCT,SIRPA,IL1RN,MARCKS,CREB5,TYROBP,LRRK2,MPEG1,CD86,KCNE3,SLC37A2,SCIMP,CD33,SORT1,LILRB4,TLR7,PTAFR,BST1,C5AR1,CPVL,PYGL,TGFBI,CD93,PLXNB2,CST3,FCER1G,LILRB3,ALDH2,LYN,MGST1,FGL2,SECTM1,FES,VEGFA,DOK3,SYK,NCF1,RAB32,MCTP1,MTMR11,RAB31,FGD2,TLR2,PILRA,LRP1,PLAUR,KIF13A,SLC15A3,LAT2,GNB4,RNASE6,BTK,SLC31A2,HLA-DMB, DMXL2, TIMP2, AOAH, TNFSF13, GRN, SIGLEC9, SIRPB1, CPPED1, STX11, PLCG2,CORO1C,ALOX5,INSR,SRC,CTSS,CTSZ,ADAM9,TRPM2,BLVRB,PECAM1,TTYH3,ENTPD1,CTSH,PSAP,FCGRT,SCPEP1,GCA,APLP2,WARS,CFP,RAB3D,CYFIP1,PRKCD,LGALS3,CTNNA1,NAGA,SLC31A1,SNX10,PGD,AP1S2,GNS,TKT,CREG1,UNC93B1,CCDC88A,DPYSL2,RHOQ,PYCARD,RP2,ATP6V1B2,AGTRAP,ASAH1,BRI3,ATP6V0D1,IDH1,CTSD,TALDO1,CTNND1,ATP6V0B,ATP6V1A,PRCP,RALB,HEXB,QSOX1,CHP1,ARAP1,NUMB,MANBA,S100A9,SOGA1,FUCA2,ITGAX,LTA4H,NOD2,NIBAN2,LIN7A,IFNGR2,CYBRD1,GLB1,ANXA2,MYOF,DENND1A,ADAM15,HLA-DMA, IMPDH1, ANXA5, HAVCR2, NAGK, METTL7A, HVCN1, STX3, ARRB1, CLEC4D,DYSF,COTL1,TNFRSF8,GLIPR2,TPP1,PLOD1,SBF2,OSBPL11,FKBP15,FBP1,ALDH1A1,MYD88,VASP,TSPAN4,LAMP2,PPT1,TOR4A,ST3GAL6,CAT,ANKRD50,ADA2,CPNE2,STXBP2,PRDX3,BMF,CNTLN,PRKACA,VAMP3,PLIN3,ZYX,CKAP4,CRISPLD2,NPC2,CLIC4,MAN2B1,GM2A,TSPO,PADI2,C3AR1,RUSC2,TLR1,RNASE2,ATG7,ITGB2,OSCAR,TLR6,CAPZA2,RNPEP,ACSL4,SNX2,HLA-DRA, WDFY4, MCOLN1, PICALM, TNFRSF1B, ATP6V0A1,ARSB,GSTP1,G6PD,GLA,HGF,CTSB,FIG4,AP2A1,CD63,PSEN1,S100A11,SDCBP,GSN,TMEM150B,DENND3,MCEMP1,HLA-DPA1, TBC1D2, GOLIM4, NID1, ATP11A, C9ORF72, CLIC1, CD74, CFD, VPS37C, PLD3, ATP8B4,YWHAE,CTSA,HSPA6,TMEM127,GLIPR1,RGS19,CLEC5A,LYZ,LAPTM4A,ADPRH,RNF13,GBGT1,CXORF21,STX7,NCF2,FTL,JAK2,PTGS1,CPD,ICAM1,IQGAP1,SEMA3C,GAA,SMPDL3A,RIPOR1,SRGAP2,COMT,APP,SERPINB8,NRGN,ARRDC4,NCF4,SERPINB1,MYO1F,MARCO,CLCN5,AGPAT2,APAF1,FCAR,RAB11FIP1,RAB44,RHOB,SIGLEC14,SPINT1,SNX11,PLEKHO2,SCARB2,CPM,STEAP4,AP2S1,IGFBP7,RNH1,RCBTB2,DIAPH2,HPSE,LHFPL2,VIM,S100A6,NANS,TCN2,UHRF1BP1L,DLG4,GSTO1,DSC2,STS,RENBP,FCGR3A,SNX21,GLUL,CLEC12A,FCHO2,CDC42EP4,FCGR1A,CAPG,BASP1,P2RX1,RAB20,ANXA4,TIMP1,CMTM6,ADGRE3,TBC1D12,LGALS1,ITSN1,RAB34,GABARAP,PDGFC,IL15,CLTCL1,SLC1A5,SCARB1,CPQ,GYG1,GK,CTSC,PRLR,RAB24,BLVRA,CPNE8,HEBP1,WIPI1,TPCN2,VNN1,RASGEF1B,FUCA1,MYO1E,NEU1,SPRED1,STEAP3,HLA-DPB1, CEACAM3, ROGDI, SNX18, PLSCR1, FCGR1B, PLD1, TUBA1A, FAM20C, SOD2, RAB39A, LPAR1, SNX8, RRAS, LAP3,RETN,PHETA1,GALK1,PPM1L,GBA,SRGN,ANPEP,MMP14,SYNGR1,UACA,FGR,TNFSF10,ALDOA,FZD2,TUBB6,DENND6B,LY96,NAMPT,SIGLEC1,MPST,DAGLA,HFE,CD1C,FTH1,SPHK1,SNCA,CDC42BPB,BPI,HIP1,LAMB2,RRAGD,ITGAM,PTX3,SCARF1,BACE1,RUFY4,C3,B3GNT8,F13A1,NECTIN2,HP,OSBPL1A,GNG5,CR1,RHOC,METRNL,CD101,RIPK2,SNX33,CA2,ZDHHC1,DIPK2A,CXCR2,SERINC2,LAMC1,FSTL3,RILP,MYO7A,LRG1,SKIL,MGAM,RPH3A,JUP,HYAL2,SLC15A2,MMP25,MOSPD2,RUBCNL,ACRBP,HLA-DRB1, CD40, HBEGF, BAIAP2,SCCPDH,MSR1,CD300A,ANXA1,CYSTM1,FOLR3,ALOX15B,PLD4,TMEM38A,NME8,DAB2,CXCL1,ANO5,FSCN1,IFITM3,SERPING1,PLCB1,MYO7B,NRP1,SPIRE1,GHRL,SLC17A9,S100P,CALCRL,DDAH2,ABCA1,F2RL1,MLC1,TNFAIP6,FCGR3B,SLC2A8,THBS1,SH3BP4,PRKAR2B,CTSL,MPO,HLA-DRB5, EREG, CXCR1, EPS8, TP53INP2, FAM20A, SPRED2, ALPL,ADRB2,MMP9,CHI3L1,MME,SPARC,ADGRG3,SLCO4C1,CD9,HLA-DQA1,FCER2,HLA-DQB1, PPBP, TUBB1* |
| GO:CC | secretory vesicle | GO:0099503 | 3,33E-49 | *S100A12,SERPINA1,FPR2,FCN1,CD36,CD14,LILRB2,CYBB,SLC11A1,FPR1,ALDH3B1,FCGR2A,S100A8,CDA,MNDA,HK3,NFAM1,QPCT,SIRPA,TYROBP,LRRK2,CD33,PTAFR,BST1,C5AR1,PYGL,CD93,CST3,FCER1G,LILRB3,MGST1,FGL2,VEGFA,DOK3,MCTP1,RAB31,TLR2,LRP1,PLAUR,DMXL2,TIMP2,GRN,SIGLEC9,SIRPB1,CPPED1,STX11,ALOX5,CTSS,CTSZ,TRPM2,PECAM1,CTSH,PSAP,GCA,APLP2,CFP,RAB3D,CYFIP1,PRKCD,LGALS3,CTNNA1,SNX10,GNS,CREG1,PYCARD,ASAH1,BRI3,ATP6V0D1,IDH1,CTSD,PRCP,HEXB,QSOX1,MANBA,S100A9,FUCA2,ITGAX,LTA4H,GLB1,ANXA2,ADAM15,IMPDH1,METTL7A,HVCN1,STX3,CLEC4D,COTL1,LAMP2,PPT1,TOR4A,CAT,ADA2,STXBP2,BMF,PRKACA,VAMP3,CKAP4,CRISPLD2,NPC2,MAN2B1,GM2A,PADI2,C3AR1,RNASE2,ATG7,ITGB2,OSCAR,PICALM,TNFRSF1B,ATP6V0A1,ARSB,GSTP1,GLA,HGF,CTSB,CD63,PSEN1,S100A11,SDCBP,GSN,MCEMP1,ATP11A,CFD,ATP8B4,CTSA,HSPA6,GLIPR1,CLEC5A,LYZ,STX7,NCF2,FTL,IQGAP1,GAA,APP,SERPINB1,CLCN5,AGPAT2,APAF1,FCAR,RAB44,SIGLEC14,PLEKHO2,RCBTB2,HPSE,LHFPL2,DLG4,CLEC12A,P2RX1,TIMP1,CMTM6,ADGRE3,GYG1,CTSC,RAB24,VNN1,FUCA1,NEU1,CEACAM3,ROGDI,PLD1,RETN,SRGN,ANPEP,SYNGR1,FGR,ALDOA,FTH1,SNCA,BPI,ITGAM,PTX3,BACE1,C3,F13A1,HP,CR1,CXCR2,FSTL3,LRG1,SKIL,MGAM,RPH3A,JUP,MMP25,MOSPD2,ACRBP,BAIAP2,SCCPDH,CD300A,CYSTM1,FOLR3,CXCL1,SERPING1,GHRL,SLC17A9,S100P,TNFAIP6,FCGR3B,SLC2A8,THBS1,CTSL,MPO,CXCR1,MMP9,CHI3L1,MME,SPARC,ADGRG3,SLCO4C1,CD9,PPBP* |
| KEGG | Tuberculosis | KEGG:05152 | 6,86E-15 | *CLEC4E,CD14,FCGR2A,CLEC7A,TLR4,FCER1G,CARD9,SYK,TLR2,HLA-DMB, SRC, CTSS, ATP6V0D1, CTSD, ATP6V0B,TNFRSF1A,ITGAX,NOD2,IFNGR2,BID,IFNGR1,HLA-DMA,MYD88,LAMP2,TLR1,ITGB2,TLR6,HLA-DRA,ATP6V0A1,HLA-DPA1,CIITA,CD74,VDR,JAK2,CEBPB,APAF1,KSR1,FCGR3A,FCGR1A,HLA-DPB1, IL1B, IL18, SPHK1, ITGAM, C3,CR1,RIPK2,HLA-DRB1,FCGR2B,FCGR3B,HLA-DRB5,HLA-DQB1* |
| KEGG | Phagosome | KEGG:04145 | 5,11E-14 | *CD36,CD14,CYBB,FCGR2A,CLEC7A,TLR4,NCF1,TLR2,HLA-DMB, CTSS, ATP6V1B2, ATP6V0D1, ATP6V0B, ATP6V1A,HLA-DMA,LAMP2,VAMP3,ITGB2,TLR6,HLA-DRA,ATP6V0A1,HLA-DPA1, STX7, NCF2, NCF4, MARCO,FCAR,FCGR3A,FCGR1A,SCARB1,HLA-DPB1,TUBA1A,TUBB6,ITGAM,C3,RILP,HLA-DRB1, MSR1, FCGR2B,FCGR3B,THBS1,CTSL,MPO,HLA-DRB5,HLA-DQB1,TUBB1* |
| KEGG | Leishmaniasis | KEGG:05140 | 1,49E-13 | *CYBB,FCGR2A,TLR4,NCF1,TLR2,HLA-DMB,IFNGR2,IFNGR1,HLA-DMA,MYD88,ITGB2,HLA-DRA,HLA-DPA1, NCF2,JAK2,NCF4,FCGR3A,FCGR1A,HLA-DPB1,IL1B,PTGS2,FOS,ITGAM,C3,CR1,HLA-DRB1, NFKBIA, FCGR3B,HLA-DRB5,JUN,HLA-DQB1* |
| KEGG | Lysosome | KEGG:04142 | 1,53E-13 | *SLC11A1,SORT1,CTSS,CTSZ,CTSH,PSAP,NAGA,AP1S2,GNS,ASAH1,ATP6V0D1,CTSD,ATP6V0B,HEXB,MANBA,FUCA2,GLB1,TPP1,LAMP2,PPT1,NPC2,MAN2B1,GM2A,MCOLN1,ATP6V0A1,ARSB,GLA,CTSB,CD63,CTSA,LAPTM4A,GAA,SCARB2,SUMF1,CLTCL1,CTSC,FUCA1,NEU1,GBA,LIPA,HYAL2,CTSL* |
| KEGG | Rheumatoid arthritis | KEGG:05323 | 1,58E-11 | *TLR4,CD86,VEGFA,TLR2,HLA-DMB,TNFSF13,ATP6V1B2,ATP6V0D1,ATP6V0B,ATP6V1A,HLA-DMA, ITGB2, HLA-DRA,ATP6V0A1,TNFSF13B,HLA-DPA1,ICAM1,IL15,HLA-DPB1,IL1B,CXCL8,IL18,FOS,CXCL2,CCL3,HLA-DRB1,CXCL1,CTSL,HLA-DRB5,CCL2,JUN,HLA-DQB1* |
| REAC | Neutrophil degranulation | REAC:R-HSA-6798695 | 1,63E-60 | *S100A12,SERPINA1,FPR2,FCN1,CD36,CD14,LILRB2,CYBB,SLC11A1,FPR1,ALDH3B1,FCGR2A,S100A8,CDA,MNDA,HK3,NFAM1,QPCT,SIRPA,TYROBP,CD33,PTAFR,BST1,C5AR1,PYGL,CD93,CST3,FCER1G,LILRB3,MGST1,FGL2,DOK3,RAB31,TLR2,PLAUR,TIMP2,GRN,SIGLEC9,SIRPB1,CPPED1,ALOX5,CTSS,CTSZ,TRPM2,PECAM1,CTSH,PSAP,GCA,CFP,RAB3D,CYFIP1,PRKCD,LGALS3,GNS,CREG1,PYCARD,ASAH1,BRI3,IDH1,CTSD,PRCP,HEXB,QSOX1,MANBA,S100A9,FUCA2,ITGAX,LTA4H,GLB1,ANXA2,IMPDH1,METTL7A,HVCN1,CLEC4D,COTL1,LAMP2,CAT,ADA2,CKAP4,CRISPLD2,NPC2,MAN2B1,GM2A,PADI2,C3AR1,RNASE2,ATG7,ITGB2,OSCAR,TNFRSF1B,ATP6V0A1,ARSB,GSTP1,GLA,CTSB,CD63,PSEN1,S100A11,SDCBP,GSN,MCEMP1,ATP11A,CFD,ATP8B4,CTSA,HSPA6,GLIPR1,CLEC5A,LYZ,FTL,IQGAP1,GAA,SERPINB1,AGPAT2,APAF1,FCAR,RAB44,SIGLEC14,PLEKHO2,HPSE,CLEC12A,P2RX1,CMTM6,ADGRE3,GYG1,CTSC,RAB24,VNN1,FUCA1,NEU1,CEACAM3,PLD1,RETN,ANPEP,SYNGR1,FGR,ALDOA,FTH1,BPI,ITGAM,PTX3,C3,HP,CR1,CXCR2,LRG1,MGAM,JUP,MMP25,MOSPD2,CD300A,CYSTM1,FOLR3,CXCL1,S100P,TNFAIP6,FCGR3B,MPO,CXCR1,MMP9,CHI3L1,MME,ADGRG3,SLCO4C1,PPBP* |
| REAC | Immune System | REAC:R-HSA-168256 | 1,67E-52 | *CD300E,CLEC4E,S100A12,SERPINA1,FPR2,FCN1,CD36,CD14,CSF1R,TLR8,LILRA1,LILRB2,MEFV,CYBB,SLC11A1,FPR1,NLRC4,TREM1,ALDH3B1,FCGR2A,CCR1,CLEC7A,S100A8,CD1D,CD300C,CDA,MNDA,HK3,NFAM1,LILRA2,SIGLEC7,TLR4,HCK,LILRA5,QPCT,SIRPA,CD300LB,IL1RN,TYROBP,CSF3R,CD86,CD33,LILRA6,LILRB4,CD300LF,TLR7,PTAFR,BST1,DUSP6,C5AR1,CEBPD,IL13RA1,PYGL,CD93,HMOX1,CST3,FCER1G,IRAK3,LILRB3,LTBR,CARD9,LYN,MGST1,FGL2,SIGLEC10,VEGFA,DOK3,SYK,NCF1,RAB31,LY86,TLR2,PIK3AP1,RASGRP4,PILRA,PLAUR,LAT2,RNASE6,BTK,MEF2C,HLA-DMB, TIMP2, TNFSF13, GRN, SIGLEC9, SIRPB1, CPPED1, PLCG2,NLRP3,ALOX5,SRC,CTSS,CTSZ,TRPM2,PECAM1,GAB2,RNF130,CTSH,PSAP,GCA,IRF5,CFP,RAB3D,CYFIP1,PRKCD,PAK1,LGALS3,RNF135,AP1S2,GNS,DUSP3,CREG1,UNC93B1,PYCARD,ATP6V1B2,ASAH1,BRI3,ATP6V0D1,IDH1,CTSD,TALDO1,PEA15,ATP6V0B,TNFRSF1A,ATP6V1A,PRCP,HEXB,QSOX1,CASP1,MANBA,S100A9,FUCA2,ITGAX,LTA4H,NOD2,IFNGR2,GLB1,ANXA2,IFNGR1,FBXL5,HLA-DMA, IMPDH1, HAVCR2, VAV2, METTL7A,HVCN1,STX3,ARRB1,CLEC4D,COTL1,TNFRSF8,CNPY3,MYD88,VASP,LAMP2,CAT,ADA2,STXBP2,PRKACA,VAMP3,CKAP4,MAP3K3,IRF8,CRISPLD2,NPC2,MAN2B1,GM2A,PADI2,C3AR1,TLR1,RNASE2,ATG7,ITGB2,OSCAR,UBE2D1,TLR6,CSF2RB,CAPZA2,FKBP1A,HLA-DRA, TNFRSF1B, ATP6V0A1, ARSB, GSTP1, GLA,HGF,TNFSF13B,CLEC10A,CCR2,CTSB,AP2A1,CD63,PSEN1,S100A11,SDCBP,GSN,MCEMP1,HLA-DPA1, NKIRAS2,CIITA,ATP11A,CD74,BCL6,CFD,PLD3,ATP8B4,CTSA,TLR5,HSPA6,GLIPR1,CLEC5A,UBE2E2,LYZ,NCF2,FTL,PTPN12,JAK2,TRIM8,MAPKAPK3,ICAM1,IQGAP1,GAA,P2RX7,PVR,C5AR2,RNF217,APP,DAPP1,NCF4,SERPINB1,GAB1,AGPAT2,APAF1,FCAR,RAB44,KSR1,SIGLEC14,PLEKHO2,IL17RA,AP2S1,RNF144B,MAP2K3,HPSE,IFI30,VIM,DLG4,PELI2,GSTO1,FCGR3A,CLEC12A,MRAS,FCGR1A,P2RX1,TIMP1,CMTM6,ADGRE3,IL15,JAML,GYG1,CTSC,PRLR,RAB24,LGALS9,CD180,VNN1,FUCA1,NEU1,IL17RC,SPRED1,NFKBIE,HLA-DPB1, CEACAM3,FCGR1B,PLD1,IL1B,TUBA1A,SOD2,RETN,EGR1,OAS1,NRG1,CXCL8,ANPEP,SYNGR1,LMNB1,FGR,ALDOA,TUBB6,LY96,SIGLEC15,RNF19B,IL18,SIGLEC1,CLEC4A,CD1C,PTGS2,FTH1,FOS,CLEC6A,BPI,C1QA,ITGAM,PTX3,C3,F13A1,NECTIN2,HP,OSBPL1A,CR1,CD101,RIPK2,CXCR2,CXCL2,RILP,LRG1,CCL3,MGAM,DUSP1,JUP,CDKN1A,SH3RF1,MMP25,MOSPD2,HLA-DRB1, HBEGF, BAIAP2, CD300A, ANXA1, CYSTM1, FOLR3, IL31RA,SERPINB2,CXCL10,FBXO6,PLD4,FLT3,IFIT2,CXCL1,FCGR2B,FSCN1,IFITM3,SERPING1,S100P,MAP3K8,IFIT3,IL1R2,NFKBIA,TNFAIP6,FCGR3B,ICAM4,CTSL,MPO,HLA-DRB5, CCL2, EREG, CXCR1, SH2D1B, SPRED2, TNFRSF12A,GFRA2,MMP9,IFIT1,CHI3L1,MME,JUN,WASF1,ADGRG3,SLCO4C1,HLA-DQA1, FCER2, PPBP,FCER1A,TUBB1* |
| REAC | Innate Immune System | REAC:R-HSA-168249 | 2,26E-50 | *CD300E,CLEC4E,S100A12,SERPINA1,FPR2,FCN1,CD36,CD14,TLR8,LILRB2,MEFV,CYBB,SLC11A1,FPR1,NLRC4,TREM1,ALDH3B1,FCGR2A,CLEC7A,S100A8,CDA,MNDA,HK3,NFAM1,TLR4,HCK,QPCT,SIRPA,CD300LB,TYROBP,CD33,TLR7,PTAFR,BST1,DUSP6,C5AR1,PYGL,CD93,CST3,FCER1G,IRAK3,LILRB3,CARD9,LYN,MGST1,FGL2,DOK3,SYK,NCF1,RAB31,LY86,TLR2,RASGRP4,PLAUR,LAT2,RNASE6,BTK,MEF2C,TIMP2,GRN,SIGLEC9,SIRPB1,CPPED1,PLCG2,NLRP3,ALOX5,CTSS,CTSZ,TRPM2,PECAM1,GAB2,CTSH,PSAP,GCA,CFP,RAB3D,CYFIP1,PRKCD,PAK1,LGALS3,RNF135,GNS,DUSP3,CREG1,UNC93B1,PYCARD,ATP6V1B2,ASAH1,BRI3,ATP6V0D1,IDH1,CTSD,ATP6V0B,ATP6V1A,PRCP,HEXB,QSOX1,CASP1,MANBA,S100A9,FUCA2,ITGAX,LTA4H,NOD2,GLB1,ANXA2,IMPDH1,VAV2,METTL7A,HVCN1,CLEC4D,COTL1,CNPY3,MYD88,LAMP2,CAT,ADA2,PRKACA,CKAP4,CRISPLD2,NPC2,MAN2B1,GM2A,PADI2,C3AR1,TLR1,RNASE2,ATG7,ITGB2,OSCAR,UBE2D1,TLR6,CAPZA2,TNFRSF1B,ATP6V0A1,ARSB,GSTP1,GLA,CLEC10A,CCR2,CTSB,CD63,PSEN1,S100A11,SDCBP,GSN,MCEMP1,NKIRAS2,ATP11A,CFD,PLD3,ATP8B4,CTSA,TLR5,HSPA6,GLIPR1,CLEC5A,LYZ,NCF2,FTL,MAPKAPK3,IQGAP1,GAA,P2RX7,C5AR2,APP,NCF4,SERPINB1,AGPAT2,APAF1,FCAR,RAB44,SIGLEC14,PLEKHO2,MAP2K3,HPSE,PELI2,FCGR3A,CLEC12A,FCGR1A,P2RX1,CMTM6,ADGRE3,GYG1,CTSC,RAB24,CD180,VNN1,FUCA1,NEU1,CEACAM3,PLD1,IL1B,RETN,ANPEP,SYNGR1,FGR,ALDOA,LY96,SIGLEC15,CLEC4A,FTH1,FOS,CLEC6A,BPI,C1QA,ITGAM,PTX3,C3,HP,CR1,RIPK2,CXCR2,LRG1,MGAM,JUP,MMP25,MOSPD2,BAIAP2,CD300A,CYSTM1,FOLR3,PLD4,CXCL1,SERPING1,S100P,MAP3K8,NFKBIA,TNFAIP6,FCGR3B,CTSL,MPO,CXCR1,MMP9,CHI3L1,MME,JUN,WASF1,ADGRG3,SLCO4C1,PPBP,FCER1A* |
| REAC | Toll-like Receptor Cascades | REAC:R-HSA-168898 | 8,14E-10 | *S100A12,CD36,CD14,TLR8,S100A8,TLR4,TLR7,DUSP6,IRAK3,LY86,TLR2,BTK,MEF2C,PLCG2,CTSS,DUSP3,UNC93B1,S100A9,NOD2,CNPY3,MYD88,TLR1,ITGB2,UBE2D1,TLR6,CTSB,NKIRAS2,TLR5,MAPKAPK3,APP,MAP2K3,PELI2,CD180,LY96,FOS,BPI,ITGAM,RIPK2,MAP3K8,NFKBIA,CTSL,JUN* |
| REAC | Interleukin-10 signaling | REAC:R-HSA-6783783 | 5,02E-09 | *FPR1,CCR1,IL1RN,CD86,PTAFR,TNFRSF1A,TNFRSF1B,CCR2,ICAM1,TIMP1,IL1B,CXCL8,IL18,PTGS2,CXCL2,CCL3,CXCL10,CXCL1,IL1R2,CCL2,FCER2* |
| WP | TYROBP Causal Network | WP:WP3945 | 3,53E-14 | *SLC7A7,TYROBP,IGSF6,KCNE3,IL13RA1,ADAP2,RBM47,CXCL16,RNASE6,PYCARD,DPYD,LOXL3,ITGAX,GAPT,FKBP15,NPC2,ITGB2,TNFRSF1B,SH2B3,HLX,NCF2,LYL1,LHFPL2,CAPG,SLC1A5,NRROS,PLEK,TMEM106A,IL18,ITGAM,C3* |
| WP | Microglia Pathogen Phagocytosis Pathway | WP:WP3937 | 5,62E-08 | *CYBB,TREM1,SIGLEC7,HCK,TYROBP,FCER1G,LYN,SYK,NCF1,PLCG2,VAV2,ITGB2,PIK3R6,NCF2,NCF4,FCGR1A,C1QA,ITGAM,C1QB* |
| WP | IL1 and megakaryocytes in obesity | WP:WP2865 | 5,75E-08 | *TLR2,TIMP2,NLRP3,S100A9,MYD88,PLA2G7,TLR1,ICAM1,TIMP1,IL1B,IL18,HBEGF,CCL2,MMP9,FCER1A* |
| WP | Vitamin D Receptor Pathway | WP:WP2877 | 6,52E-06 | *CD14,LRRC25,TREM1,S100A8,KLF4,SLC37A2,SLC8A1,CLMN,TIMP2,RXRA,ALOX5,IRF5,S100A9,CEBPA,IRF8,G6PD,BCL6,VDR,SERPINB1,NINJ1,STEAP4,ABCD1,S100A6,STS,LGALS9,CYP2S1,MXD1,ITGAM,CASP5,CDKN1A,G0S2,HLA-DRB1,CD40,ADAMTS5,THBD,ASAP2,ID1,CRACR2B,CDKN2B,ADRB2,CD9* |
| WP | Human Complement System | WP:WP2806 | 2,49E-05 | *FCN1,FPR1,C5AR1,CD93,TLR2,PLAUR,CFP,ITGAX,GNA15,PRKACA,C3AR1,ITGB2,CFD,ICAM1,C5AR2,FCGR3A,PTX3,C3,F13A1,CR1,ADM,LAMC1,CD40,VSIG4,SERPING1,THBS1,FCER2* |

Only first five terms of each source with adjusted *p-*value < 0,05 were included.

*GO:MF* Gene ontology molecular function, *GO:BP* Gene ontology biological process, *GO:CC* Gene ontology cellular component, *KEGG* KEGG pathways, *REAC* Reactome pathways, *WK* WikiPathways.

**Supplementary Table 12**. Differential expression corresponding to genes with significant interactions overlapping SSc GWAS loci in CD4^+^ T cells vs CD14^+^ monocytes.

| **Chr** | **Bp (start - end)** | **GWAS locus** | **Gene** | **log_2_FC** | **log_2_CPM** | **FDR** |
| --- | --- | --- | --- | --- | --- | --- |
| 1 | 167445635 - 167465040 | rs2056626 *(CD247)* | *CD247* | 7,49 | 7,26 | 4,00E-210 |
|  |  |  | *CREG1* | -3,68 | 6,98 | 1,32E-325 |
| 2 | 190642047 - 190698201 | rs16832798 *(NAB1)* | *MFSD6* | 0,29 | 5,73 | 2,29E-02 |
|  |  |  | *NEMP2* | 2,03 | 3,63 | 3,16E-61 |
|  |  |  | *HIBCH* | 0,33 | 3,46 | 7,38E-03 |
|  |  |  | *INPP1* | -1,88 | 3,85 | 1,29E-39 |
| 2 | 191035723 - 191108308 | rs3821236 *(STAT4)* | *STAT4* | 7,05 | 6,43 | 1,00E-304 |
|  |  |  | *NABP1* | -0,31 | 7,08 | 2,86E-03 |
| 3 | 58084620 - 58482701 | rs4076852 *(FLNB -DNASE1L3-PXK)* | *RPP14* | 0,13 | 4,03 | 2,46E-01 |
|  |  |  | *KCTD6* | -0,99 | 3,14 | 3,51E-12 |
| 3 | 119384733 - 119546340 | rs9884090 *(POGLUT1-TIMMDC1-CD80- ARHGAP31)* | *TMEM39A* | -0,65 | 4,95 | 1,03E-11 |
|  |  |  | *POGLUT1* | 0,41 | 5,23 | 6,48E-05 |
| 3 | 160002484 - 160030580 | rs589446 *(IL12A)* | *SMC4* | 1,12 | 5,89 | 9,27E-34 |
|  |  |  | *IFT80* | 2,38 | 3,98 | 1,35E-114 |
| 4 | 960523 - 990021 | rs11724804 *(DGKQ)* | *GAK* | -0,34 | 7,52 | 1,74E-08 |
|  |  |  | *TMEM175* | -0,28 | 5,65 | 5,64E-03 |
|  |  |  | *FGFRL1* | 0,25 | 4,68 | 1,50E-01 |
| 4 | 102477892 - 102615256 | rs230534 *(NFKB1)* | *SLC39A8* | 1,84 | 4,38 | 3,18E-26 |
|  |  |  | *NFKB1* | 0,04 | 7,21 | 5,86E-01 |
|  |  |  | *UBE2D3* | -0,50 | 8,69 | 3,89E-17 |
|  |  |  | *CISD2* | 0,30 | 4,03 | 1,73E-02 |
|  |  |  | *SLC9B1* | NA | NA | NA |
|  |  |  | *BDH2* | 1,94 | 3,43 | 6,09E-35 |
| 8 | 60638547 - 60664239 | rs685985 *(RAB2A-CHD7)* | *ASPH* | -1,62 | 5,25 | 5,49E-37 |
|  |  |  | *SDCBP* | -2,92 | 8,31 | 1,14E-117 |
|  |  |  | *CHD7* | 2,58 | 5,73 | 1,51E-59 |
| 11 | 2311894 - 2363262 | rs2651804 *(TSPAN32,CD81-AS1)* | *TSSC4* | -0,32 | 5,15 | 7,88E-05 |
| 11 | 118704617 - 118875175 | rs11217020 *(DDX6)* | *CXCR5* | 3,21 | 3,14 | 1,05E-09 |
|  |  |  | *UPK2* | NA | NA | NA |
|  |  |  | *DDX6* | 1,14 | 8,08 | 2,38E-83 |
|  |  |  | *IFT46* | 0,25 | 2,71 | 1,08E-01 |
|  |  |  | *ARCN1* | -0,21 | 6,96 | 3,69E-03 |
| 15 | 74739180 - 75148328 | rs1378942 *(CSK)* | *CSK* | -0,97 | 8,39 | 1,38E-28 |
|  |  |  | *CLK3* | -0,18 | 7,26 | 4,00E-03 |
|  |  |  | *ULK3* | 1,50 | 6,32 | 4,45E-47 |
|  |  |  | *SCAMP2* | -0,38 | 7,32 | 8,05E-11 |
|  |  |  | *MPI* | 1,56 | 5,30 | 3,50E-82 |
|  |  |  | *FAM219B* | 0,38 | 6,30 | 1,03E-06 |
|  |  |  | *COX5A* | -1,03 | 5,95 | 3,84E-46 |
|  |  |  | *C15orf39* | -3,64 | 7,55 | 2,83E-169 |
| 16 | 85932852 - 85979945 | rs11117420 *(IRF8)* | *IRF8* | -4,47 | 7,28 | 3,11E-72 |
| 17 | 39747478 - 39933464 | rs883770 *(IKZF3-GSDMB)* | *IKZF3* | 5,58 | 7,12 | 1,16E-34 |
|  |  |  | *ERBB2* | 1,82 | 2,04 | 1,34E-18 |
|  |  |  | *PSMD3* | -0,58 | 6,47 | 2,06E-20 |
| 19 | 18068862 - 18093031 | rs2305743 *(IL12RB1)* | *PIK3R2* | NA | NA | NA |
|  |  |  | *RAB3A* | 0,70  Genes with very low expression that were not analyzed are represented as NA.  *CPM* Counts per million, *FC* fold change, *FDR* false discovery rate. | 1,56 | 1,09E-02 |

**Supplementary Table 13.** Differential interaction values corresponding to genes with significant interactions overlapping SSc GWAS loci in CD4^+^ T cells vs CD14^+^ monocytes.

|  |  |  |  | **Chicdiff median values per baited promoter** | | |
| --- | --- | --- | --- | --- | --- | --- |
| **Chr** | **Bp (start - end)** | **GWAS locus** | **Baited gene promoters** | **log_2_FC** | **log_2_FC SE** | **weighted *p-*adjusted** |
| 1 | 167445635 - 167465040 | rs2056626 *(CD247)* | *CD247* | 2,29 | 0,09 | 2,51E-129 |
|  |  |  | *CREG1* | 0,54 | 0,09 | 1,76E-08 |
| 2 | 190642047 - 190698201 | rs16832798 *(NAB1)* | *MFSD6* | 0,73 | 0,08 | 2,03E-21 |
|  |  |  | *NEMP2* | 0,26 | 0,08 | 2,81E-03 |
|  |  |  | *HIBCH;INPP1* | -0,63 | 0,10 | 6,49E-11 |
| 2 | 191035723 - 191108308 | rs3821236 *(STAT4)* | *STAT4* | 2,07 | 0,11 | 1,73E-70 |
|  |  |  | *NABP1* | 0,96 | 0,10 | 2,73E-20 |
| 3 | 58084620 - 58482701 | rs4076852 *(FLNB -DNASE1L3-PXK)* | *RPP14* | 0,52 | 0,10 | 4,58E-06 |
|  |  |  | *KCTD6* | 0,60 | 0,07 | 7,08E-26 |
| 3 | 119384733 - 119546340 | rs9884090 *(POGLUT1-TIMMDC1-CD80- ARHGAP31)* | *TMEM39A;POGLUT1* | 0,46 | 0,10 | 1,45E-05 |
| 3 | 160002484 - 160030580 | rs589446 *(IL12A)* | *SMC4;IFT80* | -1,28 | 0,12 | 1,27E-23 |
| 4 | 960523 - 990021 | rs11724804 *(DGKQ)* | *GAK;TMEM175* | -0,16 | 0,11 | 1,77E-01 |
|  |  |  | *FGFRL1* | -0,19 | 0,08 | 1,92E-02 |
| 4 | 102477892 - 102615256 | rs230534 *(NFKB1)* | *SLC39A8* | 1,24 | 0,09 | 2,45E-40 |
|  |  |  | *NFKB1* | 1,31 | 0,11 | 7,82E-30 |
|  |  |  | *UBE2D3;CISD2* | 0,85 | 0,12 | 7,61E-12 |
|  |  |  | *SLC9B1* | 0,38 | 0,13 | 2,90E-03 |
|  |  |  | *BDH2* | -0,09 | 0,09 | 2,35E-01 |
| 8 | 60638547 - 60664239 | rs685985 *(RAB2A-CHD7)* | *ASPH* | 1,38 | 0,13 | 1,26E-25 |
|  |  |  | *SDCBP* | 0,84 | 0,17 | 5,19E-06 |
|  |  |  | *CHD7* | 0,11 | 0,07 | 1,71E-01 |
| 11 | 2311894 - 2363262 | rs2651804 *(TSPAN32,CD81-AS1)* | *TSSC4* | -0,38 | 0,09 | 8,43E-05 |
| 11 | 118704617 - 118875175 | rs11217020 *(DDX6)* | *CXCR5* | 0,97 | 0,09 | 1,70E-26 |
|  |  |  | *UPK2* | 0,87 | 0,13 | 1,30E-10 |
|  |  |  | *DDX6* | 1,56 | 0,12 | 2,63E-37 |
|  |  |  | *IFT46;ARCN1* | 0,00 | 0,10 | 1,00E+00 |
| 15 | 74739180 - 75148328 | rs1378942 *(CSK)* | *CSK* | 0,28 | 0,11 | 4,33E-02 |
|  |  |  | *CLK3* | 0,00 | 0,10 | 1,00E+00 |
|  |  |  | *ULK3* | 0,53 | 0,13 | 7,81E-05 |
|  |  |  | *SCAMP2* | 0,41 | 0,07 | 1,37E-07 |
|  |  |  | *MPI* | 0,27 | 0,08 | 2,12E-03 |
|  |  |  | *FAM219B* | 0,42 | 0,09 | 1,32E-05 |
|  |  |  | *COX5A* | 0,74 | 0,09 | 3,02E-13 |
|  |  |  | *C15orf39* | -1,61 | 0,13 | 2,50E-34 |
| 16 | 85932852 - 85979945 | rs11117420 *(IRF8)* | *IRF8* | -1,27 | 0,09 | 1,19E-39 |
| 17 | 39747478 - 39933464 | rs883770 *(IKZF3-GSDMB)* | *IKZF3* | 2,07 | 0,12 | 1,98E-67 |
|  |  |  | *ERBB2* | 0,83 | 0,11 | 8,29E-13 |
|  |  |  | *PSMD3* | 1,89 | 0,19 | 3,75E-24 |
| 19 | 18068862 - 18093031 | rs2305743 *(IL12RB1)* | *PIK3R2* | 0,52 | 0,16 | 2,99E-03 |
|  |  |  | *RAB3A* | -0,37 | 0,12 | 1,69E-03 |

*FC* fold change, *SE* standard error.

**Supplementary Table 14**. Gene set enrichment analysis of genes interacting with SSc GWAS loci overlapping enhancer regions in CD4^+^ T cells.

| **Source** | **Term name** | **Term id** | **Adjusted *p-*value** | **Genes** |
| --- | --- | --- | --- | --- |
| GO:MF | protein C-terminus binding | GO:0008022 | 2,49E-02 | *SDCBP,IFT46,CSK,ERBB2,RAB3A* |
| KEGG | Pancreatic cancer | KEGG:05212 | 3,50E-02 | *NFKB1,ERBB2,PIK3R2* |
| KEGG | Epstein-Barr virus infection | KEGG:05169 | 4,90E-02 | *CD247,NFKB1,PSMD3,PIK3R2* |

Only first five terms of each source with adjusted *p-*value < 0,05 were included.

*GO:MF* Gene ontology molecular function, *KEGG* KEGG pathways.

**Supplementary Table 15**. Gene set enrichment analysis of genes interacting with SSc GWAS loci overlapping enhancer regions in CD14^+^ monocytes.

| **Source** | **Term name** | **Term id** | **Adjusted *p-*value** | **intersections** |
| --- | --- | --- | --- | --- |
| GO:MF | transmembrane receptor protein tyrosine kinase activity | GO:0004714 | 7,22E-03 | *FGFRL1,CSK,CLK3,ERBB2* |
| GO:MF | protein tyrosine kinase activity | GO:0004713 | 1,22E-02 | *FGFRL1,CSK,CLK3,ERBB2* |
| GO:MF | transmembrane receptor protein kinase activity | GO:0019199 | 1,25E-02 | *FGFRL1,CSK,CLK3,ERBB2* |
| GO:MF | protein kinase activity | GO:0004672 | 3,16E-02 | *GAK,FGFRL1,CSK,CLK3,ULK3,ERBB2* |

Only first five terms of each source with adjusted *p-*value < 0,05 were included.

*GO:MF* Gene ontology molecular function.
